# Trans-ancestry genome-wide association study of gestational diabetes mellitus highlights genetic links with type 2 diabetes

**DOI:** 10.1101/2021.10.11.21264235

**Authors:** Natalia Pervjakova, Gunn-Helen Moen, Maria-Carolina Borges, Teresa Ferreira, James P Cook, Catherine Allard, Robin N Beaumont, Mickaël Canouil, Gad Hatem, Anni Heiskala, Anni Joensuu, Ville Karhunen, Soo Heon Kwak, Frederick TJ Lin, Jun Liu, Sheryl Rifas-Shiman, Gudmar Thorleifsson, Toby Andrew, Juha Auvinen, Bishwajit Bhowmik, Amélie Bonnefond, Fabien Delahaye, Ayse Demirkan, Philippe Froguel, Kadri Haller-Kikkatalo, Hildur Hardardottir, Sandra Hummel, Akhtar Hussain, Eero Kajantie, Elina Keikkala, Amna Khamis, Jari Lahti, Tove Lekva, Sanna Mustaniemi, Christine Sommer, Aili Tagoma, Evangelia Tzala, Raivo Uibo, Marja Vääräsmäki, Pia M Villa, Kåre I Birkeland, Luigi Bouchard, Cornelia M Duijn, Sarah Finer, Leif Groop, Esa Hämäläinen, Geoffrey M Hayes, Graham A Hitman, Hak C Jang, Marjo-Riitta Järvelin, Anne Karen Jenum, Hannele Laivuori, Olle Melander, Emily Oken, Kyong Soo Park, Patrice Perron, Rashmi B Prasad, Elisabeth Qvigstad, Sylvain Sebert, Kari Stefansson, Valgerdur Steinthorsdottir, Tiinamaija Tuomi, Marie-France Hivert, Paul W Franks, Mark I McCarthy, Cecilia M Lindgren, Rachel M Freathy, Deborah A Lawlor, Andrew P Morris, Reedik Mägi

## Abstract

Gestational diabetes mellitus (GDM) is associated with increased risk of pregnancy complications and adverse perinatal outcomes. GDM often reoccurs and is associated with increased risk of subsequent diagnosis of type 2 diabetes (T2D). To improve our understanding of the aetiological factors and molecular processes driving the occurrence of GDM, including the extent to which these overlap with T2D pathophysiology, the GENetics of Diabetes In Pregnancy (GenDIP) Consortium assembled genome-wide association studies (GWAS) of diverse ancestry in a total of 5,485 women with GDM and 347,856 without GDM. Through trans-ancestry meta-analysis, we identified five loci with genome-wide significant association (*p*<5×10^−8^) with GDM, mapping to/near *MTNR1B* (*p*=4.3×10^−54^), *TCF7L2* (*p*=4.0×10^−16^), *CDKAL1* (*p*=1.6×10^−14^), *CDKN2A-CDKN2B* (*p*=4.1×10^−9^) and *HKDC1* (*p*=2.9×10^−8^). Multiple lines of evidence pointed to genetic contributions to the shared pathophysiology of GDM and T2D: (i) four of the five GDM loci (not *HKDC1*) have been previously reported at genome-wide significance for T2D; (ii) significant enrichment for associations with GDM at previously reported T2D loci; (iii) strong genetic correlation between GDM and T2D; and (iv) enrichment of GDM associations mapping to genomic annotations in diabetes-relevant tissues and transcription factor binding sites. Mendelian randomisation analyses demonstrated significant causal association (5% false discovery rate) of higher body mass index on increased GDM risk. Our results provide support for the hypothesis that GDM and T2D are part of the same underlying pathology but that, as exemplified by the *HKDC1* locus, there are genetic determinants of GDM that are specific to glucose regulation in pregnancy.

---

Gestational diabetes mellitus (GDM), defined as hyperglycaemia with onset or first recognition during pregnancy, is associated with increased risk of pregnancy complications and adverse perinatal outcomes, including pre-eclampsia, stillbirth, large for gestational age, neonatal hypoglycaemia, preterm birth, low Apgar scores and admission to neonatal intensive care^1–4^. Whilst hyperglycaemia commonly resolves postpartum, GDM often reoccurs^5^ and is associated with subsequent diagnosis of type 2 diabetes (T2D) and coronary heart disease^6,7^. Although the global prevalence of GDM is increasing, it varies according to population characteristics (such as maternal age, ancestry and obesity rates) and the criteria used for screening and diagnosis^8^.

GDM and T2D share both genetic and non-genetic risk factors, including obesity, poor diet and sedentary lifestyle^9,10^. Family studies have demonstrated that women with GDM have 30.1% probability of having at least one parent with T2D, compared to just 13.2% for pregnant women with normal glucose tolerance^11^. Furthermore, women with a history of GDM appear to have a nearly 10-fold higher risk of developing T2D than those with a normo-glycaemic pregnancy^7^. Taken together, these observations support the hypothesis that the two diseases are part of the same underlying pathology, with pregnancy potentially acting as a stress test that reveals women at increased risk of GDM and/or T2D^12,13^.

There have been considerable advances in our understanding of the genetic contribution to T2D through large-scale genome-wide association studies (GWAS) across diverse populations^14–17^. In contrast, despite the observed familial clustering of GDM^18^, most genetic association studies of the disease have focussed on evaluating the impact of previously reported loci for T2D and glycaemic traits in modest sample sizes^19^. The most comprehensive systematic review of genetic susceptibility to GDM (from 23 studies) revealed association with T2D risk variants from seven loci, of which six are related to insulin secretion and one to insulin resistance^20^. A genetic risk score (GRS) of risk variants across 34 loci associated with T2D and/or fasting glucose was significantly associated with GDM and improved predictive power over a model including only clinical variables^21^. Variants associated with both insulin secretion and insulin resistance have also been used to construct an aggregated GRS that was shown to predict GDM risk, with and without adjustment for body mass index (BMI), maternal age, and gestational age, although this score was not compared with established clinical predictors^22^. To date, the largest GWAS of GDM has been undertaken in women from a Korean population, including 468 cases and 1,242 non-diabetic controls in the discovery stage, with an additional 931 cases and 783 non-diabetic controls in the follow-up stage^23^. Two loci were associated with GDM at genome-wide significance (*p*<5×10^−8^), mapping near *MTNR1B* and *CDKAL1*, both of which have also been previously implicated in T2D risk.

To gain novel insight into the genetic architecture of GDM, the GENetics of Diabetes In Pregnancy (GenDIP) Consortium assembled GWAS of diverse ancestry in a total of 5,485 women with GDM and 347,856 women without GDM: the effective sample size was 72.2% European, 13.4% East Asian, 9.9% South Asian, 2.8% Hispanic/Latino and 1.7% African (**Tables S1 and S2**). To maximise sample size, we used a phenotype definition that makes best use of the information available in each study, including data from health records, oral glucose tolerance tests and self-report (**Table S1**). Each GWAS was imputed to reference panels from the 1000 Genomes Project^24^, Haplotype Reference Consortium^25^, or population-specific whole-genome sequence data (**Table S3**). Within each GWAS, GDM association summary statistics were derived for all single nucleotide variants (SNVs) passing quality control after appropriate adjustment to account for population structure (**Supplementary Materials and Methods, Table S3**). With these resources, we aimed to improve our understanding of the aetiological factors and molecular processes driving the occurrence of GDM, including the extent to which these overlap with T2D pathophysiology, and investigate the effects of potential causal metabolic risk factors on the disease through Mendelian randomisation (MR).

We began by aggregating GDM association summary statistics across GWAS through trans-ancestry meta-analysis. The most powerful methods allow for potential allelic effect heterogeneity on disease between ancestry groups that cannot be accommodated in a fixed-effects model^26^. Our primary analysis used MR-MEGA^27^, which models heterogeneity between GWAS by including axes of genetic variation that represent ancestry as covariates in a meta-regression model. We considered three axes of genetic variation that separated the five ancestry groups, but which also revealed finer-scale genetic differences between GWAS of the same ancestry (**Figure S1**). We also conducted trans-ancestry and ancestry-specific fixed-effects meta-analyses. We identified five loci at genome-wide significance in the trans-ancestry meta-regression (**Table 1, Figures S2 and S3**), including the previously reported associations from GDM GWAS at *MTNR1B* (rs10830963, *p*=4.3×10^−54^) and *CDKAL1* (rs9348441, *p*=1.6×10^−14^). The remaining three loci for GDM mapped to/near *TCF7L2* (rs7903146, *p*=4.0×10^−16^), *CDKN2A-CDKN2B* (rs10811662, *p*=4.1×10^−9^) and *HKDC1* (rs9663238, *p*=2.9×10^−8^). Through approximate conditional analyses, conducted using ancestry-matched linkage disequilibrium (LD) reference panels for each GWAS (**Supplementary Materials and Methods**), we observed no evidence for multiple distinct association signals at genome-wide significance at any of the five GDM loci (**Figure S4**).

**Table 1.** Loci attaining genome-wide significant (*p*<5×10^−8^) evidence of association with GDM in trans-ancestry meta-regression (MR-MEGA) of 5,485 cases and 347,856 controls.

| Locus | Lead SNV | Chr | Position (bp, b37) | Alleles | | MR-MEGA $p$-value | Fixed-effects OR (95% CI) |
| --- | --- | --- | --- | --- | --- | --- | --- |
|  |  |  |  | Risk | Other |  |  |
| *MTNR1B* | rs10830963 | 11 | 92,708,710 | G | C | $4.3 \times 10^{-54}$ | 1.41 (1.35-1.47) |
| *TCF7L2* | rs7903146 | 10 | 114,758,349 | T | C | $4.0 \times 10^{-16}$ | 1.22 (1.16-1.27) |
| *CDKAL1* | rs9348441 | 6 | 20,680,678 | A | T | $1.6 \times 10^{-14}$ | 1.13 (1.08-1.18) |
| *CDKN2A-CDKN2B* | rs10811662 | 9 | 22,134,253 | G | A | $4.1 \times 10^{-9}$ | 1.14 (1.09-1.20) |
| *HKDC1* | rs9663238 | 10 | 70,983,629 | G | A | $2.9 \times 10^{-8}$ | 1.14 (1.09-1.19) |

Chr: chromosome. OR: odds-ratio. CI: confidence interval.

We next sought to investigate the impact of differences in ancestry and phenotype definition between GWAS on heterogeneity in allelic effects at GDM loci. To do this, we extended the MR-MEGA meta-regression model to include an additional covariate to represent whether GDM status in the study was confirmed via “a universal blood-based test” (**Supplementary Materials and Methods, Table S1**). Here, we use this term to refer to a blood-based test that was applied to all participants, including a diagnostic oral glucose tolerance test (OGTT) or a screening glucose challenge or fasting glucose test, in contrast to clinician decision, risk factor screening, or a lack of clarity on what basis women did or did not have a diagnostic OGTT. This model enables partitioning of heterogeneity into three components (**Table 2**). The first component captures heterogeneity that is correlated with genetic ancestry (that can be explained by the three axes of genetic variation), which can occur because of differences in the structure of LD between ancestry groups or interactions with lifestyle factors that vary across populations. The second component measures heterogeneity that can be explained by the use of a universal blood-based test to screen for or diagnose GDM. The final component reflects residual heterogeneity due to study design that cannot be explained by the first two components. The greatest evidence of ancestry-correlated heterogeneity (after accounting for the use of a universal blood-based test) was observed at the *CDKAL1* locus (*p*_HET_=3.4×10^−5^), where the lead SNV demonstrated stronger effects on GDM in GWAS of East Asian ancestry than in other populations, despite the risk allele being common in all ancestry groups (**Figure S5, Table S4**). A similar pattern of ancestry-correlated heterogeneity in allelic effects on T2D has been reported at the *CDKAL1* locus^16^. Weaker evidence of ancestry-correlated heterogeneity was observed at the *CDKN2A-CDKN2B* locus (*p*_HET_=0.0022), where there were marked differences in the effects on GDM of the lead SNV between GWAS undertaken in different ancestry groups (**Figure S5, Table S4**). In contrast, there was no evidence of heterogeneity due to phenotype definition for any lead SNV, suggesting that differences in allelic effects between GWAS are more likely due to factors related to genetic ancestry than the use of a blood-based test in all women to screen for or diagnose GDM.

**Table 2.** Source of heterogeneity in allelic effects on GDM between GWAS for lead SNVs derived from meta-regression of 5,485 cases and 347,856 controls.

| Locus | Lead SNV | Heterogeneity source ($p$-value) | | |
| --- | --- | --- | --- | --- |
|  |  | Ancestry | Universal blood-based test | Residual |
| *MTNR1B* | rs10830963 | 0.14 | 0.41 | 0.67 |
| *TCF7L2* | rs7903146 | 0.25 | 0.83 | 0.089 |
| *CDKAL1* | rs9348441 | $3.4 \times 10^{-5}$ | 0.28 | 0.15 |
| *CDKN2A-CDKN2B* | rs10811662 | 0.0022 | 0.45 | 0.26 |
| *HKDC1* | rs9663238 | 0.19 | 0.33 | $5.4 \times 10^{-5}$ |

Of the five GDM loci identified at genome-wide significance in the trans-ancestry meta-regression, four have been previously implicated in T2D susceptibility: *MTNR1B, TCF7L2, CDKAL1* and *CDKN2A-CDKN2B*. In fact, in previously reported trans-ancestry GWAS meta-analyses of 180,834 T2D cases and 1,159,055 controls from the DIAMANTE Consortium^16^, the lead T2D SNV is the same as we report for GDM at *MTNR1B, TCF7L2* and *CDKAL1*, and is in strong linkage disequilibrium (LD) at the *CDKN2A-CDKN2B* locus (rs10811661, *r*^2^=0.91 across diverse populations in the 1000 Genomes Project^24^). To further investigate the genetic correlation between the two diseases, we extracted GDM association summary statistics from our trans-ancestry meta-analysis for lead SNVs at 222 previously reported loci for T2D from the DIAMANTE Consortium^16^ (**Figure 1, Table S5**). We observed a strong positive correlation in log-ORs for the T2D risk allele between the two diseases: Pearson *r*=0.573 (*p*<2.2×10^−16^). There was also a highly significant enrichment of GDM associations at T2D loci (50 of 222 lead SNVs with *p*<0.05 and same direction of effect, binomial test *p*<2.2×10^−16^), indicating that they would be discovered at genome-wide significance with larger effective sample sizes. Indeed, after excluding the four overlapping GDM-T2D loci (**Supplementary Materials and Methods**), a weighted genetic risk score of lead T2D SNVs was significantly associated with GDM (*p*=9.7×10^−123^, pseudo-*R*^2^=2.86%). Extending our analyses, genome-wide, using LD-score regression, we observed strong genetic correlation between GDM and T2D: r_G_ (95% CI): 0.744 (0.052, 1.437). Weaker genetic correlations between GDM and other glycaemic traits were also observed (**Table 3, Table S6**). These results are consistent with sharing of genetic determinants of GDM and T2D, although we acknowledge that LD score regression has limited statistical power because of the relatively small GDM sample size, and we note that the correlation from LD-score regression is not bound by −1 to 1, particularly when power is low.

**Table 3.** Genetic correlation from LD-score regression of GDM with T2D and glycaemic traits.

| Trait | Genetic correlation $r_G$ (95% CI)<sup>a</sup> |
| --- | --- |
| T2D | 0.744 (0.052, 1.437) |
| Fasting glucose | 0.218 (-0.211, 0.648) |
| Fasting insulin | 0.410 (-0.114, 0.934) |
| Fasting proinsulin | 0.336 (-0.321, 0.993) |
| 2hr glucose (adjusted for BMI) | 0.444 (-0.371, 1.260) |
| HbA1c | 0.387 (-0.218, 0.991) |
| HOMA-B | -0.005 (-0.551, 0.541) |
| HOMA-IR | 0.236 (-0.382, 0.854) |

CI: Confidence Interval. T2D: type 2 diabetes. HbA1c: glycated haemoglobin. BMI: body mass index. HOMA-B: homeostasis model assessment of $\beta$ -cell activity; HOMA-IR: homeostasis model assessment of insulin resistance.
<sup>a</sup>Genetic correlation obtained from LD-score regression is not bound by -1 to 1 and estimates can therefore be found outside these limits due to high imprecision caused by factors such as low sample size in the association summary statistics used.

**Figure 1.**
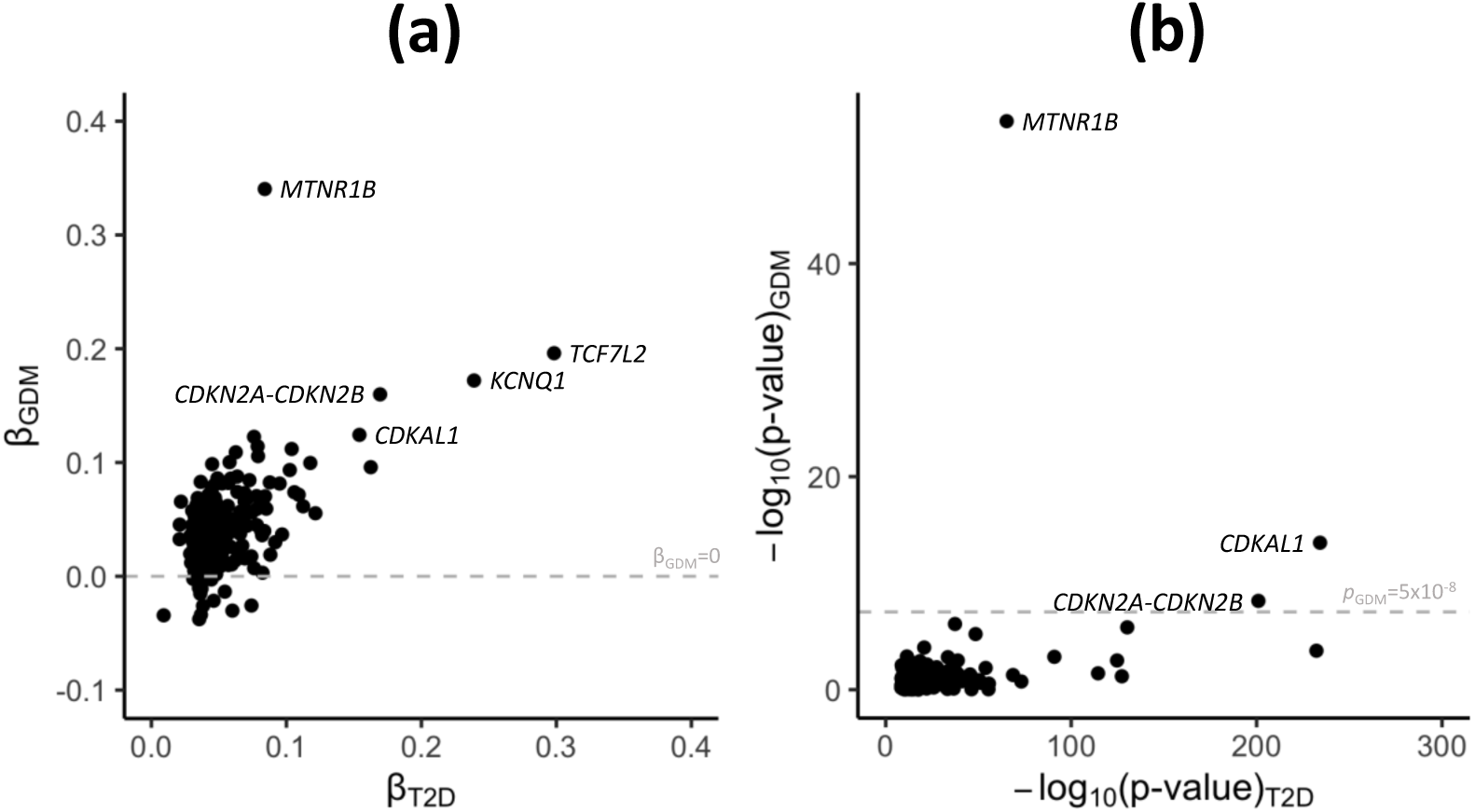
Correlation between GDM and T2D association summary statistics for lead SNVs at previously reported loci for T2D susceptibility. Association summary statistics for GDM were obtained from trans-ancestry GWAS meta-analyses of 5,485 cases and 347,856 controls. Association summary statistics for T2D were obtained from trans-ancestry GWAS meta-analyses of 180,834 cases and 1,159,055 controls from the DIAMANTE Consortium. (a) Allelic effect sizes (log-ORs) for each disease, aligned to the T2D risk allele, from fixed-effects meta-analysis. The grey line represents log-OR of zero for GDM. (b) Association evidence (log10 p-values) for each disease from meta-regression. The grey line represents genome-wide significance (p<5×10-8) for GDM. The lead SNV at the TCF7L2 locus has been removed for ease of presentation (Table S4).

The most obvious difference in allelic effect sizes between GDM and T2D was observed at the *MTNR1B* locus (**Figure 1**). The lead SNV, rs10830963, is the same for both diseases, but the allelic effect on GDM is substantially greater than on T2D: OR (95% CI) for GDM is 1.41 (1.35-1.47) and for T2D is just 1.09 (1.08-1.10). The *MTNR1B* lead SNV is associated, at genome-wide significance, with fasting glycaemic traits in non-diabetic individuals from the Meta-Analysis of Glucose and Insulin-related traits Consortium (MAGIC) Investigators^28,29^. The GDM risk allele at the lead SNV is also associated with higher fasting plasma glucose and 1-hour plasma glucose in pregnant women from the Hyperglycemia and Adverse Pregnancy Outcomes (HAPO) Study^30^. This SNV also has the strongest association of the maternal glucose-raising allele with higher offspring birth weight in women from the Early Growth Genetics Consortium^31^, in line with the known effects of maternal hyperglycaemia on fetal growth. In non-diabetic individuals from the MAGIC Investigators^32^, the *MTNR1B* lead SNV has a much larger impact on fasting glucose than those at *TCF7L2, CDKAL1* and *CDKN2A-CDKN2B*^33^ (**Table S7**). Therefore, the difference in allelic effect sizes between GDM and T2D at *MTNR1B* may reflect the fact that thresholds of fasting plasma glucose used to diagnose GDM are lower than those used to diagnose T2D, meaning that a larger proportion of GDM than T2D cases will have higher fasting glucose that is regulated within the normal range.

To gain insight into the molecular processes and tissues through which GDM association signals are mediated, genome-wide, we then undertook fGWAS enrichment analyses within three categories of functional and regulatory annotations: (i) genic regions^34^; (ii) chromatin immuno-precipitation sequence (ChIP-seq) binding sites for 165 transcription factors^35,36^; and (iii) 13 unique and recurrent chromatin states in four diabetes-relevant tissues (pancreatic islets, liver, adipose, and skeletal muscle)^37^. We observed significant joint enrichment (*p*<0.05) for GDM associations mapping to protein coding exons, binding sites for FOXA2, NFE2 and TFAP2, and chromatin states in adipose tissue and skeletal muscle that mark enhancers and transcribed regions (**Table S8**). FOXA2 is a pioneer factor involved in pancreatic and hepatic development, and T2D association signals have been previously reported to be enriched for FOXA2 binding sites^38^. Skeletal muscle is the most prominent site of insulin-mediated glucose uptake in humans, and enhancers in skeletal muscle have been reported to overlap association signals for metabolic disorders, including T2D, insulin resistance and obesity^39^. These enrichment analyses highlight molecular processes and tissues that are broadly consistent with those important in mediating T2D association signals^16^, although the involvement of pancreatic islets appears to be less prominent for GDM.

In contrast to the other GDM loci reported in this investigation, the lead SNV at the *HKDC1* locus (rs9663238) demonstrates only weak statistical evidence of T2D association in previously reported trans-ancestry GWAS meta-analyses from the DIAMANTE Consortium^16^ (*p*=0.0083, compared with *p*<10^−65^ at the other four loci). GDM risk alleles at variants in strong LD (European ancestry *r*^2^>0.9) with the lead SNV have been previously associated, at genome-wide significance, with higher 2-hour plasma glucose (2HPG) in pregnant women in the HAPO Study and two replication studies of European ancestry^30^, as well as with higher birth weight of first child (likely via greater maternal glucose availability), higher own birth weight (fetal effect independent of the maternal effect on birth weight), and comparative height and body size at age 10 in UK Biobank^40,41^ (**Table S9**). In addition to demonstrating the association of the maternal SNVs at this locus with GDM in the current study, we observed that 99% credible set variants are lead SNVs for *HKDC1* expression quantitative trait loci in a range of tissues in the GTEx Project^42^, including visceral adipose, subcutaneous adipose and pancreas (**Supplementary Materials and Methods, Table S10**). *HKDC1* (Hexokinase Domain Containing 1) catalyzes the phosphorylation of hexose to hexose 6-phosphate and is involved in glucose homeostasis and hepatic lipid accumulation. Haplotypes of variants associated with 2HPG in pregnancy disrupt regulatory element activity and reduce *HKDC1* expression across diverse tissues (including metabolically relevant liver stellate cells and pancreatic islet beta cells), which has been demonstrated to reduce hexokinase activity in multiple cellular models^43^. Knockout of hepatic HKDC1 in pregnant mice has also been demonstrated to significantly impair glucose tolerance, highlighting the importance of liver HKDC1 on glucose metabolism during pregnancy^44^. Taken together, the evidence from our study and others suggests a more important role for *HKDC1* in glucose metabolism during pregnancy than outside of pregnancy, in addition to independent maternal and offspring effects on early growth, and highlights that while GDM shares many similarities with T2D, there are differences in at least one underlying pathway.

Finally, we used two-sample MR to investigate causal effects on GDM of 282 metabolic measures and risk factors available in the MR-Base GWAS catalogue (www.mrbase.org)^45^, including metabolites, anthropometric measures, hormones, immune system phenotypes, kidney traits and metals (**Supplementary Materials and Methods, Table S11**). We did not consider glycaemic traits (including HbA1c) because they are used to define GDM status. For each metabolic measure, we selected independent SNVs attaining genome-wide significance with the trait as instrumental variables. For each SNV, we extracted association summary statistics for GDM from the European ancestry-specific meta-analysis because we assessed independence of genetic instruments using LD from European ancestry haplotypes from the 1000 Genomes Project^24^. Of the 282 exposures considered, only BMI demonstrated significant evidence for a causal effect on GDM risk at a false discovery rate of 5% (**Table S11**). The estimated causal effect of higher BMI on higher GDM risk was directionally consistent across multiple MR models (**Figure 2**). The causal relationship of BMI with GDM is consistent with its effect on T2D^46^.

**Figure 2.**
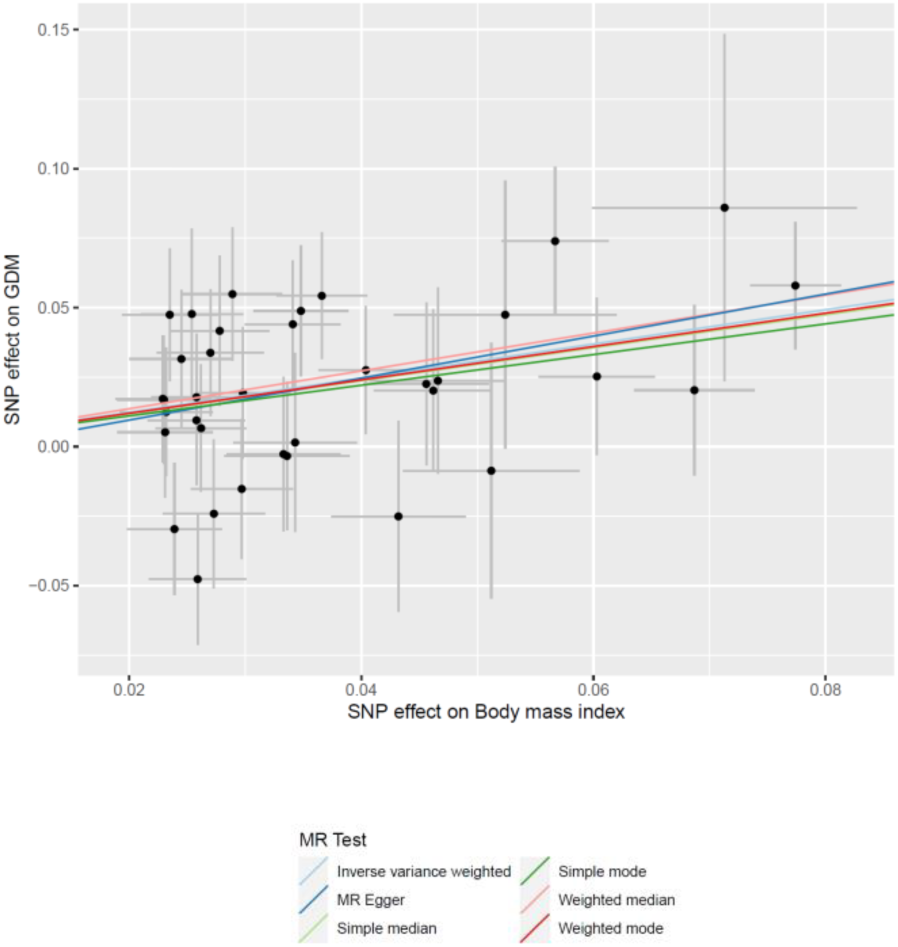
Effects of BMI on GDM from MR analyses. Each point corresponds to an independent SNV (genetic instrument), plotted according to the effect on BMI (on the x-axis) and the effect on GDM (log-OR, on the y-axis). Horizontal and vertical bars represent the standard errors of effect estimates. The coloured regression lines represent the effect of BMI on GDM from six MR models.

In conclusion, we have conducted the largest and most ancestrally diverse GWAS meta-analysis for GDM, where we identified associations mapping to *MTNR1B, TCF7L2, CDKAL1, CDKN2A-CDKN2B* and *HKDC1*. Our results demonstrated strong correlation in the effects of previously reported associations for T2D and those observed for GDM, and highlighted overlapping molecular mechanisms and tissues that mediate associations for both diseases. In contrast, variation at the *HKDC1* locus is not strongly associated with T2D, but instead plays a more important role in glucose metabolism during pregnancy than outside of pregnancy. The genetic diversity of GWAS contributing to our meta-analysis enabled identification of ancestry-correlated heterogeneity in allelic effects on GDM at two loci. Such heterogeneity could reflect variable impact of different pathophysiology driving glycaemic dysregulation in pregnancy between ancestries and emphasizes the need for increased sample sizes in under-represented population groups. In contrast, results were consistent between GWAS in which all women had a universal blood-based test and those that did not, suggesting little impact from misclassification due to selective use of diagnostic tests only in those deemed to be at high-risk. Finally, MR analyses revealed a significant causal effect of higher BMI on GDM risk, consistent with the causal association observed with T2D. Taken together, these results provide further support for the hypothesis that T2D and GDM are part of the same underlying pathology. However, they also highlight there are pathways to GDM that impact on glucose regulation only in pregnancy, and that additional GDM-specific associations will be revealed through GWAS in larger sample sizes.

## Data Availability

GWAS summary statistics will be made available once the manuscript has been accepted for publication.

## ACKNOWLEDGEMENTS

**ALSPAC**. Core funding for ALSPAC is provided by the UK Medical Research Council and Wellcome (217065/Z/19/) and the University of Bristol. Genotyping of the ALSPAC maternal samples was funded by Wellcome (WT088806) and the offspring samples were genotyped by Sample Logistics and Genotyping Facilities at the Wellcome Sanger Institute and LabCorp (Laboratory Corporation of America) using support from 23andMe. A comprehensive list of grants funding is available on the ALSPAC website (http://www.bristol.ac.uk/alspac/external/documents/grant-acknowledgements.pdf). We are extremely grateful to all the families who took part in ALSPAC, the midwives for their help in recruiting them, and the whole ALSPAC team, which includes interviewers, computer and laboratory technicians, clerical workers, research scientists, volunteers, managers, receptionists and nurses.

**AM**. We thank the participants of the ANDIS-MDC study. The study is supported by Swedish Research Council (project grant 521-2010-3490 and infrastructure grants 2010-5983, 2012-5538, and 2014-6395). The study is supported by Crafoord foundation (project grant 20200891) and Hjärt-Lungfonden (project grant: 20180522).

**BIB**. Born in Bradford (BiB) data used in this research was funded by Wellcome (WT101597MA), a joint grant from the UK Medical Research Council (MRC) and UK Economic and Social Science Research Council (ESRC) (MR/N024397/1), the British Heart Foundation (CS/16/4/32482) and the National Institute for Health Research (NIHR) under its Collaboration for Applied Health Research and Care (CLAHRC) for Yorkshire and Humber and the Clinical Research Network (CRN). Born in Bradford is only possible because of the enthusiasm and commitment of the Children and Parents in BiB. We are grateful to all the participants, teachers, school staff, health professionals and researchers who have made Born in Bradford happen.

**BOTNIA**. The Botnia Study (L.G., T.T.) have been financially supported by grants from Folkhälsan Research Foundation, the Sigrid Juselius Foundation, The Academy of Finland (grants no. 263401, 267882, 312063, 336822 to LG; 312072 and 336826 to TT), University of Helsinki, Nordic Center of Excellence in Disease Genetics, EU (EXGENESIS), MOSAIC FP7-600914, Ollqvist Foundation, Swedish Cultural Foundation in Finland, Finnish Diabetes Research Foundation, Foundation for Life and Health in Finland, Signe and Ane Gyllenberg Foundation, Finnish Medical Society, Paavo Nurmi Foundation, State Research Funding via the Helsinki University Hospital, Perklén Foundation, Närpes Health Care Foundation and Ahokas Foundation. The study has also been supported by the Ministry of Education in Finland, Municipal Heath Care Center and Hospital in Jakobstad and Health Care Centers in Vasa, Närpes and Korsholm.The research leading to these results has received funding from the European Research Council under the European Union’s Seventh Framework Programme (FP7/2007-2013) / ERC grant agreement n° 269045. The study is also supported by Crafoord foundation (project grant 20200891) and Hjärt-Lungfonden (project grant: 20180522) (RBP). The skillful assistance of the Botnia Study Group is gratefully acknowledged.

**DECODE**. We thank the women who have participated in the deCODE study.

**EGCUT**. This study was funded by the European Union through the European Regional Development Fund (Project No. 2014-2020.4.01.15-0012 and Project No. 2014-2020.4.01.16-0125), by the European Union through Horizon 2020 research and innovation programme under grant agreement No 101017802 and 810645 and by the Estonian Research Council grants PUT (PRG687, PRG1291). Data analyses were carried out in part in the High-Performance Computing Center of University of Tartu. We thank participants and support staff of Estonian Biobank.

**FINNGEDI**. The EPx-GDM study has been supported by the French government, managed by the National Research Agency (ANR) under the Future Investment Program (PIA) with the references ANR-10-LABX-0046 (European Genomic Institute for Diabetes [E.G.I.D]) and ANR-10-EQPX-07-01 (LIGAN-PM). This work was also supported by the ANR-16-CE17-0017-01, by the Fondation Francophone pour la Recherche sur le Diabète (FFRD) that is sponsored by the Fédération Française des Diabétiques (FFD), Abbott, AstraZeneca, Eli Lilly, Merck Sharp & Dohme (MSD) and Novo Nordisk, and by the National Center for Precision Diabetic Medicine – PreciDIAB, which is jointly supported by the French National Agency for Research (ANR-18-IBHU-0001), by the European Union (FEDER), by the Hauts-de-France Regional Council and by the European Metropolis of Lille (MEL). The FinnGeDi study was supported by: Academy of Finland, Diabetes Research Foundation, Foundation for Pediatric Research, Juho Vainio Foundation, Novo Nordisk Foundation, Signe and Ane Gyllenberg Foundation, Sigrid Jusélius Foundation, Yrjö Jahnsson Foundation for Pediatric Research, Finnish DiabetesMedical Foundation, Research Funds of Oulu University Hospital (state grants), Research Foundation, Novo Nordisk Foundation, funds of Helsinki University Hospital (state grants), Medical Research Center Oulu and National Institute for Health and Welfare (Finland).

**ERF**. We are grateful to all participants and their relatives, to general practitioners and neurologists for their contributions, to P. Veraart for her help in genealogy, to Jeannette Vergeer for the supervision of the laboratory work, and to P. Snijders for his help in data collection. The ERF study was supported by grants from the Netherlands Organisation for Scientific Research (Pionier, 047.016.009, 047.017.043), Erasmus MC, and the Centre for Medical Systems Biology (CMSB; National Genomics Initiative). Ayse Demrkan was supported by NWO (VENI-91616165), WCRF-2017/1641 and H2020-SC1-2019-874739.

**ESTGDM**. This study was supported by the Estonian Research Council grants IUT20-43 and PRG712. We are grateful to Dr Anne Kirss and midwife Mrs Laura Lauren from Women’s Clinic, Tartu University Hospital, for interviewing the participants and collecting the blood samples.

**GEN3G**. Gen3G has been supported by an American Diabetes Association accelerator award #1-15-ACE-26 (M.F.H), Fonds de Recherche du Québec en Santé #20697 (M.F.H); Canadian Institute of Health Research #MOP 115071 (M.F.H), and Diabète Québec grants (P.P. and L.B.). Gen3G research team thanks all Gen3G participants.

**GIFTS**. This study was supported by Medical Research Council [Clinical Research Training Fellowship (G0800441)], the European Union (FP7 EU grant: 83599025) and the Diabetes Association of Bangladesh. The authors are indebted to all the participants from Dhaka (Bangladesh) and London (UK).

**HONGKONG**. This study was supported by National Institutes of Health (NIH) grants HD-34242, HD-34243, HG-004415, and CA-141688, Institutes of Health Research–INMD (Funding Reference Number 110791), and by the American Diabetes Association. The authors are indebted to the participants of the HAPO Study at Hong Kong.

**NFBC Cohort 1966 31y follow-up study**. We thank all cohort members and researchers who participated in the 31 yrs study. We also wish acknowledge the work of the NFBC project center. NFBC1966 received financial support from University of Oulu Grant no. 65354 and 24000692, Oulu University Hospital Grant no. 2/97, 8/97, and 24301140, Ministry of Health and Social Affairs Grant no. 23/251/97, 160/97, 190/97. National Institute for Health and Welfare, Helsinki Grant no. 54121, Regional Institute of Occupational Health, Oulu, Finland Grant no. 50621, 54231, 54121, Regional Institute of Occupational Health, Oulu, Finland Grant no. 50621, 54231 and ERDF European Regional Development Fund Grant no. 539/2010 A31592. The NFBC team is acknowledging the following Funding/Support: This work was supported by the European Union’s Horizon 2020 research and innovation program under grant agreement No. 633595 (DynaHEALTH), and grant agreement No.733206 (LifeCycle) H2020-824989 EUCANCONNECT, H2020-873749 LongITools, H2020-848158 EarlyCause and the JPI HDHL, PREcisE project MR/S03658X/1, ZonMw the Netherlands no. P75416; the academy of Finland EGEA-project (285547).

**NFBC Cohort 1966 46y follow-up study**. We thank all cohort members and researchers who participated in the 46 yrs study. We also wish acknowledge the work of the NFBC project center. NFBC1966 received financial support from University of Oulu Grant no. 24000692, Oulu University Hospital Grant no. 24301140, ERDF European Regional Development Fund Grant no. 539/2010 A31592.

**NFBC Cohort 1986**. We thank all cohort members and researchers who have participated in the study. We also wish acknowledge the work of the NFBC project center. NFBC1986 received financial support: EU QLG1-CT-2000-01643 (EUROBLCS) Grant no. E51560, NorFA Grant no. 731, 20056, 30167, USA / NIH 2000 G DF682 Grant no. 50945.. The NFBC team acknowledges the following funding/support: the European Union’s Horizon 2020 research and innovation program under grant agreement No. 633595 (DynaHEALTH), and grant agreement No.733206 (LifeCycle) H2020-824989 EUCANCONNECT, H2020-873749 LongITools, H2020-848158 EarlyCause and the JPI HDHL, PREcisE project, ZonMw the Netherlands no. P75416; the academy of Finland EGEA-project (285547).

**PREDO**. The PREDO study was supported by Academy of Finland, EVO research funding (A special Finnish state subsidy for health science research), Finnish Medical Foundation, Jane and Aatos Erkko Foundation, Päivikki and Sakari Sohlberg Foundation, University of Helsinki Research Funding, Jalmari ja Rauha Ahokas foundation, Yrjö Jahnsson foundation, Juho Vainio foundation.

**SNUH**. This work was supported by the Korea Health 21 R&D Project, Korean Ministry of Health and Welfare (grant no. 00-PJ3-PG6-GN07-001). This study was supported by a grant from the Korea Health Technology R&D Project through the Korea Health Industry Development Institute, funded by the Ministry of Health & Welfare, Republic of Korea to S.H.K (grant number HI15C3131).

**STORK**. The STORK study received additional funding from the Norwegian Diabetes Association, the Norwegian Odd Fellow Research Fund and Johan Selmer Kvanes’ Endowment for Research in Diabetes.

**STORKG**. Acknowledge Hormone Laboratory, Oslo University Hospital for DNA-extraction.

**UKBB**. UK Biobank analyses were conducted using the UK Biobank resource under application 11867.

**VIVA**. Grants from the US National Institutes of Health (R01 HD034568, UH3 OD023286). Project Viva is thankful to all Project Viva participants for their participation to research over many years.

**Individual author acknowledgements**. D.A.L. is supported by the European Research Council (669545), US National Institute for Health (R01 DK10324), and British Heart Foundation (CH/F/20/90003). D.A.L. and M.C.B. work in a Unit supported by the University of Bristol and the UK Medical Research Council (MC_UU_00011/6) and are supported by a British Heart Foundation Accelerator Award (AA/18/7/34219). R.M.F. and R.N.B. were supported by Sir Henry Dale Fellowship (Wellcome and Royal Society grant: WT104150). R.M.F. is funded by a Wellcome Senior Research Fellowship (WT220390). G.H.M. has received funding from the South-Eastern Health Authority of Norway, the Norwegian Diabetes Association and Nils Normans Minnegave. G.H.M. is supported by the Norwegian Research Council (Post doctorial mobility research grant 287198).

M.C.B. was funded by a Medical Research Council (MRC) Skills Development Fellowship [MR/P014054/1] and a University of Bristol Vice-Chancellor’s Fellowship.

The authors acknowledge the use of the University of Exeter High-Performance Computing (HPC) facility in carrying out this work.

Funders and others acknowledged here had no influence on the study design, data collection or interpretation of results. Views expressed are those of the authors and not necessarily any funder or other acknowledged here.

This research was funded in part, by Wellcome grants WT104150 and WT220390. A CC BY or equivalent licence is applied to the author accepted manuscript arising from this submission, in accordance with the grant’s open access conditions.

## AUTHOR CONTRIBUTIONS

**Central analysis group:** N.P., G-H..M., M.-C.B., T.F., J.P.C., R.M.F., D.A.L., A.P.M., R.M. **Cohort analysis group:** G.M., M.B., T.F., C.A., R.N.B., M.C., G.H., A.Heiskala, A.Joensuu, V.K., S.K., F.T.J.L., J.Liu, S.R.-S., G.T., S.H., R.B.P. **Cohort sample collection, phenotyping, genotyping and additional analysis:** T.A., J.A., B.B., A.B., F.D., A.D., P.F., K.H., H.H., S.H., A.Hussain, E.Kajantie, E.Keikkala, A.K., J.Lahti, T.L., S.M., C.S., A.T., E.T., R.U., M.V., P.M.V., R.B.P., R.M.F. **Cohort PI:** K.I.B., L.B., C.M.D., S.F., L.G., E.H., G.M.H., G.A.H., H.C.J., M.J., A.Jenum, H.L., O.M., E.O., K.P., P.P., R.B.P., E.Q., S.S., K.S., V.S., T.T., M.-F.H., P.W.F., M.I.M, C.M.L., D.A.L., A.P.M., R.M. **Writing group:** G.-H.M., M.-C.B., C.M.L., R.M.F., D.A.L., A.P.M., R.M.

## REFERENCES

1. Metzger, B. E. Hyperglycaemia and adverse pregnancy outcome (HAPO) study: Associations with maternal body mass index. BJOG An Int. J. Obstet. Gynaecol. 117, 575–584 (2010).

2. Farrar, D. et al. Association between hyperglycaemia and adverse perinatal outcomes in south Asian and white British women: Analysis of data from the Born in Bradford cohort. Lancet Diabetes Endocrinol. 3, 795–804 (2015).

3. Farrar, D. et al. Hyperglycaemia and risk of adverse perinatal outcomes: Systematic review and meta-analysis. BMJ (Online) vol. 354 (2016).

4. Stacey, T. et al. Gestational diabetes and the risk of late stillbirth: a case–control study from England, UK. BJOG An Int. J. Obstet. Gynaecol. 126, 973–982 (2019).

5. Kwak, S. H. et al. Subsequent pregnancy after gestational diabetes mellitus. Diabetes Care 31, 1867–1871 (2008).

6. Kramer, C. K., Campbell, S. & Retnakaran, R. Gestational diabetes and the risk of cardiovascular disease in women: a systematic review and meta-analysis. Diabetologia vol. 62 905–914 (2019).

7. Vounzoulaki, E. et al. Progression to type 2 diabetes in women with a known history of gestational diabetes: Systematic review and meta-analysis. BMJ 369, (2020).

8. Farrar, D. Hyperglycemia in pregnancy: Prevalence, impact, and management challenges. Int. J. Womens. Health 8, 519–527 (2016).

9. American Diabetes Association. Gestational Diabetes Mellitus. Diabetes Care vol. 27 s88–s90 (2004).

10. Zhang, C. & Ning, Y. Effect of dietary and lifestyle factors on the risk of gestational diabetes: Review of epidemiologic evidence. in American Journal of Clinical Nutrition vol. 94 (Am J Clin Nutr, 2011).

11. Jang, H. C., Min, H. K., Lee, H. K., Cho, N. H. & Metzger, B. E. Short stature in Korean women: A contribution to the multifactorial predisposition to gestational diabetes mellitus. Diabetologia 41, 778–783 (1998).

12. Sattar, N. & Greer, I. A. Pregnancy complications and maternal cardiovascular risk: Opportunities for intervention and screening? British Medical Journal vol. 325 157–160 (2002).

13. Rich-Edwards, J. W., Fraser, A., Lawlor, D. A. & Catov, J. M. Pregnancy characteristics and women’s future cardiovascular health: An underused opportunity to improve women’s health? Epidemiol. Rev. 36, 57–70 (2014).

14. Vujkovic, M. et al. Discovery of 318 new risk loci for type 2 diabetes and related vascular outcomes among 1.4 million participants in a multi-ancestry meta-analysis. Nat. Genet. 52, 680–691 (2020).

15. Spracklen, C. N. et al. Identification of type 2 diabetes loci in 433,540 East Asian individuals. Nature 582, 240–245 (2020).

16. Mahajan, A. et al. Trans-ancestry genetic study of type 2 diabetes highlights the power of diverse populations for discovery and translation. medRxiv (2020).

17. Mahajan, A. et al. Fine-mapping type 2 diabetes loci to single-variant resolution using high-density imputation and islet-specific epigenome maps. Nat. Genet. 50, 1505–1513 (2018).

18. Watanabe, R. M. Inherited destiny? Genetics and gestational diabetes mellitus. Genome Medicine vol. 3 (2011).

19. Powe, C. E. & Kwak, S. H. Genetic Studies of Gestational Diabetes and Glucose Metabolism in Pregnancy. Current Diabetes Reports vol. 20 (2020).

20. Zhang, C. et al. Genetic variants and the risk of gestational diabetes mellitus: A systematic review. Hum. Reprod. Update 19, 376–390 (2013).

21. Kawai, V. K. et al. A genetic risk score that includes common type 2 diabetes risk variants is associated with gestational diabetes. Clin. Endocrinol. (Oxf). 87, 149–155 (2017).

22. Powe, C. E. et al. Genetic determinants of glycemic traits and the risk of gestational diabetes mellitus. Diabetes 67, 2703–2709 (2018).

23. Kwak, S. H. et al. A genome-wide association study of gestational diabetes mellitus in Korean women. Diabetes 61, 531–541 (2012).

24. Auton, A. et al. A global reference for human genetic variation. Nature vol. 526 68–74 (2015).

25. McCarthy, S. et al. A reference panel of 64,976 haplotypes for genotype imputation. Nat. Genet. 48, 1279–1283 (2016).

26. Gurdasani, D., Barroso, I., Zeggini, E. & Sandhu, M. S. Genomics of disease risk in globally diverse populations. Nature Reviews Genetics vol. 20 520–535 (2019).

27. Mägi, R. et al. Trans-ethnic meta-regression of genome-wide association studies accounting for ancestry increases power for discovery and improves fine-mapping resolution. Hum. Mol. Genet. 26, 3639–3650 (2017).

28. Scott, R. A. et al. Large-scale association analyses identify new loci influencing glycemic traits and provide insight into the underlying biological pathways. Nat. Genet. 44, 991–1005 (2012).

29. Chen, J. et al. The trans-ancestral genomic architecture of glycemic traits. Nat. Genet. 53, 840–860 (2021).

30. Hayes, M. G. et al. Identification of HKDC1 and BACE2 as genes influencing glycemic traits during pregnancy through genome-wide association studies. Diabetes 62, 3282–3291 (2013).

31. Beaumont, R. N. et al. Genome-wide association study of offspring birth weight in 86 577 women identifies five novel loci and highlights maternal genetic effects that are independent of fetal genetics. Hum. Mol. Genet. 27, 742–756 (2018).

32. Chen, J. et al. The Trans-Ancestral Genomic Architecture of Glycaemic Traits. Astrid van Hylckama Vlieg 14, 203 (2020).

33. Morris, A. P. et al. Large-scale association analysis provides insights into the genetic architecture and pathophysiology of type 2 diabetes. Nat. Genet. 44, 981–990 (2012).

34. Harrow, J. et al. GENCODE: The reference human genome annotation for the ENCODE project. Genome Res. 22, 1760–1774 (2012).

35. Dunham, I. et al. An integrated encyclopedia of DNA elements in the human genome. Nature 489, 57–74 (2012).

36. Pasquali, L. et al. Pancreatic islet enhancer clusters enriched in type 2 diabetes risk-associated variants. Nat. Genet. 46, 136–143 (2014).

37. Varshney, A. et al. Genetic regulatory signatures underlying islet gene expression and type 2 diabetes. Proc. Natl. Acad. Sci. U. S. A. 114, 2301–2306 (2017).

38. Gaulton, K. J. et al. Genetic fine mapping and genomic annotation defines causal mechanisms at type 2 diabetes susceptibility loci. Nat. Genet. 47, 1415–1425 (2015).

39. Williams, K. et al. Skeletal muscle enhancer interactions identify genes controlling whole-body metabolism. Nat. Commun. 11, (2020).

40. Tyrrell, J. S., Yaghootkar, H., Freathy, R. M., Hattersley, A. T. & Frayling, T. M. Parental diabetes and birthweight in 236 030 individuals in the UK Biobank study. Int. J. Epidemiol. 42, 1714–1723 (2013).

41. Warrington, N. M. et al. Maternal and fetal genetic effects on birth weight and their relevance to cardio-metabolic risk factors. Nat. Genet. 51, 804–814 (2019).

42. GTEx Consortium. The GTEx Consortium atlas of genetic regulatory effects across human tissues. Science 369, 1318–1330 (2020).

43. Guo, C. et al. Coordinated regulatory variation associated with gestational hyperglycaemia regulates expression of the novel hexokinase HKDC1. Nat. Commun. 6, (2015).

44. Khan, M. W., Priyadarshini, M., Cordoba-Chacon, J., Becker, T. C. & Layden, B. T. Hepatic hexokinase domain containing 1 (HKDC1) improves whole body glucose tolerance and insulin sensitivity in pregnant mice. Biochim. Biophys. Acta - Mol. Basis Dis. 1865, 678–687 (2019).

45. Hemani, G. et al. The MR-base platform supports systematic causal inference across the human phenome. Elife 7, (2018).

46. Yuan, S. & Larsson, S. C. An atlas on risk factors for type 2 diabetes: a wide-angled Mendelian randomisation study. Diabetologia 63, 2359–2371 (2020).

