## Supplementory Figures for "Trans-ancestry genome-wide association study of gestational diabetes mellitus highlights genetic links with type 2 diabetes"

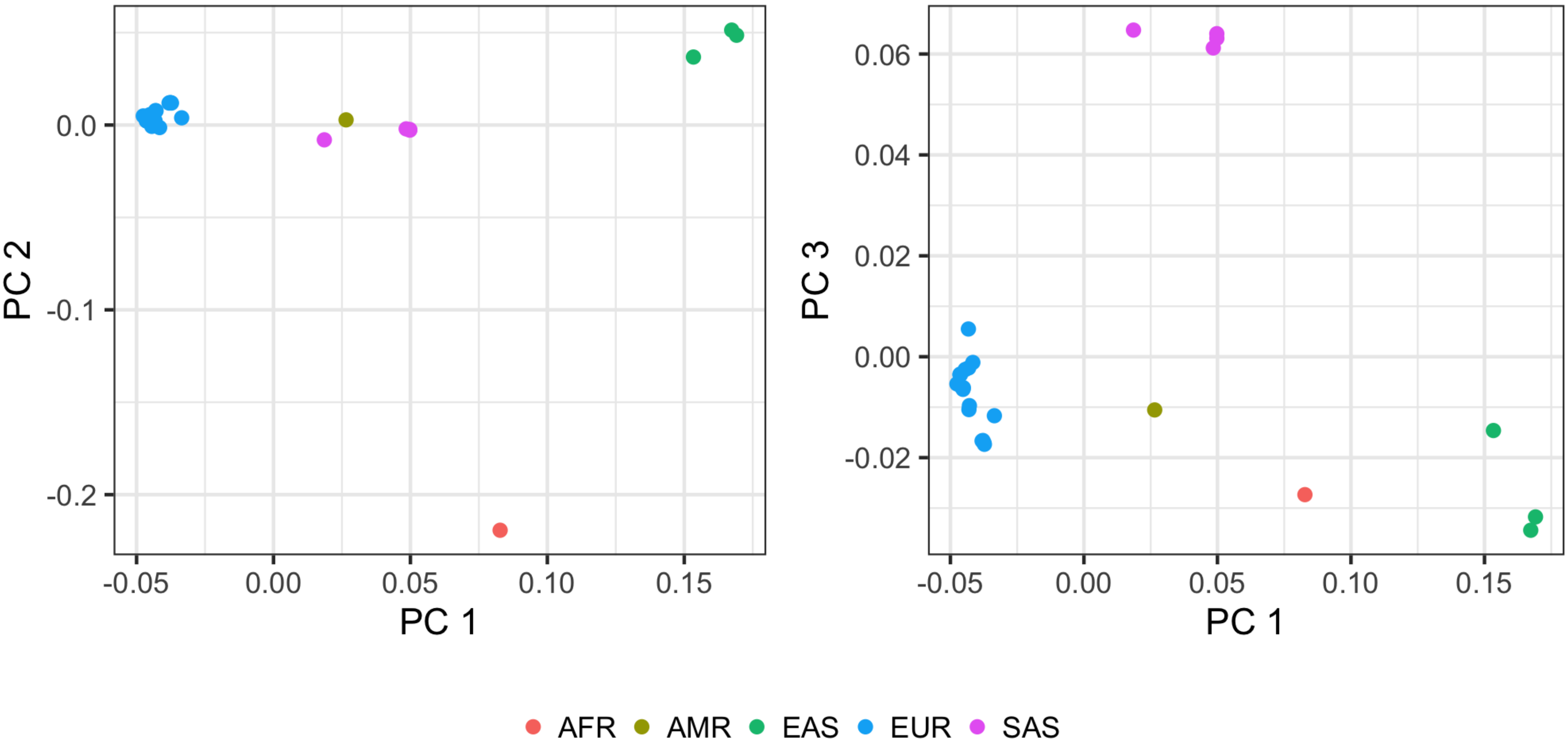


**Figure S1. Axes of genetic variation separating GWAS of GDM across diverse populations.** The first three axes of genetic variation (PC 1, PC 2 and PC 3) from multi-dimensional scaling of the Euclidean distance matrix between populations are sufficient to separate five ancestry groups: African (AFR), American (AMR), East Asian (EAS), European (EUR), and South Asian (SAS).

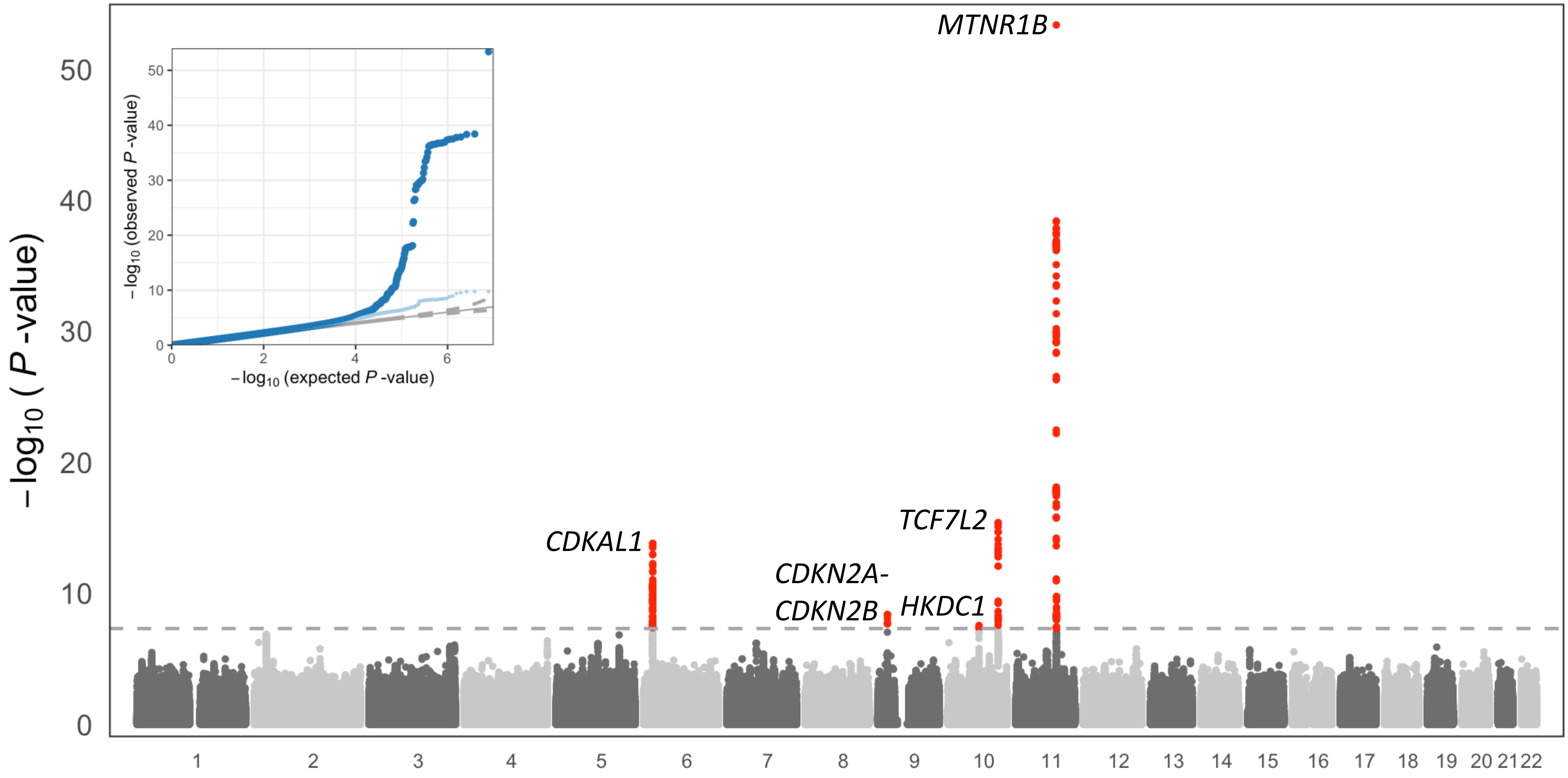


**Figure S2. Manhattan plot of genome-wide GDM association from trans-ancestry meta-regression (MR-MEGA) of 5,485 cases and 347,856 controls.** Each point represents a SNV passing quality control in the trans-ancestry meta-regression, plotted with their association *p*-value (on a $-\log_{10}$ scale) as a function of genomic position (NCBI build 37). Association signals attaining genome-wide significance are highlighted in red ($p<5\times10^{-8}$). In the quantile-quantile plot, each point represents a SNV, plotted with their observed and expected association *p*-values (on a $-\log_{10}$ scale). The plot in dark blue corresponds to the distribution including all SNVs, whilst the plot in light blue corresponds to the distribution after excluding the loci attaining genome-wide significance.

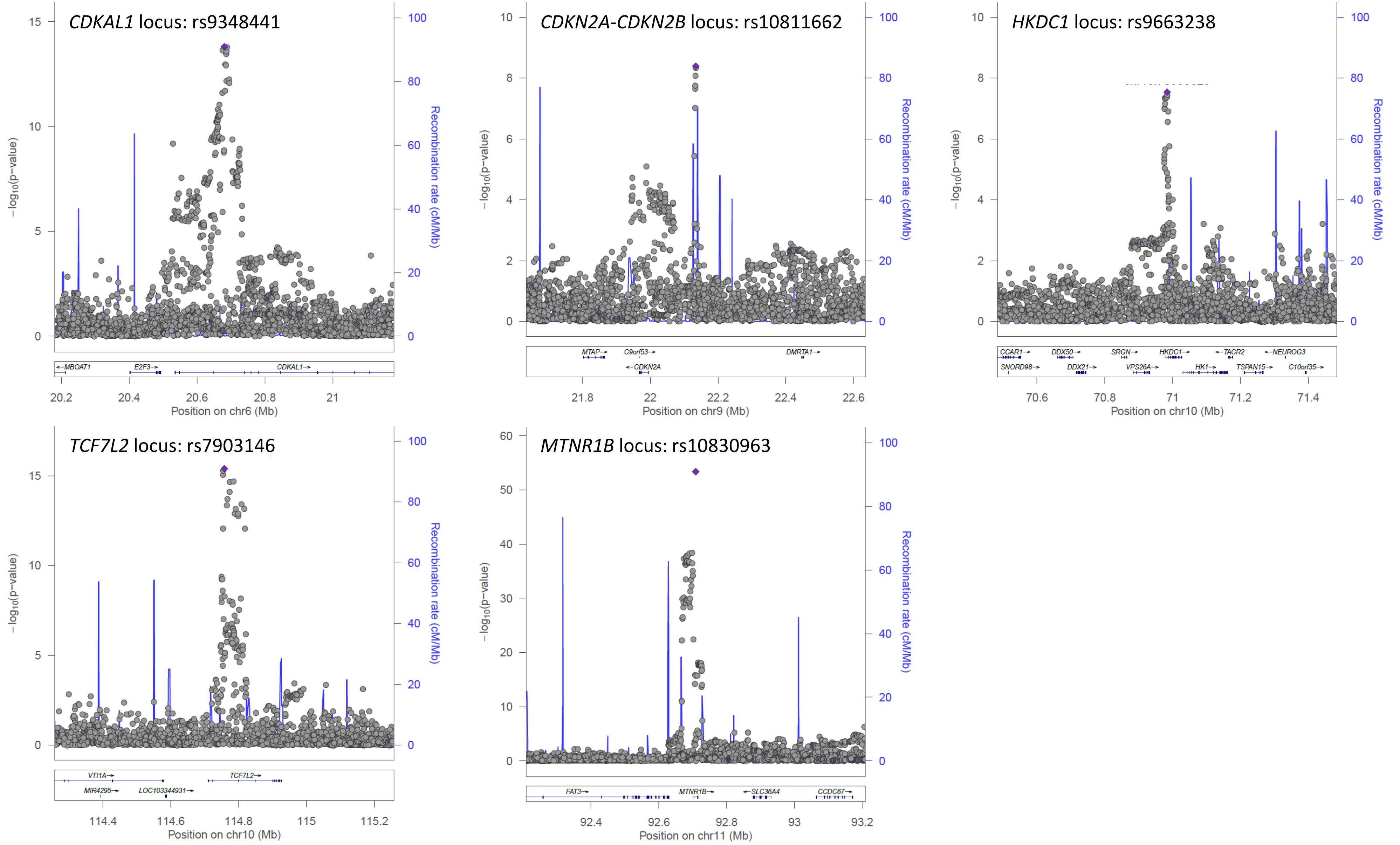


**Figure S3. Signal plots for association signals with GDM at loci attaining genome-wide significance ($p<5\times10^{-8}$) in trans-ancestry meta-regression (MR-MEGA) of up to 5,485 cases and 347,856 controls.** Each point represents a SNV passing quality control in the trans-ancestry meta-regression, plotted with their conditional *p*-value (on a $-\log_{10}$ scale) as a function of genomic position (NCBI build 37). The lead SNP is highlighted with the purple diamond. Gene annotations are taken from the University of California Santa Cruz genome browser. Recombination rates are estimated from the Phase II HapMap.

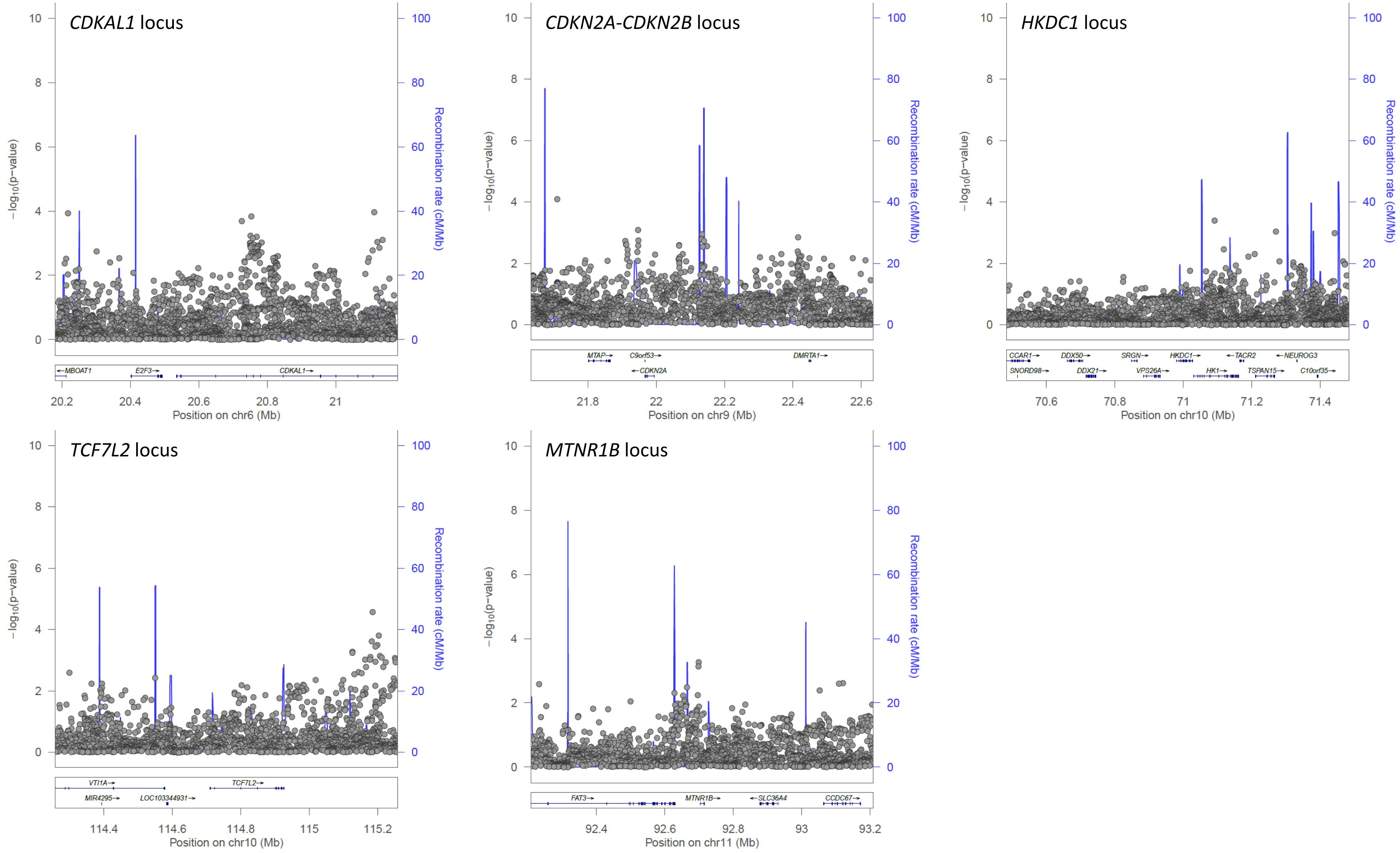


**Figure S4. Signal plots for residual association signals with GDM after adjusting for the lead SNV at loci in trans-ancestry meta-regression (MR-MEGA) of up to 5,485 cases and 347,856 controls.** Each point represents a SNV passing quality control in the trans-ancestry meta-regression, plotted with their conditional *p*-value (on a $-\log_{10}$ scale) as a function of genomic position (NCBI build 37). Gene annotations are taken from the University of California Santa Cruz genome browser. Recombination rates are estimated from the Phase II HapMap.

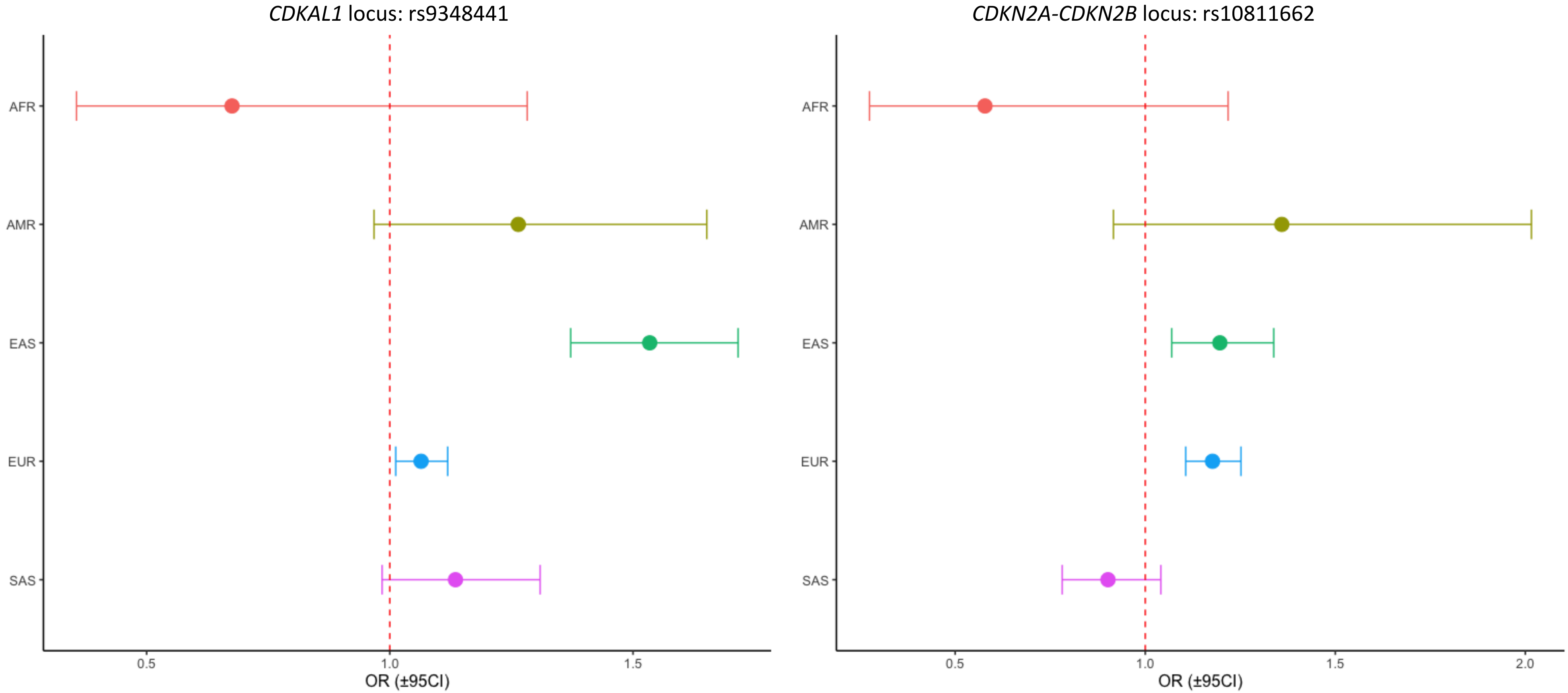


**Figure S5. Forest plot presenting ancestry-specific allelic effects on GDM of the risk allele for lead SNPs at the *CDKAL1* locus and *CDKN2A-CDKN2B* locus.** The plot presents the odds ratio (OR) for the risk allele, and corresponding 95% confidence interval (CI), from ancestry-specific fixed-effects meta-analysis: African (AFR); American (AMR); East Asian (EAS); European (EUR); and South Asian (SAS).
