## Supplementary Materials and Methods for "Trans-ancestry genome-wide association study of gestational diabetes mellitus highlights genetic links with type 2 diabetes"

**Ethics statement.** All human research was approved by the relevant institutional review boards and conducted according to the Declaration of Helsinki. All participants provided written informed consent.

**Study-level analyses.** Individuals were assayed with a range of GWAS genotyping arrays, with sample and SNV quality control undertaken within each study (**Tables S2 and S3**). Samples were pre-phased and imputed up to reference panels from the 1000 Genomes Project (phase 1, March 2012 release; phase 3, October 2014 release)[1,2], Haplotype Reference Consortium[3], or population-specific whole-genome sequencing[4–6] (Table S3). SNVs with poor imputation quality ($r^2$<0.3 or info<0.4) and/or minor allele count <5 were excluded from downstream association analyses (**Table S3**). Association with GDM was evaluated in a regression framework, under an additive model in the dosage of the minor allele, with adjustment for principal components and other study-specific covariates to minimize the population stratification effects (**Table S3**). Phenotype definition and covariate adjustments were not harmonised between GWAS because of differences in individual study design and availability of non-genetic risk factor information. Analyses accounted for structure (population stratification and/or familial relationships) by: (i) excluding related samples and adjustment for principal components derived from a genetic relatedness matrix (GRM) as additional covariates in the regression model; or (ii) incorporating a random effect for the GRM in a mixed model (**Table S3**). Allelic effects and corresponding standard errors that were estimated from a linear (mixed) model were converted to the log-odds scale[7]. Study-level association summary statistics (p-values and standard error of allelic effects) were corrected for residual structure by means of genomic control[8] if the inflation factor was >1 (**Table S3**).

**Trans-ancestry meta-analyses.** To account for the different reference panels used for imputation across GWAS, we restricted our analyses to autosomal bi-allelic SNVs from the 1000 Genomes Project reference panel (phase 3, October 2014 release)[1] that are also present in the Haplotype Reference Consortium reference panel[3]. We considered only those SNVs with MAF >0.5% in haplotypes in at least one of the five ancestry groups represented in the 1000 Genomes Project (phase 3, October 2014 release).

Our primary trans-ancestry analysis utilised meta-regression, implemented in the MR-MEGA software, which allows for allelic effect heterogeneity between GWAS that is correlated with ancestry[9]. We first constructed a distance matrix of mean effect allele frequency differences between each pair of GWAS across a subset of SNVs reported in all studies. We implemented multi-dimensional scaling of the distance matrix to obtain three principal components that defined axes of genetic variation to separate GWAS from the five ancestry groups (**Figure S1**). For each SNV, we then modelled allelic log-ORs across GWAS in a linear regression framework, weighted by the inverse of the variance of the effect estimates, incorporating the three axes of genetic variation as covariates. Under this model, we tested for association with GDM allowing for allelic effect heterogeneity between GWAS that is correlated with ancestry. We corrected the meta-regression association *p*-values for inflation due to residual structure between GWAS using genomic control adjustment. We considered only those SNVs reported ≥50% of the total effective sample size in downstream analyses.

For each SNV, we also conducted fixed-effects meta-analysis across GWAS under an inverse-variance weighting of allelic log-ORs using GWAMA[10]. We corrected standard errors of the resulting effect estimates for inflation due to residual structure between GWAS by genomic control adjustment.

**Defining GDM loci.** We identified lead SNVs attaining genome-wide significant evidence of association ($p<5\times10^{-8}$) in the trans-ancestry meta-regression that were separated by at least 500kb. Loci were defined by the genomic interval mapping 500kb up- and downstream of each lead SNV.

**Assessing evidence for multiple distinct association signals at GDM loci.** Each GWAS was first assigned to one of the ancestry groups (**Table S2**) represented in the 1000 Genomes Project reference panel (phase 3, October 2014 release)[1]. Haplotypes in the panel that were specific to that ancestry group were used as a reference for LD between SNVs across loci for the GWAS in approximate conditional analyses implemented in GCTA[11]. For each locus, we applied GCTA to each GWAS to condition on the lead SNV at the locus, using the study-level association summary statistics and matched LD reference. Allelic log-ORs from the approximate conditional analyses across GWAS were modelled in the trans-ancestry meta-regression framework implemented in MR-MEGA[9], incorporating the three axes of genetic variation as covariates, and weighted by the inverse of the variance of the effect estimates. The meta-regression association *p*-values were corrected for inflation due to residual structure between GWAS by using the same genomic control adjustment as in the unconditional analysis. If no SNVs attained genome-wide significant ($p<5\times10^{-8}$) evidence of residual GDM association in the meta-regression, we concluded that there was a single association signal at the locus.

**Ancestry-specific meta-analyses.** We aggregated association summary statistics across GWAS from the same ancestry group via fixed-effects meta-analysis based on inverse-variance weighting of allelic log-OR to obtain effect size estimates using GWAMA[10]. We corrected association *p*-values and standard errors of allelic effects from each ancestry group for residual inflation due to structure between GWAS by genomic control adjustment if the inflation factor was >1. We estimated the mean effect allele frequency across GWAS from each ancestry group, weighted by the effective sample size of the study.

**Investigating the source of heterogeneity in allelic effects on GDM.** We extended the meta-regression model implemented in the MR-MEGA software to investigate the impact of ancestry and the use of a universal blood-based test to define GDM status on heterogeneity in allelic effects on GDM at lead SNVs. We modelled allelic log-ORs across GWAS in a linear regression framework, weighted by the inverse of the variance of the effect estimates, incorporating a covariate indicating whether GDM status was defined by a universal blood-based test (**Table S1**) in addition to the three axes of genetic variation.

**Genetic risk score of T2D on GDM.** We considered lead SNVs at 237 previously reported loci for T2D from the DIAMANTE Consortium[12] obtained from a trans-ancestry meta-analysis of 180,834 cases and 1,159,055 controls (48.9% non-European ancestry). For each of the 222 SNVs that were reported in our trans-ancestry meta-analysis, we compared association summary statistics (risk allele, other allele, log-OR and *p*-value) for GDM and those reported for T2D. We excluded lead SNVs for T2D that also attained genome-wide significance for GDM. For the remaining SNVs, we regressed the log-ORs for GDM (weighted by their corresponding variances) on the log-OR for T2D, as implemented in grs.summary function[13] of the gtx package in R version 3.4.2. We estimated the percentage of GDM variance explained, as measured by pseudo $R^2$.

**Genetic correlation between GDM and glycaemic traits.** We used LD Hub[14] to perform LD score regression[15] of the European ancestry association summary statistics for GDM on other glycaemic traits. We included T2D[16], fasting glucose[17], fasting insulin[17], fasting proinsulin[17], glucose 2 h post oral glucose tolerance test (adjusted for BMI)[18], HbA1c[19], HOMA-B[20] and HOMA-IR[20]. European ancestry association summary statistics for GDM were filtered so that only SNVs with minor allele frequency > 0.01 was included before performing the LD score regression. Genetic correlations between the different glycaemic traits were obtained from the LD Hub lookup centre. Visualisation was performed using the R package ggplot2[21] in R version 3.6.1.

**Enrichment of GDM association signals in genomic annotations.** We mapped each SNV across the genome to three categories of functional and regulatory annotations. First, we considered genic regions, as defined by the GENCODE Project[22], including protein-coding exons, and 3' and 5' UTRs as different annotations. Second, we considered chromatin immuno-precipitation sequence (ChIP-seq) binding sites for 165 transcription factors: 161 proteins from the ENCODE Project[23] and four additional factors assayed in primary pancreatic islets[24]. Third, we considered 13 unique and recurrent chromatin states, including promoter, enhancer, transcribed, and repressed regions, in four diabetes-relevant tissues[25]: pancreatic islets, liver, adipose, and skeletal muscle. This resulted in a total of 220 genomic annotations for enrichment analyses.

We tested for genome-wide enrichment of GDM associations that map to genomic annotations using fGWAS[26]. To do this, we approximated the Bayes' factor in favour of GDM association for the $j$th SNV by

$$\Lambda_j = \exp\left[\frac{D_j \; -4\ln K_j}{2}\right],$$

where $D_j$ is the deviance across $K_j$ contributing GWAS contributing to the trans-ancestry meta-regression[9]. We first considered each annotation separately and identified those with significant enrichment ($p$<0.05). We then used an iterative approach to identify a joint model of enriched annotations from this set. At each iteration, we dropped the annotation from the joint model that minimised the reduction in the penalised likelihood. We continued until no additional annotations worsened the fit of the joint model at nominal significance ($p$<0.05). We next used the cross-validation likelihood because the significance of parameter estimates from the penalised likelihood cannot be assessed using standard statistical approaches. For the selected joint model, we identified the penalty that maximised the cross-validation likelihood. Finally, we dropped any annotations from the joint model that resulted in a decrease in the cross-validation likelihood.

**Annotation informed fine-mapping of the *HKDC1* locus.** At the *HKDC1* locus, we calculated the posterior probability of driving the GDM association for each SNV under an annotation-informed prior model derived from the globally enriched functional and regulatory annotations identified by fGWAS. Specifically, for the $j$th SNV at the locus, the posterior probability $\pi_j \propto \gamma_j \Lambda_j$, where $\Lambda_j$ is the Bayes' factor in favour of GDM association from the meta-regression, derived above. In this expression, the relative annotation-informed prior for the SNV is given by

$$\gamma_j = \exp\left[\sum_k \hat{\beta}_k z_{jk}\right],$$

where the summation is over the enriched annotations, $\hat{\beta}_k$ is the estimated log-fold enrichment of the $k$th annotation from the final joint model, and $z_{jk}$ is an indicator variable taking the value 1 if the $j$th SNV maps to the $k$th annotation, and 0 otherwise. We derived a 99% credible set[27] for the locus by: (i) ranking all SNVs according to their posterior probability $\pi_j$; and (ii) including ranked SNVs until their cumulative posterior probability attained or exceeded 0.99.

We conducted a look-up of 99% credible set variants at the *HKDC1* locus for significant ($q$<0.05) *cis*-expression quantitative trait loci (eQTLs) across tissues in the GTEx Project[28]. We reported only those 99% credible variants that were the lead SNV for the eQTL signal.

**MR assessment of the effects of metabolic traits on GDM risk.** We systematically searched the MR-Base GWAS catalogue (https://www.mrbase.org) for metabolic measures. We selected all subcategories of metabolites, which included "amino acid", "carbohydrate", "cofactors and vitamins", "energy", "fatty acid", "keto acid", "lipid", "metabolite salt", "metabolites ratio", "NA", "nucleotide", "peptide", "protein", "unknown metabolite" and "xenobiotics". We also selected the following subcategories of risk factors: "anthropometric", "hormone", "immune system", "kidney" and "metal". We identified European ancestry GWAS in MR-Base for each selected metabolic trait. Where more than one GWAS was available for a trait, we gave preference to: women-specific studies with the largest sample sizes and numbers of SNVs. Any GWAS undertaken only in men were excluded.

For each metabolic trait with more than five genetic instruments, we conducted MR analyses using a "mixture of experts" (MoE) machine learning approach[29]. This approach maximises statistical power whilst minimising the impact of horizontal pleiotropy by combining four instrument selection approaches to 14 different MR models. The four approaches for selecting genetic instruments using MoE were: (i) "top hits" corresponding to independent variants associated at genome-wide significance ($p$<5x10$^{-8}$, $r^2$<0.001 using 1000G CEU as the reference population); (ii) "directional filtration" that removed instruments from "top hits" that are likely to be related to the outcome through reverse causation using Steiger filtering[30]; (iii) "heterogeneity filtering" that removed instruments from "top hits" that make a substantial contribution to Cochran's Q statistic ($p$<0.05); and (iv) combined "directional filtration" and "heterogeneity filtering". The 14 MR models were: seven mean-based methods (inverse variance weighting with fixed effects, IVW random effects, MR-Egger fixed effects, MR-Egger random effects, and the three Rucker estimates), three median-based methods (simple, weighted and penalised median estimator), and four mode-based methods (simple and weighted mode, each weighted with or without the assumption of no measurement error in the exposure estimates). The best combination of instrument selection-MR method was identified using a variable predicted by MoE, scaled between 0 and 1, where 1 indicates best performance.

For metabolic traits with five or fewer genetic instruments, the MoE approach could not be applied because many of the MR models require larger numbers of SNVs. For these metabolic traits, we used either the Wald ratio estimate (one SNV) or the inverse-variance weighted estimate (between two and five SNVs).

All analyses were conducted in R version 3.6 using the packages “TwoSampleMR” (version 0.5.4) and “MRInstruments” for the MR analyses and “EpiCircos” (https://github.com/mattlee821/EpiCircos).

### References


1. Auton, A. *et al.* A global reference for human genetic variation. *Nature* **526**, 68–74 (2015).
2. Altshuler, D. M. *et al.* An integrated map of genetic variation from 1,092 human genomes. *Nature* **491**, 56–65 (2012).
3. McCarthy, S. *et al.* A reference panel of 64,976 haplotypes for genotype imputation. *Nat. Genet.* **48**, 1279–1283 (2016).
4. Mitt, M. *et al.* Improved imputation accuracy of rare and low-frequency variants using population-specific high-coverage WGS-based imputation reference panel. *Eur. J. Hum. Genet.* **25**, 869–876 (2017).
5. Gudbjartsson, D. F. *et al.* Large-scale whole-genome sequencing of the Icelandic population. *Nat. Genet.* **47**, 435–444 (2015).
6. Surakka, I. *et al.* The rate of false polymorphisms introduced when imputing genotypes from global imputation panels. *bioRxiv* 080770 (2016). doi:10.1101/080770
7. Cook, J. P., Mahajan, A. & Morris, A. P. Guidance for the utility of linear models in meta-analysis of genetic association studies of binary phenotypes. *Eur. J. Hum. Genet.* **25**, 240–245 (2017).
8. Devlin, B. & Roeder, K. Genomic control for association studies. *Biometrics* **55**, 997–1004 (1999).
9. Mägi, R. *et al.* Trans-ethnic meta-regression of genome-wide association studies accounting for ancestry increases power for discovery and improves fine-mapping resolution. *Hum. Mol. Genet.* **26**, 3639–3650 (2017).
10. Mägi, R. & Morris, A. P. GWAMA: software for genome-wide association meta-analysis. *BMC Bioinformatics* **11**, 288 (2010).
11. Yang, J. *et al.* Conditional and joint multiple-SNP analysis of GWAS summary statistics identifies additional variants influencing complex traits. *Nat. Genet.* **44**, 369–375 (2012).
12. Mahajan, A. *et al.* Trans-ancestry genetic study of type 2 diabetes highlights the power of diverse populations for discovery and translation. *medRxiv* (2020).
13. Dastani, Z. *et al.* Novel loci for adiponectin levels and their influence on type 2 diabetes and metabolic traits: A multi-ethnic meta-analysis of 45,891 individuals. *PLoS Genet.* **8**, (2012).
14. Zheng, J. *et al.* LD Hub: A centralized database and web interface to perform LD score regression that maximizes the potential of summary level GWAS data for SNP

heritability and genetic correlation analysis. *Bioinformatics* **33**, 272–279 (2017).
15. Bulik-Sullivan, B. K. *et al.* LD Score regression distinguishes confounding from polygenicity in genome-wide association studies. *Nat Genet* **47**, 291–295 (2015).
16. Morris, A. P. *et al.* Large-scale association analysis provides insights into the genetic architecture and pathophysiology of type 2 diabetes. *Nat. Genet.* **44**, 981–990 (2012).
17. Manning, A. K. *et al.* A genome-wide approach accounting for body mass index identifies genetic variants influencing fasting glycemic traits and insulin resistance. *Nat. Genet.* **44**, 659–669 (2012).
18. Saxena, R. *et al.* Genetic variation in GIPR influences the glucose and insulin responses to an oral glucose challenge. *Nat. Genet.* **42**, 142–148 (2010).
19. Soranzo, N. *et al.* Common variants at 10 genomic loci influence hemoglobin A1C levels via glycemic and nonglycemic pathways. *Diabetes* **59**, 3229–3239 (2010).
20. Dupuis, J. *et al.* New genetic loci implicated in fasting glucose homeostasis and their impact on type 2 diabetes risk. *Nat. Genet.* **42**, 105–116 (2010).
21. Wickham, H. *ggplot2: Elegant Graphics for Data Analysis*. (Springer-Verlag New York, 2016).
22. Harrow, J. *et al.* GENCODE: The reference human genome annotation for the ENCODE project. *Genome Res.* **22**, 1760–1774 (2012).
23. Dunham, I. *et al.* An integrated encyclopedia of DNA elements in the human genome. *Nature* **489**, 57–74 (2012).
24. Pasquali, L. *et al.* Pancreatic islet enhancer clusters enriched in type 2 diabetes risk-associated variants. *Nat. Genet.* **46**, 136–143 (2014).
25. Varshney, A. *et al.* Genetic regulatory signatures underlying islet gene expression and type 2 diabetes. *Proc. Natl. Acad. Sci. U. S. A.* **114**, 2301–2306 (2017).
26. Pickrell, J. K. Joint Analysis of Functional Genomic Data and Genome-wide Association Studies of 18 Human Traits. *Am. J. Hum. Genet.* **94**, 559–573 (2014).
27. Maller, J. B. *et al.* Bayesian refinement of association signals for 14 loci in 3 common diseases. *Nat. Genet.* **44**, 1294–1301 (2012).
28. GTEx Consortium. The GTEx Consortium atlas of genetic regulatory effects across human tissues. *Science* **369**, 1318–1330 (2020).
29. Hemani, G. *et al.* Automating Mendelian randomization through machine learning to construct a putative causal map of the human phenome. *bioRxiv* 173682 (2017). doi:10.1101/173682
30. Hemani, G., Tilling, K. & Davey Smith, G. Orienting the causal relationship between imprecisely measured traits using GWAS summary data. *PLoS Genet.* **13**, (2017).
