## Supplementary Tables for "Trans-ancestry genome-wide association study of gestational diabetes mellitus highlights genetic links with type 2 diabetes"

**Table S1. GDM case and control ascertainment criteria for GWAS contributing to trans-ancestry meta-regression.**

| Study acronym | Study name | PMID | Ascertainment | | Ancestry group(s) |
|---|---|---|---|---|---|
| | | | Cases | Controls | |
| ALSPAC | Avon Longitudinal Study of Parents and Children | 22507742 | Clinical diagnosis noted in medical records or self-reported and no previous self-reported diabetes | Absence of clinical diagnosis noted in medical records or self-reported diabetes during or before the pregnancy | EUR |
| AM | ANDIS-MDC | https://doi.org/10.1101/2020.09.29.20203935 | Gestational diabetes diagnosis with no previous diabetes diagnosis | Healthy non-diabetic women | EUR |
| BIB | Born in Bradford | 23064411 | Modified WHO criteria applied to fasting and 2-hour postload glucose without previously recorded diabetes | Healthy women who did not have any record of diabetes during or before the pregnancy | EUR, SAS |
| BOTNIA | The Botnia Study | https://doi.org/10.1101/2020.09.29.20203935 | Gestational diabetes diagnosis with no previous diabetes diagnosis | Healthy women who did not have any record of diabetes during the pregnancy | EUR |
| DECODE | deCODE Genetics | - | Diagnosis of gestational diabetes in medical records from The National University Hospital of Iceland 1997-2016, ICD10: O24.4 and O24.9). | Women recruited through different genetic research projects at deCODE, matched with the cases on age, and county of origin. Women with type 2 diabetes were excluded from the control group. | EUR |
| EGCUT | Estonian Biobank, University of Tartu | 24518929 | Gestational diabetes diagnosis with no previous diabetes diagnosis | Women without GDM diagnosis | EUR |
| ERF | Erasmus Rucphen Family Study | 15054401 | Gestational diabetes diagnosis with no previous diabetes diagnosis | Women without GDM diagnosis | EUR |
| ESTGDM | Estonian Gestational Diabetes Samples | 29879500 | All women performed fasting oral glucose tolerance test with 75g glucose between gestational weeks 23 to 31 weeks. GDM was diagnosed according to the IADPSG criteria. | All women performed fasting oral glucose tolerance test with 75g glucose between gestational weeks 23 to 31 weeks. Women who stayed normoglycemic were classified as non-GDM. | EUR |
| FINNGEDI | The Finnish Gestational Diabetes Study | 32374401 | GDM status based on 75 g OGTT at 12-16 weeks (high risk women) or at 24-28 weeks of gestation. Fasting plasma | Women without GDM diagnosis (normal OGTT at 24-28 weeks of gestation). | EUR |

| | | | | | |
|---|---|---|---|---|---|
| | | | glucose ≥5.3 mmol/l, 1-hour glucose ≥10.0 mmol/l or 2- hour glucose ≥8.6 mmol/l. | | |
| GEN3G | Genetics of Glucose regulation in Gestation and Growth | 26842272 | All women performed a fasting 75g oral glucose tolerance test (OGTT) between 24 to 30 weeks of gestation. GDM was defined based on IADPSG criteria. Women also had a screening at 1st trimester (A1c and/or GCT) to exclude pre-existing diabetes prior to pregnancy | All women performed a fasting 75g oral glucose tolerance test (OGTT) between 24 to 30 weeks of gestation. Women who did not meet the IADPSG criteria for GDM were classified as "normoglycemic" controls. Women also had a screening at 1st trimester (A1c and/or GCT) to exclude pre-existing diabetes prior to pregnancy | EUR |
| GIFTS | Genomic and Lifestyle predictors of foetal outcome relevant to diabetes and obesity and their relevance to prevention strategies in South Asian peoples | 31711436 | GDM was diagnosed based on revised WHO 1999 guidelines. Women with a prior history of type 2 diabetes, or gestational diabetes or pregnancy induced hypertension were excluded. | Women without a GDM diagnosis that were recruited | SAS |
| HONGKONG | The Hyperglycemia and Adverse Pregnancy Outcome (HAPO) study conducted in the Hong Kong field centre | 28279981 | All pregnant women less than 31 weeks of gestation. GDM was diagnosed according to the World Health Organization 2013 criteria for women with either FPG ≥5.1 mmol/l, 1-hour glucose ≥10.0 mmol/l or 2- hour glucose ≥8.5 mmol/l during pregnancy | Healthy women who did not have any record of diabetes during the pregnancy | EAS |
| NFBC66 | Northern Finland Birth Cohort 1966 | - | Self-reported GDM (ever in their life) at 46yrs follow-up OR GDM diagnosis found in the birth register/Care Register for Health Care (registers up to 2012) | Self-reported (been pregnant but) never had GDM at 46yrs follow-up.<br>Men, and subjects with no information on pregnancies nor GDM were excluded | EUR |
| NFBC86 | Northern Finland Birth Cohort 1986 | - | Cohort subjects who had abnormal OGTT or blood glucose curve during pregnancy | Cohort subjects who had no indications (glucosuria, prior gestational diabetes, macrosomia in current pregnancy, previous macrosomic infant (weight more than 4500g), BMI > 25 and maternal age > 40 yr) and did not take OGTT OR OGTT was normal. Cohort subjects who had pre-pregnancy DM OR information on | EUR |

| | | | | | |
|---|---|---|---|---|---|
| | | | | indications and/or missing OGTT were excluded | |
| NU | Northwestern Feinburg School of Medicine | 23903356 | age<18 or > 40, multiple pregnancy, pregestational diabetes (type 1 or 2) or diabetes discovered at the first trimester, drugs and/or alcohol abuse, uncontrolled endocrine disease, renal failure, or other major medical conditions that would affect glucose regulation | Healthy women who did not have any record of diabetes during the pregnancy | AFR, EAS, EUR, HIS |
| PREDO | The Prediction and Prevention of Preeclampsia and Intrauterine Growth Restriction study | 27639277 | Gestational diabetes diagnose based on 75 g glucose tolerance test. Fasting plasma glucose ≥5.2 mmol/l, 1-hour glucose ≥10.0 mmol/l or 2- hour glucose ≥8.7 mmol/l. | Women with no record of gestational diabetes, type 1 or type 2 DM. Control women may have other pregnancy disorders | EUR |
| SNUH | Seoul National University Hospital | 22233651 | The GDM case group was selected from a hospital-based cohort enrolled between January 1996 and February 2003 from Cheil General Hospital. A 50-g 1-h glucose challenge test was performed during 24–28 weeks' gestation in order to screen for GDM, and a glucose level of >= 7.2 mmol/L was considered positive and warranted a diagnostic 100-g oral glucose tolerance test. Glucose and insulin concentrations were measured at 0, 1, 2, and 3 h of the glucose challenge. GDM was diagnosed according to the criteria of the Third International Workshop Conference on GDM. The thresholds for the diagnosis of GDM were as follows: fasting >= 5.8 mmol/L, 1 h >= 10.6 mmol/L, 2 h >= 9.2 mmol/L, and 3 h >= 8.1 mmol/L. | Nondiabetic control subjects were selected from two population-based cohort studies, the rural Ansung and the urban Ansan cohorts. The two cohorts comprised the Korean Genome Epidemiology Study and included 5,018 and 5,020 subjects, respectively. Only women were eligible for enrollment. From the Korean Genome Epidemiology Study subjects, we included 1,242 nondiabetic women according to the following criteria: age >= 50 years, no previous history of type 2 diabetes, no first-degree relatives with type 2 diabetes, fasting plasma glucose level < 5.6 mmol/L, and HbA1c < 6.0%. Information on parity and glucose tolerance status during pregnancy was not available for the control group. | EAS |
| STORK | Project STORK | 22108914 | Diagnosis based on WHO99 criteria | Women without GDM diagnosis | EUR |

| | | | | | |
|---|---|---|---|---|---|
| STORKG | STORK Groruddalen | 21062840 | Universal screening of GDM with 75 g oral glucose tolerance test in pregnancy week 28. GDM diagnosed according to WHO99 criteria. Normoglycemic at inclusion in pregnancy week 15. | Normal fasting glucose and 2-hour glucose after 75 g oral glucose tolerance test in pregnancy week 28. | EUR |
| UKBB | UK Biobank | 25826379, 30305743 | - O24.4 (Diabetes mellitus arising in pregnancy) or O24.9 (Diabetes mellitus in pregnancy, unspecified) diagnosis in the Hospital inpatient admissions data<br>- GDM self-report diagnosis (code 1221, field 20002 “Non-cancer illness code, self-reported”)<br>- GDM self-report diagnosis using field 4041 "Did you only have diabetes during pregnancy?"<br>- E10 - E14 diagnosis in the hospital inpatient admission data, followed by an obstetrical diagnosis within 0–120 days or if it preceded an obstetrical diagnosis by 0–180 days | Women with at least one pregnancy (number of births field field 2734) and without GDM or T2D diagnoses | EUR |
| VIVA | Project Viva | 24639442 | We defined gestational glucose tolerance in categories according to the results of routine prenatal screening. Clinicians screened all pregnant women at 26–28 weeks of gestation with a non-fasting oral glucose challenge test, in which venous blood was sampled 1 h after a 50 g oral glucose load. If the blood glucose exceeded 140 mg/dl, the clinician referred the woman for a fasting 3-h 100 g oral glucose tolerance test (OGTT). Abnormal OGTT results were a blood glucose >95 mg/dl fasting, >180 mg/dl at 1 h, >155 mg/dl at 2 h, or >140 mg/dl at 3 h. Women with two or more abnormal values on the OGTT | Women without GDM. The control group included women with GIGT, IH, and normal glucose as controls. | EUR |

| | | | |
|---|---|---|---|
| | | | were diagnosed with GDM. We further classified those with one abnormal value on the OGTT as having gestational impaired glucose tolerance (GIGT), those with an abnormal glucose challenge test but a normal OGTT as having isolated hyperglycemia (IH) and the remaining women as having normal glucose tolerance. |

### Table S2. Sample characteristics of GWAS contributing to trans-ancestry meta-regression.

| Study acronym | Population group | Country of origin | Genotyping array(s) | Effective sample size | GDM status | Sample size | Age (years) mean (SD) | BMI (kg/m²) mean (SD) |
|---|---|---|---|---|---|---|---|---|
| **African ancestry** | | | | | | | | |
| NU (AFR) | African American | USA | Illumina Human1M-Duov3 B SNP array | 333.2 | Case | 91 | 29.4 (6.4) | 32.1 (7.5) |
| | | | | | Control | 985 | 24.9 (5.3) | 27.2 (5.7) |
| **East Asian ancestry** | | | | | | | | |
| HONGKONG | Chinese | Hong Kong | Illumina InfiniumOmniZhongHua-8 v1.3 BeadChip | 500.1 | Case | 148 | 33.1 (4.3) | 21.7 (3.1) |
| | | | | | Control | 805 | 30.9 (4.6) | 20.8 (2.8) |
| NU (EAS) | Thai | Thai | Illumina HumanOmni1-Quad v1-0 B SNP array | 791.9 | Case | 251 | 29.4 (5.7) | 26.6 (3.9) |
| | | | | | Control | 937 | 27.3 (5.4) | 25.4 (3.5) |
| SNUH | Korean | South Korea | Affymetrix Genome-Wide Human Single Nucleotide Polymorphism (SNP) Array 5.0 | 1359.7 | Case | 468 | 31.5 (4.0) | 23.3 (3.2) |
| | | | | | Control | 1,242 | 59.1 (5.6) | 24.6 (3.2) |
| **European ancestry** | | | | | | | | |
| ALSPAC | British | UK | Illumina human660W-quad array | 257.5 | Case | 65 | 28.6 (5.2) | 25.9 (6.3) |
| | | | | | Control | 6,803 | 27.9 (4.7) | 22.9 (3.7) |
| AM | Swedish | Sweden | Illumina InfiniumCoreExome-24v1-1 | 1019.2 | Case | 305 | - | - |
| | | | | | Control | 1,548 | - | - |
| BIB (EUR) | British | UK | Illumina HumanCoreExome12v1.0, HumanCoreExome12v1.1, HumanCoreExome24v1.0 | 580.7 | Case | 153 | 30.2 (5.5) | 29.3 (6.8) |
| | | | | | Control | 2,837 | 26.5 (6.0) | 26.7 (6.0) |
| BOTNIA | Finnish | Finland | Illumina HumanOmniExpress-12v1-1 | 250.2 | Case | 69 | - | - |
| | | | | | Control | 670 | - | - |
| DECODE | Icelandic | Iceland | various Illumina SNP arrays | 2980.4 | Case | 750 | 32.4 (5.5) | 30.7 (6.8) |
| | | | | | Control | 114,000 | - | 25.6 (4.9) |
| EGCUT | Estonian/Russian | Estonia | Illumina Global Screening Array, Illumina OmniExpress, Illumina HumanHap 370CNV, Illumina CoreExomeChip | 1017.1 | Case | 257 | 27.7 (6.1) | 24.7 (4.7) |
| | | | | | Control | 23,874 | 43.9 (16.6) | 25.8 (5.3) |
| ERF | Dutch | The Netherlands | Illumina 318K, Illumina 370K and Affymetrix 250K | 188.4 | Case | 49 | 49.2 (13.2) | 28.5 (5.2) |
| | | | | | Control | 1,222 | 50.9 (12.7) | 27.0 (5.0) |
| ESTGDM | Estonian/Russian | Estonia | Illumina Global Screening Array | 163.7 | Case | 52 | 32.2 (5.3) | 26.8 (6.0)- |
| | | | | | Control | 192 | 30.0 (5.4) | 25.0(4.7) |
| FINNGEDI | Finnish | Finland | Infinium Omni2.5-8 BeadChip | 529.2 | Case | 298 | 32.5 (5.3) | 27.9 (6.1) |
| | | | | | Control | 238 | 31.5 (5.2) | 25.6 (4.8) |
| GEN3G | Canadian | Canada | Illumina Infinium Expanded Multi-Ethnic Genotyping Array (MEGA EX) | 162.0 | Case | 44 | 29.7 (5.9) | 28.6 (7.3) |
| | | | | | Control | 511 | 28.1 (4.1) | 25.5 (5.6) |
| NFBC66 | Finnish | Finland | Illumina HumanCNV370DUO Analysis BeadChip | 782.7 | Case | 220 | - | 28.9 (6.2) |
| | | | | | Control | 1,769 | - | 26.3 (5.1) |
| NFBC86 | Finnish | Finland | Human Omni Express Exome 8v1.2 | 106.8 | Case | 27 | 29.3 (6.2) | 23.8 (4.4) |
| | | | | | Control | 2,441 | 27.4 (5.0) | 27.4 (5.0) |

| | | | | | | | | |
|---|---|---|---|---|---|---|---|---|
| NU (EUR) | European American | USA | Illumina Human 610 Quad v1 B SNP array | 589.5 | Case | 168 | 32.6 (4.9) | 31.0 (5.8) |
| | | | | | Control | 1,200 | 31.1 (5.3) | 28.0 (4.5) |
| PREDO | Finnish | Finland | Illumina Global Screening Array | 653.9 | Case | 213 | 32.8 (5.6) | 30.7 (6.7) |
| | | | | | Control | 703 | 31.9 (5.8) | 26.5 (6.1) |
| STORK | Norwegian | Norway | Illumina Infinium CoreExome chip | 39.2 | Case | 10 | 31.3 (3.9) | 27.6 (3.3) |
| | | | | | Control | 497 | 31.7 (4.4) | 23.4 (3.7) |
| STORKG | European | Norway | Illumina Infinium CoreExome chip | 130.4 | Case | 37 | 30.9 (4.5) | 24.5 (4.7) |
| | | | | | Control | 275 | 31.3 (4.9) | 25.9 (5.1) |
| UKBB | British | UK | UK Biobank Axiom Array and UK BiLEVE Axiom Array | 4117.0 | Case | 1,035 | 52.4 (8.3) | 29.1 (6.2) |
| | | | | | Control | 185,550 | 57.1 (7.8) | 26.9 (4.9) |
| VIVA | American | USA | Illumina Infinium Expanded Multi-Ethnic Genotyping Array (MEGA EX) | 103.1 | Case | 27 | 33.2 (4.6) | 27.2 (6.5) |
| | | | | | Control | 572 | 33.3 (4.4) | 24.1 (4.6) |
| **Hispanic/Latino ancestry** | | | | | | | | |
| NU (HIS) | Hispanic American | USA | Illumina Human1M-Duov3 B SNP array | 543.3 | Case | 174 | 30.8 (5.5) | 33.1 (6.4) |
| | | | | | Control | 619 | 28.3 (5.3) | 29.1 (4.9) |
| **South Asian ancestry** | | | | | | | | |
| BIB (SAS) | Pakistani British | UK | Illumina HumanCoreExome12v1.0, HumanCoreExome12v1.1, HumanCoreExome24v1.0 | 1437.6 | Case | 405 | 30.7 (5.5) | 28.4 (6.1) |
| | | | | | Control | 3,192 | 27.5 (5.0) | 25.5 (5.3) |
| GIFTS (1) | Bangladeshi | UK | Illumina Global Screening Array | 125.8 | Case | 41 | 20.0 (2.6) | 21.5 (2.6) |
| | | | | | Control | 135 | 20.3 (3.0) | 20.5 (3.4) |
| GIFTS (2) | Bangladeshi | Bangladesh | Illumina Global Screening Array | 37.1 | Case | 11 | 23.1 (4.1) | 23.6 (4.0) |
| | | | | | Control | 59 | 22.5 (4.3) | 21.9 (3.9) |
| GIFTS (3) | Bangladeshi | Bangladesh | Illumina Global Screening Array | 354.4 | Case | 116 | 30.4 (5.7) | 27.2 (4.0) |
| | | | | | Control | 375 | 29.1 (5.3) | 25.6 (4.5) |

**Table S3. Study-level quality control, pre-phasing, imputation and association analysis of GWAS contributing to trans-ancestry meta-regression.**

| Study acronym | Sample QC | SNV QC | | Imputation | | | Association analysis | | | | |
|---|---|---|---|---|---|---|---|---|---|---|---|
| | Call rate | Call rate | HWE *p*-value | Reference panel | Software | QC | QC+ SNVs | Relatedness and structure | Software | Covariates | $\lambda_{GC}$ |
| **African ancestry** | | | | | | | | | | | |
| NU (AFR) | >95% | >95% | $>1\times10^{-4}$ | 1000G Phase 3 | SHAPEIT2 + IMPUTEv2 | info > 0.4 | 9,743,681 | PC adjustment | EPACTS | - | 1.01 |
| **East Asian ancestry** | | | | | | | | | | | |
| HONGKONG | >98% | >99% | $>1\times10^{-4}$ | 1000G Phase 3 | SHAPEIT2 (phasing) + Michigan imputation server | $r^2$>0.4 | 5,986,977 | PC adjustment | EPACTS | - | 1.01 |
| NU (EAS) | >95% | >95% | $>1\times10^{-4}$ | 1000G Phase 3 | SHAPEIT2 + IMPUTEv2 | info > 0.4 | 6,698,571 | PC adjustment | EPACTS | - | 0.98 |
| SNUH | >95% | >97% | $>1\times10^{-6}$ | 1000G Phase 3 | Michigan imputation server | info > 0.4 | 5,751,976 | Relateds removed, PC adjustment | EPACTS | - | 1.02 |
| **European ancestry** | | | | | | | | | | | |
| ALSPAC | >95% | >95% | $>1\times10^{-6}$ | HRC | SHAPEIT2 + Sanger imputation server | $r^2$>0.4 | 5,723,169 | PC adjustment | EPACTS | - | 1.10 |
| AM | >95% | >95% | $>1\times10^{-4}$ | HRC | MINIMAC3 | info > 0.4 | 7,201,324 | PC adjustment | EPACTS | - | 1.02 |
| BIB (EUR) | ≥99.5% | ≥99.5% | | Merged UK10K & 1000G Phase 3 | EAGLE2 + Sanger Imputation Service | | 6,411,781 | Mixed model | GCTA | - | 1.02 |
| BOTNIA | >95% | >95% | $>1\times10^{-4}$ | 1000G Phase 3 | MINIMAC3 | info > 0.4 | 5,825,870 | PC adjustment | EPACTS | - | 1.03 |
| DECODE | >98% | >98% | $>1\times10^{-6}$ | Icelandic reference panel | deCODE software | info > 0.4 | 8,356,461 | LD-score regression intercept | deCODE software | - | 1.01 |
| EGCUT | >95% | >95% | $>1\times10^{-4}$ | Estonian reference panel | SHAPEIT2 + IMPUTEv2 | info > 0.4 | 6,695,203 | Mixed model | EPACTS | - | 1.04 |
| ERF | >98% | >98% | $>1\times10^{-6}$ | HRC | Michigan imputation server | $r^2$>0.4 | 4,459,573 | Mixed model | Rvtest | Age | 1.00 |
| ESTGDM | >95% | >95% | $>1\times10^{-4}$ | Estonian Reference panel | SHAPEIT2 + IMPUTEv2 | info > 0.4 | 5,297,361 | Mixed model | EPACTS | - | 0.99 |
| FINNGEDI | >95% | >95% | $>1\times10^{-4}$ | HRC | SHAPEIT2 + PBWT | info > 0.4 | 7,646,486 | Relateds removed, PC adjustment | PLINK | Age, BMI | 1.05 |
| GEN3G | >98% | >95% | >1x10-8 | HRC | ShapeIT v2.r790 (phasing) + Michigan imputation server | info > 0.4 | 5,120,027 | None | EPACTS | - | 1.04 |
| NFBC66 | >95% | >95% | $>1\times10^{-4}$ | HRC | IMPUTE2 | info > 0.4 | 7,302,500 | Relateds removed, PC adjustment | EPACTS | - | 1.02 |
| NFBC86 | >95% | >95% | $>1\times10^{-4}$ | HRC | IMPUTE2 | info > 0.4 | 4,416,827 | Relateds removed, PC adjustment | EPACTS | - | 1.11 |
| NU (EUR) | >95% | >95% | $>1\times10^{-4}$ | HRC | SHAPEIT2 + IMPUTEv2 | info > 0.4 | 7,041,417 | PC adjustment | EPACTS | - | 1.03 |
| PREDO | >95% | >95% | $>1\times10^{-6}$ | Finnish SISu v2 imputation panel | Eagle v2.3 + IMPUTEv2 | info > 0.4 | 6,910,245 | PC adjustment | EPACTS | - | 0.97 |
| STORK | >95% | >95% | $>1\times10^{-6}$ | 1000G Phase 3 | SHAPEIT2 + IMPUTEv2 | info > 0.4 | 2,271,920 | Relateds removed | SNPTEST | - | 1.01 |
| STORKG | >95% | >95% | $>1\times10^{-6}$ | 1000G Phase 3 | SHAPEIT2 + IMPUTEv2 | info > 0.4 | 4,607,366 | Relateds removed | EPACTS | - | 1.01 |

| | | | | | | | | | | | |
|---|---|---|---|---|---|---|---|---|---|---|---|
| UKBB | >99% | >95% | >1x10$^{-12}$ | HRC and merged UK10K & 1000G Phase 3 | IMPUTE4 | info > 0.4 | 8,827,701 | Mixed-model | BOLT-LMM | Age, genotyping array | 1.02 |
| VIVA | >95% | >98% | >1x10$^{-6}$ | HRC | ShapeIT v2.r790 + Michigan imputation server | info > 0.4 | 4,395,851 | PC adjustment | EPACTS | - | 1.05 |
| **Hispanic/Latino ancestry** | | | | | | | | | | | |
| NU (HIS) | >95% | >95% | | 1000G Phase 3 | SHAPEIT2 + IMPUTEv2 | info > 0.4 | 7,368,540 | PC adjustment | EPACTS | - | 1.02 |
| **South Asian ancestry** | | | | | | | | | | | |
| BIB (SAS) | ≥99.5% | ≥99.5% | | 1000GPhase 3 | EAGLE2 + Sanger Imputation Service | info > 0.4 | 7,517,804 | Mixed model | GCTA | - | 1.00 |
| GIFTS (1) | >98% | >98% | >1x10$^{-6}$ | HRC | Michigan imputation server | info > 0.4 | 5,169,445 | Relateds removed, PC adjustment | EPACTS | - | 0.97 |
| GIFTS (2) | >98% | >98% | >1x10$^{-6}$ | HRC | Michigan imputation server | info > 0.4 | 2,570,490 | Relateds removed, PC adjustment | EPACTS | - | 0.99 |
| GIFTS (3) | >98% | >98% | >1x10$^{-6}$ | HRC | Michigan imputation server | info > 0.4 | 6,519,637 | Relateds removed, PC adjustment | EPACTS | - | 0.98 |

**Table S6. Ancestry-specific GDM association summary statistics for lead SNVs at loci attaining genome-wide significance ($p$<5x10$^{-8}$) in the trans-ancestry meta-regression (MR-MEGA).**

*MTNR1B* locus. Lead SNV: rs10830963. Risk/other allele: G/C

| Ancestry | RAF | Association *p*-value | OR (95% CI) | Cases/Controls |
|---|---|---|---|---|
| African | 0.046 | 0.94 | 1.04 (0.35-3.15) | 91/985 |
| East Asian | 0.432 | 4.6x10$^{-12}$ | 1.51 (1.34-1.69) | 867/2,984 |
| European | 0.264 | 2.3x10$^{-42}$ | 1.42 (1.35-1.49) | 3,523/323,982 |
| Hispanic/Latino | 0.243 | 0.010 | 1.42 (1.09-1.85) | 174/619 |
| South Asian | 0.403 | 0.0016 | 1.22 (1.08-1.39) | 573/3,761 |

*TCF7L2* locus. Lead SNV: rs7903146. Risk/other allele: T/C

| Ancestry | RAF | Association *p*-value | OR (95% CI) | Cases/Controls |
|---|---|---|---|---|
| African | 0.296 | 0.22 | 0.70 (0.39-1.24) | 91/985 |
| East Asian | 0.034 | 0.14 | 1.24 (0.93-1.66) | 867/2,984 |
| European | 0.284 | 8.7x10$^{-16}$ | 1.22 (1.17-1.29) | 3,780/347,856 |
| Hispanic/Latino | 0.224 | 0.46 | 1.11 (0.84-1.48) | 174/619 |
| South Asian | 0.313 | 0.0061 | 1.21 (1.06-1.39) | 573/3,761 |

*CDKAL1* locus. Lead SNV: rs9348441. Risk/other allele: A/T

| Ancestry | RAF | Association *p*-value | OR (95% CI) | Cases/Controls |
|---|---|---|---|---|
| African | 0.185 | 0.23 | 0.68 (0.36-1.28) | 91/985 |
| East Asian | 0.401 | 6.8x10$^{-14}$ | 1.53 (1.37-1.72) | 867/2,984 |
| European | 0.256 | 0.015 | 1.06 (1.01-1.12) | 3,780/347,856 |
| Hispanic/Latino | 0.258 | 0.086 | 1.26 (0.97-1.65) | 174/619 |
| South Asian | 0.261 | 0.082 | 1.13 (0.98-1.31) | 573/3,761 |

*CDKN2A-CDKN2B* locus. Lead SNV: rs10811662. Risk/other allele: G/A

| Ancestry | RAF | Association *p*-value | OR (95% CI) | Cases/Controls |
|---|---|---|---|---|
| African | 0.885 | 0.15 | 0.58 (0.27-1.22) | 91/985 |
| East Asian | 0.572 | 0.0017 | 1.20 (1.07-1.34) | 867/2,984 |
| European | 0.824 | 2.4x10$^{-7}$ | 1.18 (1.11-1.25) | 3,770/347,359 |
| Hispanic/Latino | 0.860 | 0.13 | 1.36 (0.92-2.02) | 174/619 |
| South Asian | 0.547 | 0.16 | 0.90 (0.78-1.04) | 562/3,702 |

*HKDC1* locus. Lead SNV: rs9663238. Risk/other allele: G/A

| Ancestry | RAF | Association *p*-value | OR (95% CI) | Cases/Controls |
|---|---|---|---|---|
| African | 0.217 | 0.22 | 0.70 (0.40-1.23) | 91/985 |
| East Asian | 0.264 | 0.0011 | 1.22 (1.08-1.38) | 867/2,984 |
| European | 0.708 | 3.7x10$^{-7}$ | 1.14 (1.09-1.20) | 3,557/346,656 |
| Hispanic/Latino | 0.531 | 0.77 | 1.04 (0.81-1.32) | 174/619 |
| South Asian | 0.585 | 0.094 | 1.11 (0.98-1.26) | 573/3,761 |

**Table S5. Association of previously reported SNVs for T2D (180,834 cases and 1,159,055 controls of diverse ancestry from the DIAMANTE Consortium) on GDM in 5,485 cases and 347,856 controls of diverse ancestry.**

| Locus | Lead SNV | Chr | Position (bp, b37) | Alleles | | T2D association | | | GDM association | | |
|---|---|---|---|---|---|---|---|---|---|---|---|
| | | | | T2D risk | Other | Beta | SE | *p*-value | Beta | SE | *p*-value |
| *VWA5B1* | rs10916784 | 1 | 20,729,451 | G | C | 0.0342 | 0.0051 | 1.2E-11 | 0.0604 | 0.0204 | 1.8E-02 |
| *MACF1* | rs3768301 | 1 | 39,870,793 | T | C | 0.0692 | 0.0063 | 6.2E-31 | 0.0157 | 0.0259 | 7.7E-02 |
| *MAST2* | rs34444543 | 1 | 46,358,862 | G | A | 0.0349 | 0.0053 | 5.5E-13 | 0.0169 | 0.0216 | 6.8E-01 |
| *FAF1* | rs12073283 | 1 | 51,219,188 | C | G | 0.0726 | 0.0086 | 6.7E-18 | 0.0845 | 0.0366 | 8.4E-02 |
| *PGM1* | rs11576729 | 1 | 64,114,429 | G | T | 0.0487 | 0.0062 | 2.5E-17 | 0.0318 | 0.0247 | 1.5E-02 |
| *PTGFRN* | rs1127215 | 1 | 117,532,790 | C | T | 0.0408 | 0.0050 | 3.9E-17 | 0.0176 | 0.0202 | 7.2E-01 |
| *NOTCH2* | rs835576 | 1 | 120,455,586 | C | T | 0.0637 | 0.0081 | 2.8E-17 | 0.0741 | 0.0311 | 1.1E-01 |
| *SEC16B* | rs539515 | 1 | 177,889,025 | C | A | 0.0536 | 0.0060 | 4.6E-20 | 0.0254 | 0.0250 | 4.2E-01 |
| *ZNF281* | rs10919928 | 1 | 200,416,099 | A | G | 0.0339 | 0.0079 | 4.6E-09 | N/A | N/A | N/A |
| *DSTYK* | rs6689629 | 1 | 204,539,291 | A | G | 0.0382 | 0.0059 | 2.4E-10 | 0.0024 | 0.0242 | 8.8E-01 |
| *SRGAP2* | rs9429893 | 1 | 206,600,992 | A | G | 0.0316 | 0.0053 | 2.3E-09 | 0.0192 | 0.0201 | 3.3E-01 |
| *PROX1* | rs340874 | 1 | 214,159,256 | C | T | 0.0562 | 0.0050 | 2.5E-33 | 0.0521 | 0.0199 | 9.5E-02 |
| *LYPLAL1* | rs2820446 | 1 | 219,748,818 | C | G | 0.0444 | 0.0054 | 2.7E-18 | -0.0028 | 0.0217 | 9.9E-01 |
| *ABCB10* | rs348330 | 1 | 229,672,955 | G | A | 0.0451 | 0.0054 | 3.7E-18 | 0.0428 | 0.0211 | 2.0E-01 |
| *TMEM18* | rs6548240 | 2 | 636,929 | A | C | 0.0673 | 0.0071 | 2.2E-25 | 0.0270 | 0.0275 | 5.2E-01 |
| *DTNB* | rs55928417 | 2 | 25,533,568 | G | T | 0.0341 | 0.0053 | 4.0E-11 | 0.0217 | 0.0205 | 8.1E-01 |
| *GCKR* | rs1260326 | 2 | 27,730,940 | C | T | 0.0625 | 0.0050 | 4.6E-38 | 0.1090 | 0.0207 | 6.8E-07 |
| *THADA* | rs13414140 | 2 | 43,671,176 | C | T | 0.0881 | 0.0087 | 5.2E-31 | 0.0189 | 0.0352 | 3.1E-01 |
| *SIX3-SIX2* | rs12712928 | 2 | 45,192,080 | C | G | 0.0091 | 0.0059 | 2.4E-14 | -0.0343 | 0.0258 | 6.8E-01 |
| *BNIPL* | rs17049712 | 2 | 58,961,136 | T | C | 0.0342 | 0.0056 | 2.3E-09 | 0.0094 | 0.0221 | 6.8E-01 |
| *BCL11A* | rs243018 | 2 | 60,586,707 | G | C | 0.0595 | 0.0050 | 6.7E-35 | 0.0104 | 0.0200 | 4.3E-01 |
| *CEP68* | rs6752053 | 2 | 65,666,674 | T | C | 0.0527 | 0.0054 | 4.1E-24 | 0.0147 | 0.0208 | 5.2E-01 |
| *GLI2* | rs11677557 | 2 | 121,317,747 | A | G | 0.0527 | 0.0071 | 6.5E-15 | 0.0144 | 0.0293 | 9.6E-01 |
| *CYTIP* | rs7594480 | 2 | 158,390,468 | T | C | 0.0760 | 0.0107 | 4.0E-12 | 0.1226 | 0.0471 | 7.6E-04 |
| *RBSM1* | rs1020731 | 2 | 161,144,055 | A | G | 0.0307 | 0.0056 | 1.2E-09 | 0.0427 | 0.0242 | 1.9E-01 |
| *KCNH7* | rs12614955 | 2 | 163,649,480 | T | C | 0.0323 | 0.0051 | 8.6E-10 | 0.0096 | 0.0224 | 3.3E-01 |
| *GRB14* | rs10184004 | 2 | 165,508,389 | C | T | 0.0642 | 0.0056 | 5.2E-34 | 0.0206 | 0.0224 | 8.5E-01 |
| *IKZF2* | rs16849467 | 2 | 213,818,731 | T | C | 0.0367 | 0.0062 | 4.8E-09 | -0.0338 | 0.0241 | 5.0E-01 |

| | | | | | | | | | | | |
|---|---|---|---|---|---|---|---|---|---|---|---|
| *IRS1* | rs2943648 | 2 | 227,100,490 | G | A | 0.0820 | 0.0058 | 3.6E-52 | 0.0361 | 0.0229 | 2.3E-01 |
| *ATG16L1* | rs117809958 | 2 | 234,191,103 | A | T | 0.2114 | 0.0313 | 5.6E-12 | N/A | N/A | N/A |
| *PPARG* | rs17036160 | 3 | 12,329,783 | C | T | 0.1059 | 0.0086 | 2.9E-38 | 0.0739 | 0.0312 | 1.8E-02 |
| *UBE2E2* | rs13094957 | 3 | 23,457,080 | T | C | 0.0780 | 0.0059 | 5.2E-48 | 0.0592 | 0.0240 | 9.4E-02 |
| *RBM6* | rs2624847 | 3 | 50,174,197 | G | T | 0.0333 | 0.0060 | 6.2E-11 | 0.0020 | 0.0237 | 6.7E-01 |
| *CACNA2D3* | rs76263492 | 3 | 54,828,827 | T | G | 0.0918 | 0.0150 | 8.7E-10 | N/A | N/A | N/A |
| *PXK* | rs12629058 | 3 | 58,338,809 | T | C | 0.0358 | 0.0057 | 3.5E-11 | 0.0425 | 0.0224 | 1.8E-01 |
| *PSMD6-ADAMTS9* | rs704360 | 3 | 63,884,800 | G | A | 0.0676 | 0.0063 | 8.0E-33 | 0.0488 | 0.0280 | 5.1E-01 |
| *ZBTB20* | rs1459513 | 3 | 114,960,798 | C | A | 0.0519 | 0.0069 | 9.6E-15 | 0.0243 | 0.0327 | 8.8E-01 |
| *ADCY5* | rs11708067 | 3 | 123,065,778 | A | G | 0.0950 | 0.0071 | 3.4E-46 | 0.0815 | 0.0257 | 3.7E-02 |
| *SLC12A8* | rs9873519 | 3 | 124,921,457 | T | C | 0.0375 | 0.0050 | 6.5E-14 | 0.0422 | 0.0202 | 2.2E-01 |
| *MBNL1* | rs9877505 | 3 | 152,432,042 | C | T | 0.0378 | 0.0051 | 1.4E-13 | 0.0408 | 0.0212 | 3.6E-01 |
| *SLC2A2* | rs8192675 | 3 | 170,724,883 | T | C | 0.0542 | 0.0055 | 3.5E-28 | 0.0297 | 0.0224 | 7.8E-03 |
| *IGF2BP2* | rs7633675 | 3 | 185,510,613 | G | T | 0.1177 | 0.0053 | 5.8E-131 | 0.0996 | 0.0211 | 1.4E-06 |
| *ST6GAL1* | rs3887925 | 3 | 186,665,645 | T | C | 0.0458 | 0.0049 | 3.8E-22 | 0.0387 | 0.0197 | 1.1E-01 |
| *LPP* | rs4686471 | 3 | 187,740,899 | C | T | 0.0553 | 0.0062 | 2.9E-21 | 0.0350 | 0.0220 | 2.8E-01 |
| *TFRC* | rs74289356 | 3 | 195,825,077 | T | C | 0.0463 | 0.0068 | 6.8E-12 | 0.0501 | 0.0322 | 3.0E-02 |
| *PCGF3-MAEA* | rs730831 | 4 | 1,240,299 | T | G | 0.0917 | 0.0074 | 7.6E-41 | 0.0299 | 0.0243 | 8.4E-02 |
| *WFS1* | rs9998835 | 4 | 6,293,237 | G | C | 0.0781 | 0.0058 | 3.1E-51 | 0.0450 | 0.0218 | 1.3E-01 |
| *LCORL* | rs6855926 | 4 | 18,047,401 | A | G | 0.0354 | 0.0053 | 8.4E-12 | 0.0201 | 0.0216 | 7.3E-01 |
| *GNPDA2* | rs13130484 | 4 | 45,175,691 | T | C | 0.0403 | 0.0051 | 1.3E-15 | 0.0215 | 0.0211 | 8.3E-01 |
| *MOB1B* | rs7674402 | 4 | 71,835,822 | A | G | 0.0579 | 0.0078 | 1.2E-13 | 0.1003 | 0.0422 | 1.7E-01 |
| *SCD5* | rs10471048 | 4 | 83,587,562 | G | C | 0.0310 | 0.0050 | 2.3E-11 | 0.0213 | 0.0206 | 6.2E-01 |
| *NKX6-1-CDS1* | rs117624659 | 4 | 85,339,618 | T | C | 0.2135 | 0.0281 | 3.1E-14 | N/A | N/A | N/A |
| *SMARCAD1* | rs6821438 | 4 | 95,091,911 | A | G | 0.0209 | 0.0050 | 2.5E-09 | 0.0326 | 0.0201 | 5.6E-01 |
| *PPP3CA* | rs2659518 | 4 | 102,135,363 | A | G | 0.0383 | 0.0066 | 4.5E-09 | 0.0118 | 0.0314 | 6.5E-01 |
| *SLC9B1* | rs223423 | 4 | 103,725,894 | G | A | 0.0210 | 0.0048 | 1.6E-09 | 0.0454 | 0.0198 | 6.2E-02 |
| *TET2* | rs17035289 | 4 | 106,048,291 | C | T | 0.0410 | 0.0062 | 1.3E-12 | 0.0513 | 0.0253 | 2.8E-01 |
| *TMEM154* | rs6813195 | 4 | 153,520,475 | C | T | 0.0622 | 0.0051 | 3.0E-36 | 0.0398 | 0.0217 | 2.9E-01 |
| *PDGFC* | rs1425482 | 4 | 157,725,916 | T | C | 0.0287 | 0.0055 | 2.3E-11 | 0.0198 | 0.0226 | 8.9E-02 |
| *ACSL1* | rs1996546 | 4 | 185,714,289 | G | T | 0.0621 | 0.0086 | 1.6E-13 | 0.0139 | 0.0319 | 1.5E-01 |
| *ANKH* | rs6885132 | 5 | 14,768,092 | C | G | 0.0586 | 0.0064 | 9.5E-23 | 0.0260 | 0.0296 | 7.8E-01 |
| *MRPS30* | rs6884702 | 5 | 44,682,589 | G | A | 0.0306 | 0.0049 | 6.0E-12 | 0.0130 | 0.0201 | 5.4E-01 |
| *ITGA1* | rs17261179 | 5 | 51,791,225 | T | C | 0.0335 | 0.0050 | 6.9E-11 | 0.0309 | 0.0199 | 1.3E-01 |
| *ARL15* | rs7736354 | 5 | 53,297,591 | T | C | 0.0444 | 0.0053 | 1.7E-20 | 0.0155 | 0.0223 | 9.1E-02 |

| | | | | | | | | | | | |
|---|---|---|---|---|---|---|---|---|---|---|---|
| *ANKRD55* | rs465002 | 5 | 55,808,475 | T | C | 0.0740 | 0.0053 | 1.5E-51 | 0.0175 | 0.0216 | 1.8E-01 |
| *PIK3R1* | rs57634870 | 5 | 67,716,793 | G | T | 0.0423 | 0.0060 | 6.0E-12 | 0.0464 | 0.0268 | 4.3E-01 |
| *POC5* | rs2307111 | 5 | 75,003,678 | T | C | 0.0349 | 0.0050 | 1.3E-18 | 0.0331 | 0.0201 | 6.4E-02 |
| *ZBED3* | rs7732130 | 5 | 76,435,004 | G | A | 0.0588 | 0.0064 | 6.0E-22 | 0.0859 | 0.0235 | 6.1E-03 |
| *DMGDH* | rs10052346 | 5 | 78,472,599 | G | T | 0.0348 | 0.0050 | 5.7E-14 | 0.0230 | 0.0209 | 6.1E-01 |
| *SLCO6A1-PAM* | rs115505614 | 5 | 102,422,968 | T | C | 0.1625 | 0.0151 | 5.9E-29 | 0.0959 | 0.0525 | 3.7E-02 |
| *CEP120* | rs4267865 | 5 | 122,704,342 | G | T | 0.0741 | 0.0105 | 9.7E-13 | -0.0257 | 0.0470 | 5.2E-01 |
| *PHFI5* | rs329122 | 5 | 133,864,599 | A | G | 0.0392 | 0.0049 | 6.0E-16 | 0.0131 | 0.0199 | 8.8E-01 |
| *NSD1* | rs244708 | 5 | 176,589,585 | G | A | 0.0294 | 0.0050 | 3.8E-09 | 0.0119 | 0.0198 | 8.7E-02 |
| *SSR1-RREB1* | rs9379084 | 6 | 7,231,843 | G | A | 0.0851 | 0.0079 | 6.1E-30 | 0.0594 | 0.0328 | 1.8E-01 |
| *CDKAL1* | rs9348441 | 6 | 20,680,678 | A | T | 0.1541 | 0.0053 | 6.2E-235 | 0.1243 | 0.0219 | 1.6E-14 |
| MHC region | rs879882 | 6 | 31,139,452 | C | T | 0.0507 | 0.0051 | 5.0E-26 | 0.0425 | 0.0208 | 2.3E-02 |
| *ZFAND3* | rs34247110 | 6 | 39,282,371 | A | G | 0.0429 | 0.0049 | 3.4E-21 | 0.0403 | 0.0200 | 1.8E-01 |
| *LRFN2-KCNK16* | rs34298980 | 6 | 40,409,243 | T | C | 0.0305 | 0.0053 | 4.1E-09 | 0.0309 | 0.0207 | 3.9E-01 |
| *VEGFA* | rs6458354 | 6 | 43,814,190 | C | T | 0.0412 | 0.0059 | 1.3E-13 | 0.0487 | 0.0225 | 2.4E-01 |
| *TFAP2B* | rs3798519 | 6 | 50,788,778 | C | A | 0.0563 | 0.0059 | 2.4E-22 | 0.0367 | 0.0243 | 5.0E-01 |
| *BEND3* | rs1665901 | 6 | 107,433,400 | A | T | 0.0362 | 0.0054 | 6.1E-12 | -0.0153 | 0.0213 | 8.2E-01 |
| *NUS1* | rs72951506 | 6 | 118,011,723 | C | T | 0.0427 | 0.0065 | 9.9E-11 | 0.0400 | 0.0265 | 5.6E-01 |
| *CENPW-SOGA3* | rs11759026 | 6 | 126,792,095 | G | A | 0.0684 | 0.0058 | 1.0E-36 | 0.0466 | 0.0239 | 9.8E-02 |
| *MED23* | rs7739842 | 6 | 131,954,797 | G | T | 0.0374 | 0.0055 | 1.8E-13 | 0.0563 | 0.0233 | 1.2E-01 |
| *SLC35D3* | rs6937795 | 6 | 137,291,281 | A | C | 0.0354 | 0.0049 | 5.3E-15 | 0.0471 | 0.0199 | 1.4E-01 |
| *REPS1* | rs9376353 | 6 | 138,855,975 | A | T | 0.0306 | 0.0050 | 2.7E-10 | 0.0315 | 0.0202 | 4.7E-01 |
| *HIVEP2* | rs6570526 | 6 | 143,058,692 | G | C | 0.0310 | 0.0049 | 8.2E-10 | 0.0162 | 0.0199 | 9.7E-03 |
| *RGS17* | rs6932473 | 6 | 153,438,573 | T | A | 0.0365 | 0.0051 | 5.5E-13 | 0.0830 | 0.0207 | 1.6E-03 |
| *SLC22A3* | rs539298 | 6 | 160,770,360 | A | G | 0.0386 | 0.0049 | 1.4E-15 | 0.0207 | 0.0200 | 8.5E-01 |
| *QKI* | rs4709746 | 6 | 164,133,001 | C | T | 0.0475 | 0.0076 | 7.8E-10 | 0.0658 | 0.0313 | 2.1E-01 |
| *ETV1* | rs12154701 | 7 | 13,887,008 | A | C | 0.0317 | 0.0049 | 1.4E-11 | 0.0426 | 0.0205 | 3.1E-01 |
| *DGKB* | rs2215383 | 7 | 15,062,983 | C | T | 0.0657 | 0.0049 | 2.1E-44 | 0.0374 | 0.0200 | 8.0E-02 |
| *JAZF1* | rs849133 | 7 | 28,192,280 | C | T | 0.0830 | 0.0051 | 2.3E-69 | 0.0625 | 0.0202 | 4.2E-02 |
| *CRHR2* | rs917195 | 7 | 30,728,452 | C | T | 0.0486 | 0.0060 | 3.6E-16 | 0.0614 | 0.0244 | 8.1E-02 |
| *GCK* | rs878521 | 7 | 44,255,643 | A | G | 0.0471 | 0.0055 | 2.2E-20 | 0.0690 | 0.0231 | 4.8E-02 |
| *GRB10* | rs13236710 | 7 | 50,809,085 | G | A | 0.0491 | 0.0076 | 2.0E-11 | 0.0261 | 0.0309 | 3.4E-01 |
| *AUTS2* | rs2533457 | 7 | 69,055,951 | G | A | 0.0409 | 0.0051 | 3.3E-19 | 0.0409 | 0.0204 | 2.4E-01 |
| *STEAP1* | rs6978118 | 7 | 89,800,241 | A | T | 0.0335 | 0.0051 | 2.0E-14 | 0.0630 | 0.0203 | 2.8E-02 |
| *FBXL13-RELN* | rs7781557 | 7 | 102,481,891 | C | T | 0.0527 | 0.0077 | 8.9E-12 | 0.0216 | 0.0285 | 1.9E-01 |

| | | | | | | | | | | | |
|---|---|---|---|---|---|---|---|---|---|---|---|
| *GCC1-PAX4-LEP* | rs12669223 | 7 | 127,250,831 | A | G | 0.1912 | 0.0156 | 2.4E-39 | N/A | N/A | N/A |
| *KLF14* | rs1562396 | 7 | 130,457,914 | G | A | 0.0385 | 0.0051 | 1.7E-16 | 0.0124 | 0.0215 | 7.3E-01 |
| *BRAF* | rs11983228 | 7 | 140,631,823 | C | G | 0.0523 | 0.0083 | 5.0E-10 | 0.0816 | 0.0368 | 1.2E-01 |
| *AOC1* | rs62492368 | 7 | 150,537,635 | A | G | 0.0315 | 0.0050 | 1.5E-10 | 0.0341 | 0.0212 | 3.5E-01 |
| *MNX1* | rs10085650 | 7 | 156,993,413 | T | C | 0.0512 | 0.0050 | 3.9E-26 | 0.0507 | 0.0210 | 8.7E-02 |
| *MSRA-XKR6* | rs4240673 | 8 | 10,787,612 | T | C | 0.0382 | 0.0058 | 1.1E-11 | -0.0260 | 0.0220 | 1.1E-01 |
| *LONRF1* | rs12680692 | 8 | 12,618,225 | A | T | 0.0324 | 0.0057 | 6.3E-10 | 0.0086 | 0.0224 | 2.0E-01 |
| *LPL* | rs7819706 | 8 | 19,844,415 | A | G | 0.0510 | 0.0076 | 4.3E-13 | 0.0437 | 0.0330 | 4.8E-01 |
| *KCNU1* | rs12680217 | 8 | 37,397,803 | T | C | 0.0498 | 0.0068 | 5.2E-15 | 0.0128 | 0.0321 | 8.5E-01 |
| *ANK1* | rs508419 | 8 | 41,522,991 | G | A | 0.0823 | 0.0062 | 5.7E-47 | 0.0031 | 0.0242 | 9.1E-01 |
| *GDAP1* | rs3780012 | 8 | 75,147,209 | C | G | 0.1320 | 0.0218 | 8.0E-10 | N/A | N/A | N/A |
| *TP53INP1* | rs13257021 | 8 | 95,965,695 | A | G | 0.0428 | 0.0049 | 3.3E-20 | 0.0381 | 0.0200 | 3.2E-01 |
| *TRPS1* | rs800909 | 8 | 116,497,173 | T | C | 0.0335 | 0.0053 | 8.1E-12 | 0.0034 | 0.0204 | 6.6E-01 |
| *SLC30A8* | rs13266634 | 8 | 118,184,783 | C | T | 0.1124 | 0.0053 | 3.2E-115 | 0.0615 | 0.0210 | 2.9E-02 |
| *PVT1* | rs4733612 | 8 | 129,569,999 | G | A | 0.0423 | 0.0063 | 2.4E-12 | 0.0717 | 0.0234 | 1.9E-03 |
| *BOP1* | rs3890400 | 8 | 145,544,720 | A | G | 0.0458 | 0.0054 | 3.3E-18 | 0.0145 | 0.0211 | 1.7E-01 |
| *GLIS3* | rs4237150 | 9 | 4,290,085 | C | G | 0.0566 | 0.0049 | 1.5E-36 | 0.0619 | 0.0204 | 8.1E-03 |
| *HAUS6* | rs12380322 | 9 | 19,074,538 | G | A | 0.0315 | 0.0051 | 4.9E-10 | 0.0258 | 0.0205 | 3.7E-01 |
| *CDKN2A-CDKN2B* | rs10811661 | 9 | 22,134,094 | T | C | 0.1694 | 0.0060 | 1.1E-201 | 0.1599 | 0.0261 | 4.6E-09 |
| *LINGO2* | rs1412234 | 9 | 28,410,683 | C | T | 0.0342 | 0.0056 | 1.5E-11 | 0.0377 | 0.0217 | 7.0E-02 |
| *UBAP2* | rs12001437 | 9 | 34,074,476 | C | T | 0.0315 | 0.0050 | 5.6E-11 | 0.0459 | 0.0204 | 1.5E-01 |
| *TLE4* | rs13290396 | 9 | 81,914,978 | C | T | 0.0968 | 0.0094 | 1.4E-26 | 0.0368 | 0.0363 | 6.2E-01 |
| *TLE1* | rs2796441 | 9 | 84,308,948 | G | A | 0.0645 | 0.0050 | 8.0E-42 | 0.0435 | 0.0201 | 1.6E-01 |
| *ZNF169* | rs12345069 | 9 | 96,971,175 | C | T | 0.0382 | 0.0062 | 4.4E-10 | 0.0339 | 0.0236 | 4.8E-01 |
| *PTCH1* | rs113154802 | 9 | 98,278,413 | C | T | 0.0511 | 0.0084 | 4.3E-12 | 0.0468 | 0.0340 | 7.2E-01 |
| *STRBP* | rs2416899 | 9 | 126,015,103 | T | G | 0.0344 | 0.0063 | 3.5E-10 | 0.0296 | 0.0268 | 4.5E-01 |
| *ABO* | rs505922 | 9 | 136,149,229 | C | T | 0.0457 | 0.0050 | 2.0E-21 | 0.0271 | 0.0214 | 2.7E-02 |
| *GPSM1* | rs28429551 | 9 | 139,243,334 | A | T | 0.0842 | 0.0069 | 1.4E-39 | 0.0703 | 0.0261 | 1.8E-03 |
| *CDC123-CAMK1D* | rs11257655 | 10 | 12,307,894 | T | C | 0.1025 | 0.0055 | 1.2E-91 | 0.0934 | 0.0227 | 8.2E-04 |
| *MYO3A* | rs7923442 | 10 | 26,497,704 | A | G | 0.0354 | 0.0058 | 1.2E-09 | 0.0300 | 0.0254 | 8.5E-03 |
| *JMJD1C* | rs41274074 | 10 | 64,974,380 | G | C | 0.0600 | 0.0092 | 3.1E-11 | -0.0302 | 0.0472 | 8.3E-01 |
| *VPS26A-NEUROG3* | rs177045 | 10 | 71,321,279 | G | A | 0.0450 | 0.0051 | 5.0E-23 | 0.0611 | 0.0211 | 4.3E-03 |
| *ZNF503* | rs3012060 | 10 | 77,244,336 | T | A | 0.0450 | 0.0068 | 1.6E-12 | 0.0986 | 0.0294 | 1.8E-02 |
| *ZMIZ1* | rs703980 | 10 | 80,943,841 | G | A | 0.0620 | 0.0049 | 8.7E-40 | 0.0507 | 0.0199 | 3.6E-02 |
| *PTEN* | rs10887775 | 10 | 89,766,368 | A | G | 0.0388 | 0.0060 | 8.9E-11 | 0.0646 | 0.0265 | 4.0E-02 |

| | | | | | | | | | | | |
|---|---|---|---|---|---|---|---|---|---|---|---|
| *HHEX-IDE* | rs10882101 | 10 | 94,462,427 | T | C | 0.1091 | 0.0050 | 1.8E-125 | 0.0717 | 0.0201 | 1.8E-03 |
| *ARHGAP19-SLIT1* | rs10748694 | 10 | 99,056,190 | A | T | 0.0394 | 0.0050 | 5.1E-17 | 0.0357 | 0.0202 | 4.5E-01 |
| *BBIP1* | rs7067540 | 10 | 112,621,837 | C | T | 0.0397 | 0.0053 | 1.1E-14 | -0.0029 | 0.0221 | 9.5E-01 |
| *TCF7L2* | rs7903146 | 10 | 114,758,349 | T | C | 0.2983 | 0.0062 | 0.0E+00 | 0.1961 | 0.0230 | 4.0E-16 |
| *WDR11* | rs72631105 | 10 | 122,915,345 | A | G | 0.0504 | 0.0062 | 1.9E-18 | 0.0347 | 0.0243 | 5.9E-02 |
| *PLEKHA1* | rs2421016 | 10 | 124,167,512 | C | T | 0.0425 | 0.0049 | 9.6E-23 | 0.0144 | 0.0199 | 5.7E-01 |
| *DUSP8-INS-IGF2-KCNQ1* | rs2237897 | 11 | 2,858,546 | C | T | 0.2390 | 0.0081 | 5.5E-233 | 0.1721 | 0.0387 | 2.2E-04 |
| *TRIM66* | rs10769936 | 11 | 8,654,528 | C | T | 0.0322 | 0.0051 | 6.9E-11 | 0.0388 | 0.0219 | 4.0E-01 |
| *KCNJ11-ABCC8* | rs5215 | 11 | 17,408,630 | C | T | 0.0743 | 0.0050 | 1.3E-54 | 0.0560 | 0.0202 | 9.0E-03 |
| *BDNF* | rs4923464 | 11 | 27,683,618 | C | T | 0.0317 | 0.0057 | 9.4E-10 | 0.0050 | 0.0243 | 7.7E-01 |
| *QSER1* | rs145678014 | 11 | 32,927,778 | G | T | 0.0968 | 0.0166 | 5.7E-10 | N/A | N/A | N/A |
| *HSD17B12* | rs6485462 | 11 | 43,816,200 | C | T | 0.0302 | 0.0053 | 4.8E-12 | 0.0576 | 0.0212 | 7.0E-02 |
| *CRY2* | rs12419690 | 11 | 45,858,584 | G | A | 0.0343 | 0.0051 | 4.2E-11 | 0.0688 | 0.0201 | 8.9E-03 |
| *FOLH1* | rs6485981 | 11 | 49,477,266 | T | C | 0.0410 | 0.0075 | 1.5E-09 | -0.0024 | 0.0344 | 2.4E-01 |
| *MAP3K11* | rs12789028 | 11 | 65,326,154 | A | G | 0.0544 | 0.0066 | 2.1E-17 | 0.0453 | 0.0272 | 4.8E-01 |
| *TPCN2-CCND1* | rs3918298 | 11 | 69,463,273 | G | A | 0.1106 | 0.0138 | 3.3E-17 | N/A | N/A | N/A |
| *CENTD2* | rs77464186 | 11 | 72,460,398 | A | C | 0.1039 | 0.0075 | 3.6E-49 | 0.1119 | 0.0271 | 6.0E-06 |
| *C11orf30* | rs61894507 | 11 | 76,156,973 | G | A | 0.0352 | 0.0057 | 2.2E-10 | -0.0102 | 0.0219 | 5.3E-01 |
| *MTNR1B* | rs10830963 | 11 | 92,708,710 | G | C | 0.0840 | 0.0054 | 6.1E-66 | 0.3404 | 0.0218 | 4.3E-54 |
| *ETS1* | rs11819995 | 11 | 128,389,391 | T | C | 0.0445 | 0.0059 | 2.5E-14 | 0.0379 | 0.0246 | 1.9E-01 |
| *CCND2* | rs76895963 | 12 | 4,384,844 | T | G | 0.4575 | 0.0280 | 3.7E-71 | N/A | N/A | N/A |
| *CDKN1B* | rs2066827 | 12 | 12,871,099 | G | T | 0.0467 | 0.0071 | 7.1E-11 | 0.0477 | 0.0258 | 1.4E-01 |
| *ITPR2* | rs10842708 | 12 | 26,474,867 | G | A | 0.0385 | 0.0055 | 5.5E-14 | -0.0035 | 0.0222 | 9.1E-01 |
| *KLHDC5* | rs12578595 | 12 | 27,964,996 | C | T | 0.0663 | 0.0058 | 7.8E-33 | 0.0566 | 0.0247 | 1.5E-01 |
| *FAM60A* | rs78345706 | 12 | 31,417,019 | A | G | 0.1157 | 0.0116 | 1.2E-24 | N/A | N/A | N/A |
| *PKP2-SYT10* | rs6488140 | 12 | 33,370,406 | A | G | 0.0354 | 0.0060 | 9.1E-12 | -0.0377 | 0.0269 | 1.1E-01 |
| *FAIM2* | rs7132908 | 12 | 50,263,148 | A | G | 0.0319 | 0.0053 | 6.5E-10 | 0.0532 | 0.0207 | 7.5E-02 |
| *HMGA2* | rs2583930 | 12 | 66,246,181 | G | A | 0.0708 | 0.0059 | 5.1E-38 | 0.0444 | 0.0256 | 1.4E-01 |
| *TSPAN8* | rs7313668 | 12 | 71,449,521 | T | G | 0.0413 | 0.0051 | 2.7E-17 | 0.0266 | 0.0204 | 6.2E-01 |
| *RMST* | rs7972074 | 12 | 97,851,611 | C | T | 0.0347 | 0.0058 | 1.1E-10 | 0.0327 | 0.0268 | 7.0E-01 |
| *WSCD2* | rs1426371 | 12 | 108,629,780 | G | A | 0.0479 | 0.0055 | 9.7E-19 | 0.0140 | 0.0227 | 6.4E-01 |
| *ALDH2-BRAP* | rs3782886 | 12 | 112,110,489 | T | C | 0.0597 | 0.0116 | 1.1E-13 | N/A | N/A | N/A |
| *PTPN11* | rs77753011 | 12 | 113,117,897 | G | T | 0.0769 | 0.0137 | 4.0E-15 | N/A | N/A | N/A |
| *KSR2* | rs34965774 | 12 | 118,412,373 | A | G | 0.0581 | 0.0063 | 1.5E-21 | 0.0496 | 0.0265 | 1.9E-01 |
| *HNF1A* | rs1169299 | 12 | 121,429,194 | C | T | 0.0219 | 0.0050 | 1.9E-21 | 0.0656 | 0.0200 | 1.1E-04 |

| | | | | | | | | | | | |
|---|---|---|---|---|---|---|---|---|---|---|---|
| *MPHOSPH9-ZNF664* | rs1790116 | 12 | 123,618,544 | T | G | 0.0433 | 0.0067 | 2.2E-11 | -0.0022 | 0.0250 | 9.2E-01 |
| *FBRSL1* | rs12811407 | 12 | 133,069,698 | A | G | 0.0462 | 0.0059 | 2.0E-15 | 0.0799 | 0.0223 | 5.5E-03 |
| *SGCG* | rs314879 | 13 | 23,309,382 | C | T | 0.0405 | 0.0062 | 3.7E-11 | 0.0101 | 0.0258 | 9.7E-01 |
| *RNF6* | rs34584161 | 13 | 26,776,999 | A | G | 0.0471 | 0.0055 | 3.5E-19 | 0.0386 | 0.0228 | 2.1E-03 |
| *KL* | rs2858980 | 13 | 33,554,587 | G | A | 0.0570 | 0.0063 | 6.2E-22 | 0.0820 | 0.0262 | 1.8E-02 |
| *DLEU1* | rs963740 | 13 | 51,096,095 | A | T | 0.0355 | 0.0053 | 3.8E-11 | 0.0448 | 0.0220 | 7.7E-02 |
| *OLFM4* | rs9568868 | 13 | 54,107,583 | T | G | 0.0467 | 0.0065 | 1.5E-12 | 0.0384 | 0.0275 | 2.1E-01 |
| *SPRY2* | rs1215468 | 13 | 80,707,429 | A | G | 0.0812 | 0.0055 | 2.4E-56 | 0.0402 | 0.0222 | 2.7E-01 |
| *MIR17HG* | rs34165267 | 13 | 91,942,919 | C | T | 0.0495 | 0.0059 | 6.7E-21 | 0.0383 | 0.0240 | 3.8E-01 |
| *AKAP6* | rs12883788 | 14 | 33,303,540 | T | C | 0.0348 | 0.0051 | 1.9E-11 | 0.0116 | 0.0205 | 1.4E-01 |
| *CLEC14A* | rs2183237 | 14 | 38,803,756 | G | A | 0.0401 | 0.0053 | 5.0E-15 | 0.0224 | 0.0217 | 3.0E-01 |
| *NRXN3* | rs8008910 | 14 | 79,944,099 | A | G | 0.0544 | 0.0069 | 3.3E-15 | -0.0135 | 0.0258 | 4.0E-01 |
| *MEG3* | rs73347525 | 14 | 101,255,172 | A | G | 0.0510 | 0.0065 | 1.8E-15 | 0.0088 | 0.0262 | 7.8E-01 |
| *MARK3* | rs11160699 | 14 | 103,252,270 | A | G | 0.0413 | 0.0059 | 5.9E-12 | 0.0188 | 0.0260 | 5.7E-01 |
| *RASGRP1* | rs12912777 | 15 | 38,852,386 | T | C | 0.0787 | 0.0099 | 2.7E-16 | 0.1142 | 0.0354 | 4.9E-03 |
| *INFAM2-LTK* | rs3743140 | 15 | 40,616,742 | A | G | 0.0489 | 0.0068 | 5.0E-16 | 0.0861 | 0.0293 | 4.0E-02 |
| *LTK* | rs1473781 | 15 | 41,818,917 | A | G | 0.0327 | 0.0056 | 2.1E-11 | 0.0275 | 0.0214 | 1.8E-01 |
| *ONECUT1-WDR72* | rs3825801 | 15 | 52,517,714 | C | T | 0.0454 | 0.0072 | 3.8E-11 | 0.0712 | 0.0309 | 1.5E-01 |
| *C2CD4A-C2CD4B* | rs7163757 | 15 | 62,391,608 | C | T | 0.0571 | 0.0049 | 2.4E-37 | 0.0099 | 0.0200 | 7.9E-01 |
| *USP3* | rs7178762 | 15 | 63,871,292 | C | T | 0.0379 | 0.0055 | 9.2E-12 | 0.0218 | 0.0209 | 1.1E-01 |
| *MAP2K5* | rs4776970 | 15 | 68,080,886 | A | T | 0.0360 | 0.0053 | 5.7E-13 | 0.0486 | 0.0208 | 2.0E-03 |
| *PTPN9* | rs11636031 | 15 | 75,815,758 | T | C | 0.0465 | 0.0054 | 4.9E-19 | 0.0115 | 0.0228 | 3.2E-02 |
| *HMG20A* | rs952472 | 15 | 77,776,562 | C | A | 0.0761 | 0.0051 | 4.1E-56 | 0.0071 | 0.0211 | 9.1E-01 |
| *AP3S2* | rs6496609 | 15 | 90,379,632 | C | A | 0.0636 | 0.0056 | 3.2E-34 | 0.0877 | 0.0221 | 8.8E-04 |
| *PRC1* | rs2890156 | 15 | 91,513,157 | A | T | 0.0690 | 0.0059 | 3.8E-34 | 0.0732 | 0.0253 | 1.8E-02 |
| *RGMA* | rs7167984 | 15 | 93,832,067 | G | A | 0.0440 | 0.0063 | 3.4E-13 | 0.0242 | 0.0276 | 3.2E-01 |
| *ITFG3* | rs6600191 | 16 | 295,795 | T | C | 0.0439 | 0.0057 | 2.4E-15 | 0.0526 | 0.0260 | 5.9E-02 |
| *CLUAP1* | rs12445430 | 16 | 3,613,126 | T | C | 0.0396 | 0.0058 | 2.5E-11 | 0.0295 | 0.0281 | 1.3E-01 |
| *FAM57B* | rs11642430 | 16 | 30,045,789 | G | C | 0.0290 | 0.0050 | 6.6E-10 | 0.0359 | 0.0201 | 3.6E-01 |
| *FTO* | rs55872725 | 16 | 53,809,123 | T | C | 0.1215 | 0.0055 | 4.7E-128 | 0.0554 | 0.0207 | 5.5E-02 |
| *NFAT5* | rs862320 | 16 | 69,651,866 | C | T | 0.0347 | 0.0055 | 1.5E-10 | 0.0194 | 0.0207 | 8.5E-02 |
| *ZFHX3* | rs6416749 | 16 | 73,100,308 | C | T | 0.0375 | 0.0056 | 1.5E-13 | -0.0023 | 0.0223 | 5.0E-01 |
| *BCAR1* | rs72802358 | 16 | 75,243,657 | G | C | 0.0877 | 0.0090 | 2.9E-29 | 0.0825 | 0.0349 | 1.7E-01 |
| *CMIP* | rs2925979 | 16 | 81,534,790 | T | C | 0.0494 | 0.0054 | 1.6E-21 | 0.0282 | 0.0219 | 4.5E-01 |
| *ZFPM1* | rs9937296 | 16 | 88,554,480 | C | T | 0.0424 | 0.0071 | 5.3E-10 | 0.0287 | 0.0295 | 3.4E-01 |

| | | | | | | | | | | | |
|---|---|---|---|---|---|---|---|---|---|---|---|
| *SPG7* | rs12920022 | 16 | 89,564,055 | A | T | 0.0420 | 0.0074 | 9.9E-10 | 0.0295 | 0.0303 | 2.1E-01 |
| *ZZEF1* | rs8071043 | 17 | 3,988,451 | C | T | 0.0379 | 0.0055 | 4.3E-15 | 0.0260 | 0.0217 | 7.8E-01 |
| *SLC16A11-SLC16A13* | rs113748381 | 17 | 6,953,155 | A | G | 0.1078 | 0.0115 | 2.3E-24 | N/A | N/A | N/A |
| *RAI1* | rs1108646 | 17 | 17,751,478 | A | G | 0.0394 | 0.0056 | 2.8E-13 | 0.0150 | 0.0223 | 7.9E-01 |
| *NF1* | rs1048317 | 17 | 29,704,002 | T | C | 0.0378 | 0.005 | 1.4E-14 | 0.0178 | 0.0202 | 7.5E-01 |
| *HNF1B* | rs10908278 | 17 | 36,099,952 | T | A | 0.0836 | 0.0053 | 7.4E-74 | 0.0398 | 0.0221 | 1.7E-01 |
| *MLX* | rs684214 | 17 | 40,696,915 | T | C | 0.0397 | 0.0056 | 3.0E-13 | 0.0106 | 0.0227 | 7.6E-01 |
| *GIP-TTLL6* | rs35895680 | 17 | 47,060,322 | C | A | 0.0480 | 0.0064 | 2.3E-14 | 0.0019 | 0.0231 | 6.4E-01 |
| *ACE* | rs57676627 | 17 | 62,203,128 | T | C | 0.0542 | 0.0087 | 1.8E-10 | 0.0556 | 0.0340 | 2.9E-01 |
| *BPTF* | rs9899520 | 17 | 65,957,568 | A | G | 0.0380 | 0.0056 | 1.3E-14 | 0.0034 | 0.0241 | 6.0E-01 |
| *CYTH1* | rs1044486 | 17 | 76,792,179 | G | A | 0.0357 | 0.0050 | 3.2E-13 | 0.0372 | 0.0199 | 1.8E-01 |
| *LAMA1* | rs9948462 | 18 | 7,076,836 | T | C | 0.0371 | 0.0051 | 3.9E-15 | -0.0110 | 0.0205 | 5.6E-01 |
| *TCF4* | rs72926932 | 18 | 53,050,646 | C | A | 0.0712 | 0.0116 | 1.2E-09 | 0.0688 | 0.0396 | 3.9E-01 |
| *GRP-MC4R* | rs6567160 | 18 | 57,829,135 | C | T | 0.0690 | 0.0058 | 1.1E-37 | 0.0657 | 0.0233 | 7.7E-02 |
| *BCL2A* | rs12454712 | 18 | 60,845,884 | T | C | 0.0461 | 0.0051 | 4.1E-20 | -0.0215 | 0.0216 | 3.1E-02 |
| *ZNF236* | rs12457906 | 18 | 74,555,593 | G | A | 0.0352 | 0.0049 | 6.0E-13 | 0.0410 | 0.0205 | 7.8E-02 |
| *UHRF1-PTPRS* | rs262549 | 19 | 4,951,064 | G | C | 0.0447 | 0.0073 | 2.5E-09 | 0.0780 | 0.0272 | 5.0E-03 |
| *MAP2K7* | rs2115107 | 19 | 7,968,168 | A | G | 0.0444 | 0.0051 | 1.1E-18 | 0.0550 | 0.0210 | 7.3E-02 |
| *FARSA* | rs3111316 | 19 | 13,038,415 | A | G | 0.0386 | 0.0053 | 1.3E-13 | 0.0126 | 0.0213 | 9.0E-01 |
| *CILP2-TM6SF2* | rs58542926 | 19 | 19,379,549 | T | C | 0.0657 | 0.0095 | 1.6E-13 | 0.0473 | 0.0376 | 4.5E-01 |
| *ZNF257* | rs142395395 | 19 | 22,100,706 | A | G | 0.2070 | 0.0263 | 1.2E-15 | N/A | N/A | N/A |
| *PEPD* | rs10406327 | 19 | 33,890,838 | C | G | 0.0426 | 0.0049 | 3.5E-20 | 0.0365 | 0.0199 | 2.7E-01 |
| *TOMM40-APOE-GIPR* | rs10406431 | 19 | 46,157,019 | A | G | 0.0656 | 0.0051 | 8.7E-44 | 0.0565 | 0.0212 | 8.5E-02 |
| *ZC3H4* | rs3810291 | 19 | 47,569,003 | A | G | 0.0462 | 0.0054 | 8.6E-19 | 0.0276 | 0.0217 | 2.3E-01 |
| *FOXA2* | rs2181063 | 20 | 22,427,370 | C | G | 0.0343 | 0.0056 | 5.9E-10 | 0.0179 | 0.0241 | 8.2E-02 |
| *RALY* | rs4911405 | 20 | 32,674,967 | T | C | 0.0359 | 0.0057 | 4.4E-12 | 0.0362 | 0.0220 | 2.4E-03 |
| *HNF4A* | rs12625671 | 20 | 42,994,812 | C | T | 0.0779 | 0.0062 | 9.5E-40 | 0.0705 | 0.0278 | 1.1E-01 |
| *EYA2* | rs6063046 | 20 | 45,596,378 | A | G | 0.0437 | 0.0067 | 1.5E-10 | 0.0283 | 0.0237 | 3.4E-01 |
| *CEBPB* | rs6091115 | 20 | 48,832,020 | T | C | 0.0473 | 0.0049 | 9.3E-23 | 0.0217 | 0.0197 | 5.3E-01 |
| *GNAS* | rs736266 | 20 | 57,387,352 | T | A | 0.0304 | 0.0049 | 6.6E-10 | 0.0453 | 0.0200 | 9.8E-02 |
| *MTMR3* | rs36575 | 22 | 30,205,572 | C | T | 0.0790 | 0.0107 | 1.9E-13 | 0.1056 | 0.0404 | 6.1E-03 |
| *YWHAH* | rs75307421 | 22 | 32,203,334 | A | G | 0.0960 | 0.0158 | 3.1E-11 | N/A | N/A | N/A |
| *PNPLA3* | rs738408 | 22 | 44,324,730 | T | C | 0.0310 | 0.0055 | 5.5E-10 | -0.0023 | 0.0232 | 6.6E-01 |
| *PIM3* | rs28691713 | 22 | 50,356,302 | C | T | 0.0484 | 0.0054 | 5.9E-22 | 0.0302 | 0.0217 | 6.5E-01 |

### Table S6. Genetic correlation ($r_G$) from LD-score regression between GDM, T2D and glycaemic traits.

| $r_G$ (95% CI)[a] | GDM | T2D | Fasting glucose | Fasting insulin | Fasting proinsulin | 2hr glucose (BMI adjusted) | HbA1c | HOMA-B | HOMA-IR |
|---|---|---|---|---|---|---|---|---|---|
| **GDM** | | (0.05, 1.44) | (-0.21, 0.65) | (-0.12, 0.93) | (-0.32, 0.99) | (-0.37, 1.26) | (-0.22, 0.99) | (-0.55, 0.54) | (-0.38, 0.85) |
| **T2D** | 0.74 | | (0.47, 0.87) | (0.29, 0.66) | (-0.02, 0.48) | (0.11, 0.59) | (0.41, 0.59) | (-0.19, 0.21) | (0.25, 0.73) |
| **Fasting glucose** | 0.22 | 0.67 | | (0.12, 0.48) | (0.08, 0.64) | (-0.19, 0.35) | (0.21, 0.59) | (-0.67, -0.27) | (0.33, 0.67) |
| **Fasting insulin** | 0.41 | 0.48 | 0.30 | | (-0.02, 0.52) | (-0.29, 0.29) | (-0.04, 0.38) | (0.95, 1.25) | (1.32, 1.74) |
| **Fasting proinsulin** | 0.34 | 0.23 | 0.36 | 0.25 | | (-0.39, 0.37) | (-0.24, 0.36) | (-0.32, 0.30) | (-0.02, 0.54) |
| **2hr glucose (BMI adjusted)** | 0.44 | 0.35 | 0.08 | 0.00 | -0.01 | | (-0.20, 0.34) | (-0.41, 0.23) | (-0.45, 0.25) |
| **HbA1c** | 0.39 | 0.59 | 0.40 | 0.17 | 0.06 | 0.07 | | (-0.43, 0.03) | (-0.04, 0.46) |
| **HOMA-B** | 0.00 | 0.01 | -0.47 | 1.10 | -0.01 | -0.09 | -0.20 | | (0.59, 0.83) |
| **HOMA-IR** | 0.24 | 0.49 | 0.50 | 1.53 | 0.26 | -0.10 | 0.21 | 0.71 | |

CI: Confidence Interval. T2D: type 2 diabetes. HbA1c: glycated haemoglobin. BMI: body mass index. HOMA-B: homeostasis model assessment of β-cell activity; HOMA-IR: homeostasis model assessment of insulin resistance.

[a]Genetic correlation obtained from LD-score regression is not bound by -1 to 1 and estimates can therefore be found outside these limits due to high imprecision caused by factors such as low sample size in the GWAS summary statistics used.

**Table S7. Impact of lead SNVs at GDM loci on T2D and glycaemic traits.**

| Locus | Lead SNV | Alleles | | GDM[a] | | T2D[b] | | Fasting glucose[c] | | HbA1c[d] | |
|---|---|---|---|---|---|---|---|---|---|---|---|
| | | Risk | Other | Log-OR (SE) | *p*-value | Log-OR (SE) | *p*-value | Beta (SE) | *p*-value | Beta (SE) | *p*-value |
| *MTNR1B* | rs10830963 | G | C | 0.3404 (0.0218) | 4.3E-54 | 0.0840 (0.0054) | 6.1E-66 | 0.0772 (0.0019) | 1.4E-321 | 0.0197 (0.0015) | 1.5E-36 |
| *TCF7L2* | rs7903146 | T | C | 0.1961 (0.0230) | 4.0E-16 | 0.2983 (0.0062) | 0 | 0.0259 (0.0019) | 2.0E-35 | 0.0133 (0.0014) | 1.0E-22 |
| *CDKAL1* | rs9348441 | A | T | 0.1243 (0.0291) | 1.6E-14 | 0.1541 (0.0053) | 6.2E-235 | 0.0176 (0.0018) | 4.4E-20 | 0.0101 (0.0014) | 2.4E-13 |
| *CDKN2A-CDKN2B* | rs10811662 | G | A | 0.1329 (0.0255) | 4.1E-9 | 0.1661 (0.0060) | 3.5E-166 | 0.0223 (0.0022) | 7.9E-25 | 0.0127 (0.0017) | 3.2E-14 |
| *HKDC1* | rs9663238 | G | A | 0.1339 (0.0222) | 2.9E-8 | 0.0139 (0.0054) | 8.3E-3 | -0.0080 (0.0019) | 2.1E-4 | 0.0037 (0.0014) | 1.5E-2 |

OR: odds-ratio. SE: standard error

[a]Trans-ancestry meta-analysis of 5,485 cases and 347,856 controls from the current study.

[b]Trans-ancestry meta-analysis of 180,834 cases and 1,159,055 controls from the DIAGRAM Consortium. Mahajan A, Spracklen CN, Zhang W, Ng MC, Petty LE, Kitajima H, et al. Trans-ancestry genetic study of type 2 diabetes highlights the power of diverse populations for discovery and translation. medRxiv. 2020

[c]European ancestry meta-analysis of 200,622 non-diabetic individuals from the MAGIC Investigators. Chen J, Spracklen CN, Marenne G, Varshney A, Corbin LJ, Luan J, et al. The trans-ancestral genomic architecture of glycemic traits. Nat Genet 53: 840–60.

[d]European ancestry meta-analysis of 146,806 non-diabetic individuals from the MAGIC Investigators. Chen J, Spracklen CN, Marenne G, Varshney A, Corbin LJ, Luan J, et al. The trans-ancestral genomic architecture of glycemic traits. Nat Genet 53: 840–60.

**Table S8. Summary statistics from joint fGWAS model of enriched functional and regulatory annotations for GDM association signals from trans-ancestry meta-regression (MR-MEGA).**

| **Annotation**[a] | **Enrichment *p*-value** | **Fold-enrichment (95% CI)** |
|---|---|---|
| Adipose active enhancer 2 | 0.0019 | 2.84 (1.55-4.58) |
| FOXA2 binding site | 0.0031 | 4.63 (1.87-8.89) |
| Skeletal muscle weak enhancer | 0.0098 | 1.99 (1.19-3.05) |
| TFAP2A binding site | 0.014 | 4.99 (1.59-10.50) |
| NFE2 binding site | 0.021 | 10.49 (1.72-30.66) |
| Protein coding exon | 0.032 | 3.57 (1.13-7.45) |
| Adipose strong transcription | 0.037 | 1.79 (1.04-2.80) |

[a]Three categories of functional and regulatory annotations were considered: (i) genic regions; (ii) chromatin immuno-precipitation sequence (ChIP-seq) binding sites for 165 transcription factors; and (iii) 13 unique and recurrent chromatin states in four diabetes-relevant tissues (pancreatic islets, liver, adipose, and skeletal muscle). Details are provided in the **Supplementary Materials and Methods**.

**Table S9. Genome-wide significant associations ($p$<5x10$^{-8}$) of 99% credible set variants at the *HKDC1* locus with other traits.**

| SNV | Chr | Position (bp, b37) | Alleles | | Trait | beta | *p*-value | Sample size | Data source |
|---|---|---|---|---|---|---|---|---|---|
| | | | GDM risk | other | | | | | |
| rs10998647 | 10 | 70,977,308 | G | T | Lymphocyte count | -0.024 | 1.1x10$^{-9}$ | 173,480 | PMID: 27863252 |
| rs10998648 | 10 | 70,977,395 | C | A | Comparative height size at age 10 | 0.011 | 3.4x10$^{-9}$ | 332,021 | UK Biobank |
| rs10823318 | 10 | 70,979,924 | T | A | Birth weight | 0.030 | 2.9x10$^{-18}$ | 193,063 | UK Biobank |
| | | | | | Comparative body size at age 10 | 0.010 | 2.8x10$^{-8}$ | 331,693 | UK Biobank |
| rs4746822 | 10 | 70,982,941 | T | C | 2-hour plasma glucose in pregnancy | 0.280 | 1.0x10$^{-22}$ | N/A | PMID: 23903356 |
| rs35199395 | 10 | 70,983,936 | C | G | Birth weight of first child | 0.029 | 3.9x10$^{-9}$ | 145,558 | UK Biobank |

**Table S10. 99% credible set variants at the *HKDC1* locus that are lead *cis*-eQTL variants in samples from the Genotype Tissue Expression (GTEx) Project.**

| Gene | Tissue | Lead eQTL variant | Chr | Position (bp, b37) | Alleles | | Association with gene expression | |
|---|---|---|---|---|---|---|---|---|
| | | | | | GDM risk | Other | Beta | p-value |
| *HKDC1* | Adipose (subcutaneous) | rs4746822 | 10 | 70,982,941 | T | C | -0.28 | $1.3x10^{-8}$ |
| | Adipose (visceral) | rs9663238 | 10 | 70,983,629 | G | A | -0.32 | $1.9x10^{-8}$ |
| | Breast (mammary) | rs5030937 | 10 | 70,975,897 | T | C | -0.41 | $8.6x10^{-10}$ |
| | Esophagus mucosa | rs10762264 | 10 | 70,976,833 | A | G | -0.30 | $1.2x10^{-9}$ |
| | Whole blood | rs4746822 | 10 | 70,982,941 | T | C | -0.42 | $5.9x10^{-26}$ |
| *SUPV3L1* | Lung | rs5030937 | 10 | 70,975,897 | T | C | -0.18 | $1.9x10^{-9}$ |
| | Whole blood | rs5030937 | 10 | 70,975,897 | T | C | -0.21 | $1.1x10^{-22}$ |

### Table S11. Effect estimates of metabolic traits on GDM from MR analyses.

| Metabolic trait | Category | Subcategory | MR method | SNVs | Beta | SE | *p*-value | *q*-value | Sample size | Sex | PMID |
|---|---|---|---|---|---|---|---|---|---|---|---|
| Body mass index | Risk factor | Anthropometric | RE IVW | 36 | 0.688 | 0.112 | $4.8x10^{-7}$ | $6.8x10^{-5}$ | 171,977 | Women | 25673413 |
| Triglycerides | Metabolites | NA | FE IVW | 175 | 0.190 | 0.065 | 0.0035 | 0.25 | NA | Both | NA |
| X-14626 | Metabolites | Unknown metabolite | RE IVW | 2 | -1.416 | 0.010 | 0.0043 | 0.25 | 6,904 | Both | 24816252 |
| Obesity class 1 | Risk factor | Anthropometric | Weighted median | 17 | 0.231 | 0.084 | 0.0059 | 0.25 | 98,697 | Both | 23563607 |
| Alanine | Metabolites | Amino acid | Weighted median | 6 | -0.492 | 0.182 | 0.0070 | 0.25 | 24,796 | Both | 27005778 |
| X-13215 | Metabolites | Unknown metabolite | RE IVW | 2 | 0.660 | 0.009 | 0.0090 | 0.25 | 6,305 | Both | 24816252 |
| X-13429 | Metabolites | Unknown metabolite | RE IVW | 3 | -0.507 | 0.049 | 0.0093 | 0.25 | 6,344 | Both | 24816252 |
| Uridine | Metabolites | Nucleotide | RE IVW | 3 | 2.706 | 0.263 | 0.0093 | 0.25 | 7,800 | Both | 24816252 |
| Childhood obesity | Risk factor | Anthropometric | RE IVW | 5 | 0.264 | 0.057 | 0.0095 | 0.25 | 13,848 | Both | 22484627 |
| Waist-to-hip ratio | Risk factor | Anthropometric | RE IVW | 21 | 0.533 | 0.187 | 0.0098 | 0.25 | 118,003 | Women | 25673412 |
| N2,N2-dimethylguanosine | Metabolites | Nucleotide | RE IVW | 2 | 1.443 | 0.027 | 0.012 | 0.27 | 5,228 | Both | 24816252 |
| X-12456 | Metabolites | Unknown metabolite | RE IVW | 2 | -0.676 | 0.013 | 0.012 | 0.27 | 4,774 | Both | 24816252 |
| Overweight | Risk factor | Anthropometric | Penalised median | 14 | 0.296 | 0.122 | 0.015 | 0.31 | 158,855 | Both | 23563607 |
| Extreme body mass index | Risk factor | Anthropometric | Penalised median | 7 | 0.131 | 0.057 | 0.021 | 0.39 | 16,068 | Both | 23563607 |
| Hip circumference | Risk factor | Anthropometric | Simple median | 17 | 0.585 | 0.259 | 0.024 | 0.40 | 127,997 | Women | 25673412 |
| Waist circumference | Risk factor | Anthropometric | Simple median | 17 | 0.585 | 0.259 | 0.024 | 0.40 | 127,997 | Women | 25673412 |
| Triglycerides in large VLDL | Metabolites | Lipid | Simple median | 8 | -0.305 | 0.143 | 0.033 | 0.52 | 21,239 | Both | 27005778 |
| Triglycerides in very large VLDL | Metabolites | Lipid | Simple median | 7 | -0.353 | 0.170 | 0.038 | 0.57 | 21,548 | Both | 27005778 |
| Tetradecanedioate | Metabolites | Lipid | RE IVW | 3 | -0.773 | 0.173 | 0.047 | 0.66 | 6,046 | Both | 24816252 |
| Gamma glutamyltransferase | Metabolites | NA | Simple mean | 221 | 0.154 | 0.079 | 0.052 | 0.66 | NA | Both | NA |
| X-12798 | Metabolites | Unknown metabolite | RE IVW | 3 | -0.217 | 0.052 | 0.054 | 0.66 | 7,552 | Both | 24816252 |
| Hexadecanedioate | Metabolites | Lipid | RE IVW | 3 | -0.919 | 0.227 | 0.056 | 0.66 | 6,887 | Both | 24816252 |
| X-12510--2-aminooctanoic acid | Metabolites | Amino acid | RE IVW | 3 | 0.108 | 0.027 | 0.056 | 0.66 | 7,566 | Both | 24816252 |
| Total lipids in large VLDL | Metabolites | Lipid | Simple median | 9 | -0.279 | 0.149 | 0.061 | 0.67 | 18,960 | Both | 27005778 |
| Hexanoylcarnitine | Metabolites | Lipid | RE IVW | 4 | 0.397 | 0.139 | 0.064 | 0.67 | 7,786 | Both | 24816252 |
| X-11491 | Metabolites | Unknown metabolite | RE IVW | 3 | -0.698 | 0.191 | 0.067 | 0.67 | 6,584 | Both | 24816252 |
| Indoleacetate | Metabolites | Amino acid | RE IVW | 2 | 1.178 | 0.128 | 0.069 | 0.67 | 7,618 | Both | 24816252 |
| Obesity class 2 | Risk factor | Anthropometric | Simple median | 11 | 0.125 | 0.070 | 0.073 | 0.67 | 72,546 | Both | 23563607 |
| X-10510 | Metabolites | Unknown metabolite | RE IVW | 3 | -1.075 | 0.308 | 0.073 | 0.67 | 7,792 | Both | 24816252 |
| Calcium | Metabolites | NA | Weighted mode | 137 | 3.529 | 1.956 | 0.073 | 0.67 | NA | Both | NA |
| X-11538 | Metabolites | Unknown metabolite | RE IVW | 3 | -0.689 | 0.203 | 0.077 | 0.68 | 7,804 | Both | 24816252 |
| Sphingomyelins | Metabolites | Lipid | Penalised median | 6 | -0.232 | 0.134 | 0.085 | 0.73 | 13,476 | Both | 27005778 |
| X-08402 | Metabolites | Unknown metabolite | RE IVW | 2 | -0.601 | 0.089 | 0.094 | 0.77 | 7,726 | Both | 24816252 |
| X-12728 | Metabolites | Unknown metabolite | RE IVW | 5 | -0.033 | 0.015 | 0.10 | 0.77 | 537 | Both | 24816252 |
| Dehydroisoandrosterone sulfate (DHEA-S) | Metabolites | Lipid | RE IVW | 2 | -0.341 | 0.056 | 0.10 | 0.77 | 7,793 | Both | 24816252 |
| Concentration of chylomicrons and largest VLDL particles | Metabolites | Lipid | RE IVW | 5 | -0.198 | 0.096 | 0.11 | 0.77 | 18,960 | Both | 27005778 |
| 1-linoleoylglycerophosphoethanolamine* | Metabolites | Lipid | RE IVW | 2 | -1.606 | 0.283 | 0.11 | 0.77 | 7,817 | Both | 24816252 |
| Free cholesterol to esterified cholesterol ratio | Metabolites | Lipid | Simple mode | 10 | -0.196 | 0.111 | 0.11 | 0.77 | 13,497 | Both | 27005778 |
| Serine | Metabolites | Amino acid | RE IVW | 2 | 0.792 | 0.141 | 0.11 | 0.77 | 7,796 | Both | 24816252 |
| Obesity class 3 | Risk factor | Anthropometric | RE IVW | 2 | 0.163 | 0.030 | 0.12 | 0.77 | 50,364 | Both | 23563607 |

| | | | | | | | | | | | |
|---|---|---|---|---|---|---|---|---|---|---|---|
| Omega-6 fatty acids | Metabolites | Fatty acid | Penalised median | 8 | -0.162 | 0.104 | 0.12 | 0.77 | 13,506 | Both | 27005778 |
| X-12092 | Metabolites | Unknown metabolite | RE IVW | 5 | 0.195 | 0.100 | 0.12 | 0.77 | 7,500 | Both | 24816252 |
| X-12188 | Metabolites | Unknown metabolite | RE IVW | 2 | 0.159 | 0.032 | 0.13 | 0.77 | 832 | Both | 24816252 |
| Valine | Metabolites | Amino acid | RE IVW | 5 | 0.277 | 0.147 | 0.13 | 0.77 | 24,900 | Both | 27005778 |
| X-12063 | Metabolites | Unknown metabolite | Simple median | 6 | -0.347 | 0.233 | 0.14 | 0.77 | 7,197 | Both | 24816252 |
| Laurate (12:0) | Metabolites | Fatty acid | RE IVW | 2 | 2.525 | 0.557 | 0.14 | 0.77 | 7,793 | Both | 24816252 |
| Succinylcarnitine | Metabolites | Energy | Simple mode | 7 | -1.563 | 0.919 | 0.14 | 0.77 | 6,948 | Both | 24816252 |
| Phospholipids in very large VLDL | Metabolites | Lipid | Simple median | 8 | -0.210 | 0.142 | 0.14 | 0.77 | 21,237 | Both | 27005778 |
| Total cholesterol in large VLDL | Metabolites | Lipid | Simple median | 7 | -0.255 | 0.174 | 0.14 | 0.77 | 21,235 | Both | 27005778 |
| X-11905 | Metabolites | Unknown metabolite | RE IVW | 2 | -0.726 | 0.167 | 0.14 | 0.77 | 4,761 | Both | 24816252 |
| Cholesterol | Metabolites | Lipid | RE IVW | 2 | -2.873 | 0.665 | 0.14 | 0.77 | 7,813 | Both | 24816252 |
| ADSGEGDFXAEGGGVR* | Metabolites | Peptide | RE IVW | 2 | -0.399 | 0.092 | 0.15 | 0.77 | 5,588 | Both | 24816252 |
| Acetylphosphate | Metabolites | Energy | RE IVW | 2 | -1.665 | 0.396 | 0.15 | 0.77 | 7,789 | Both | 24816252 |
| Palmitoyl sphingomyelin | Metabolites | Lipid | RE IVW | 2 | -3.063 | 0.733 | 0.15 | 0.77 | 7,814 | Both | 24816252 |
| Albumin | Metabolites | Protein | RE IVW | 2 | -0.248 | 0.062 | 0.16 | 0.77 | 18,960 | Both | 27005778 |
| Octadecanedioate | Metabolites | Lipid | RE IVW | 2 | -1.357 | 0.340 | 0.16 | 0.77 | 7,300 | Both | 24816252 |
| 5alpha-androstan-3beta,17beta-diol disulfate | Metabolites | Lipid | RE IVW | 3 | -0.366 | 0.168 | 0.16 | 0.77 | 7,345 | Both | 24816252 |
| Apoliprotein B | Metabolites | NA | RE Egger | 114 | -0.811 | 0.577 | 0.16 | 0.77 | NA | Both | NA |
| X-18601 | Metabolites | Unknown metabolite | RE IVW | 3 | -1.035 | 0.478 | 0.16 | 0.77 | 7,663 | Both | 24816252 |
| X-14304--leucylalanine | Metabolites | Peptide | RE IVW | 2 | 1.188 | 0.319 | 0.17 | 0.78 | 2,434 | Both | 24816252 |
| Free cholesterol in large VLDL | Metabolites | Lipid | Simple mean | 9 | -0.187 | 0.126 | 0.18 | 0.78 | 21,238 | Both | 27005778 |
| Direct bilirubin | Metabolites | NA | Simple median | 53 | -0.313 | 0.235 | 0.18 | 0.78 | NA | Both | NA |
| Alanine aminotransferase | Metabolites | NA | FE IVW | 73 | 0.010 | 0.008 | 0.18 | 0.78 | NA | Both | NA |
| Total fatty acids | Metabolites | Fatty acid | RE Egger | 10 | -0.758 | 0.523 | 0.19 | 0.78 | 13,505 | Both | 27005778 |
| X-14189--leucylalanine | Metabolites | Peptide | RE IVW | 2 | 1.041 | 0.315 | 0.19 | 0.78 | 2,745 | Both | 24816252 |
| Average number of methylene groups in a fatty acid chain | Metabolites | Fatty acid | RE IVW | 3 | -0.280 | 0.142 | 0.19 | 0.78 | 19,021 | Both | 27005778 |
| Triglycerides in chylomicrons and largest VLDL particles | Metabolites | Lipid | Simple median | 8 | -0.190 | 0.144 | 0.19 | 0.78 | 21,540 | Both | 27005778 |
| Omega-7, omega-9 and saturated fatty acids | Metabolites | Fatty acid | RE IVW | 5 | -0.133 | 0.087 | 0.20 | 0.81 | 13,506 | Both | 27005778 |
| Mono-unsaturated fatty acids | Metabolites | Fatty acid | RE IVW | 5 | -0.307 | 0.202 | 0.20 | 0.81 | 13,535 | Both | 27005778 |
| Lipoprotein A | Metabolites | NA | RE IVW | 4 | 0.003 | 0.002 | 0.21 | 0.81 | NA | Both | NA |
| X-03056--N-[3-(2-Oxopyrrolidin-1-yl)propyl]acetamide | Metabolites | Amino acid | RE IVW | 4 | -0.583 | 0.367 | 0.21 | 0.81 | 7,812 | Both | 24816252 |
| Tryptophan | Metabolites | Amino acid | Simple median | 17 | 2.003 | 1.608 | 0.21 | 0.81 | 7,804 | Both | 24816252 |
| X-11529 | Metabolites | Unknown metabolite | RE IVW | 2 | -0.207 | 0.072 | 0.21 | 0.81 | 6,664 | Both | 24816252 |
| Total lipids in medium VLDL | Metabolites | Lipid | Simple median | 6 | -0.176 | 0.142 | 0.22 | 0.81 | 19,273 | Both | 27005778 |
| Ratio of bisallylic groups to double bonds | Metabolites | Metabolites ratio | RE IVW | 4 | 0.166 | 0.107 | 0.22 | 0.81 | 13,524 | Both | 27005778 |
| X-11593--O-methylascorbate* | Metabolites | Cofactors and vitamins | Simple mean | 6 | -0.518 | 0.371 | 0.22 | 0.81 | 7,788 | Both | 24816252 |
| Triglycerides in medium VLDL | Metabolites | Lipid | Simple median | 9 | -0.160 | 0.133 | 0.23 | 0.81 | 21,241 | Both | 27005778 |
| X-11327 | Metabolites | Unknown metabolite | RE IVW | 2 | -2.262 | 0.854 | 0.23 | 0.81 | 7,671 | Both | 24816252 |
| Ratio of bisallylic groups to total fatty acids | Metabolites | Fatty acid | RE IVW | 4 | 0.243 | 0.163 | 0.23 | 0.81 | 13,171 | Both | 27005778 |
| Hydroxyisovaleroyl carnitine | Metabolites | Amino acid | RE IVW | 2 | 0.954 | 0.372 | 0.24 | 0.81 | 5,588 | Both | 24816252 |
| Average number of double bonds in a fatty acid chain | Metabolites | Fatty acid | RE IVW | 5 | 0.185 | 0.133 | 0.24 | 0.81 | 15,728 | Both | 27005778 |
| Mean diameter for LDL particles | Metabolites | Lipid | RE IVW | 3 | 0.118 | 0.070 | 0.24 | 0.81 | 19,273 | Both | 27005778 |
| Transferrin | Risk factor | Metal | Simple mode | 7 | 0.121 | 0.093 | 0.24 | 0.81 | 23,986 | Both | 25352340 |

| | | | | | | | | | | | |
|---|---|---|---|---|---|---|---|---|---|---|---|
| X-11793--oxidized bilirubin* | Metabolites | Cofactors and vitamins | RE IVW | 2 | 0.288 | 0.118 | 0.25 | 0.81 | 7,611 | Both | 24816252 |
| Citrulline | Metabolites | Amino acid | RE IVW | 4 | -1.799 | 1.258 | 0.25 | 0.81 | 7,773 | Both | 24816252 |
| X-02269 | Metabolites | Unknown metabolite | RE IVW | 3 | 0.668 | 0.419 | 0.25 | 0.81 | 7,701 | Both | 24816252 |
| 4-androsten-3beta,17beta-diol disulfate 1* | Metabolites | Lipid | RE IVW | 2 | -0.124 | 0.052 | 0.25 | 0.81 | 7,804 | Both | 24816252 |
| Other polyunsaturated fatty acids than 18:2 | Metabolites | Fatty acid | Weighted mode | 8 | -0.192 | 0.156 | 0.26 | 0.81 | 13,549 | Both | 27005778 |
| Phospholipids in IDL | Metabolites | Lipid | RE IVW | 17 | -0.061 | 0.052 | 0.26 | 0.81 | 21,559 | Both | 27005778 |
| Body fat | Risk factor | Anthropometric | Simple mean | 10 | 0.416 | 0.347 | 0.26 | 0.81 | 100,716 | Both | 26833246 |
| Average number of methylene groups per double bond | Metabolites | Metabolites ratio | RE IVW | 5 | -0.161 | 0.123 | 0.26 | 0.81 | 13,532 | Both | 27005778 |
| Glutaroyl carnitine | Metabolites | Amino acid | Simple median | 7 | -0.571 | 0.522 | 0.27 | 0.83 | 7,701 | Both | 24816252 |
| N-acetylglycine | Metabolites | Amino acid | RE IVW | 2 | 0.142 | 0.066 | 0.28 | 0.83 | 7,135 | Both | 24816252 |
| 1-eicosatrienoylglycerophosphocholine* | Metabolites | Lipid | RE IVW | 2 | -0.741 | 0.346 | 0.28 | 0.83 | 7,809 | Both | 24816252 |
| Gamma-glutamyltyrosine | Metabolites | Peptide | RE IVW | 5 | -1.646 | 1.323 | 0.28 | 0.83 | 7,468 | Both | 24816252 |
| Cis-4-decenoyl carnitine | Metabolites | Lipid | RE IVW | 2 | 0.500 | 0.241 | 0.29 | 0.84 | 7,660 | Both | 24816252 |
| Creatinine | Metabolites | Amino acid | Weighted mode | 6 | -0.313 | 0.267 | 0.29 | 0.85 | 24,810 | Both | 27005778 |
| 1,5-anhydroglucitol (1,5-AG) | Metabolites | Carbohydrate | RE IVW | 3 | -0.850 | 0.613 | 0.30 | 0.85 | 7,746 | Both | 24816252 |
| 18:2, linoleic acid (LA) | Metabolites | Fatty acid | Simple mode | 10 | -0.171 | 0.156 | 0.30 | 0.85 | 13,527 | Both | 27005778 |
| Triglycerides in small VLDL | Metabolites | Lipid | Simple median | 10 | -0.132 | 0.129 | 0.31 | 0.86 | 21,558 | Both | 27005778 |
| X-11423--O-sulfo-L-tyrosine | Metabolites | Amino acid | RE IVW | 2 | 0.959 | 0.520 | 0.32 | 0.87 | 7,765 | Both | 24816252 |
| Aspartate aminotransferase | Metabolites | NA | Simple mode | 164 | 0.314 | 0.313 | 0.32 | 0.87 | NA | Both | NA |
| Immunoglobulin G index levels | Risk factor | Immune system | RE IVW | 2 | 0.023 | 0.013 | 0.32 | 0.87 | 938 | Both | 25616667 |
| Triglycerides in very small VLDL | Metabolites | Lipid | Simple median | 14 | -0.096 | 0.098 | 0.32 | 0.87 | 19,273 | Both | 27005778 |
| Heptanoate (7:0) | Metabolites | Fatty acid | RE IVW | 2 | 0.699 | 0.397 | 0.33 | 0.88 | 7,802 | Both | 24816252 |
| Total lipids in small LDL | Metabolites | Lipid | Simple median | 12 | -0.086 | 0.090 | 0.34 | 0.88 | 19,273 | Both | 27005778 |
| Free cholesterol in IDL | Metabolites | Lipid | RE IVW | 16 | -0.061 | 0.062 | 0.34 | 0.88 | 21,559 | Both | 27005778 |
| X-11469 | Metabolites | Unknown metabolite | RE IVW | 3 | 0.593 | 0.483 | 0.34 | 0.88 | 7,779 | Both | 24816252 |
| Total cholesterol in very large HDL | Metabolites | Lipid | Simple median | 8 | 0.108 | 0.114 | 0.35 | 0.88 | 21,540 | Both | 27005778 |
| Birth weight | Risk factor | Anthropometric | Simple mode | 37 | -0.342 | 0.360 | 0.35 | 0.88 | 143,677 | Both | 27680694 |
| Zinc | Risk factor | Metal | RE IVW | 2 | -0.146 | 0.089 | 0.35 | 0.88 | 2,603 | Both | 23720494 |
| Isobutyrylcarnitine | Metabolites | Amino acid | RE IVW | 3 | -0.369 | 0.306 | 0.35 | 0.88 | 7,812 | Both | 24816252 |
| Serum total cholesterol | Metabolites | Lipid | RE IVW | 18 | -0.091 | 0.098 | 0.37 | 0.89 | 21,491 | Both | 27005778 |
| Asparagine | Metabolites | Amino acid | RE IVW | 2 | 0.161 | 0.105 | 0.37 | 0.89 | 7,761 | Both | 24816252 |
| Cholesterol esters in very large HDL | Metabolites | Lipid | Weighted median | 7 | 0.098 | 0.110 | 0.37 | 0.89 | 19,273 | Both | 27005778 |
| X-11445--5-alpha-pregnan-3beta,20alpha-disulfate | Metabolites | Lipid | RE IVW | 3 | -0.422 | 0.374 | 0.38 | 0.89 | 2,570 | Both | 24816252 |
| Concentration of large VLDL particles | Metabolites | Lipid | Simple mean | 8 | -0.145 | 0.157 | 0.39 | 0.89 | 18,960 | Both | 27005778 |
| X-12556 | Metabolites | Unknown metabolite | RE IVW | 2 | 0.586 | 0.408 | 0.39 | 0.89 | 7,483 | Both | 24816252 |
| X-12644 | Metabolites | Unknown metabolite | RE IVW | 3 | -1.390 | 1.303 | 0.40 | 0.89 | 7,795 | Both | 24816252 |
| X-12100--hydroxytryptophan* | Metabolites | Amino acid | RE IVW | 2 | 0.336 | 0.243 | 0.40 | 0.89 | 7,499 | Both | 24816252 |
| ADpSGEGDFXAEGGGVR* | Metabolites | Peptide | RE IVW | 2 | -0.234 | 0.169 | 0.40 | 0.89 | 3,939 | Both | 24816252 |
| X-13435 | Metabolites | Unknown metabolite | RE IVW | 2 | -0.793 | 0.576 | 0.40 | 0.89 | 6,970 | Both | 24816252 |
| Free cholesterol in medium VLDL | Metabolites | Lipid | Simple median | 7 | -0.128 | 0.153 | 0.40 | 0.89 | 21,240 | Both | 27005778 |
| Triglycerides in IDL | Metabolites | Lipid | Simple mean | 16 | 0.054 | 0.064 | 0.41 | 0.89 | 19,273 | Both | 27005778 |
| Pyroglutamine* | Metabolites | Amino acid | RE IVW | 4 | -0.278 | 0.292 | 0.41 | 0.89 | 7,800 | Both | 24816252 |
| X-03094 | Metabolites | Unknown metabolite | RE IVW | 5 | -1.270 | 1.399 | 0.42 | 0.89 | 7,804 | Both | 24816252 |

| | | | | | | | | | | | |
|---|---|---|---|---|---|---|---|---|---|---|---|
| Gamma-glutamylglutamine | Metabolites | Peptide | RE IVW | 3 | 2.340 | 2.342 | 0.42 | 0.89 | 7,662 | Both | 24816252 |
| Concentration of small VLDL particles | Metabolites | Lipid | RE IVW | 13 | -0.072 | 0.088 | 0.43 | 0.89 | 19,273 | Both | 27005778 |
| X-11261 | Metabolites | Unknown metabolite | RE IVW | 2 | -0.486 | 0.392 | 0.43 | 0.89 | 7,771 | Both | 24816252 |
| X-12696 | Metabolites | Unknown metabolite | RE IVW | 2 | -0.886 | 0.730 | 0.44 | 0.89 | 7,409 | Both | 24816252 |
| 4-androsten-3beta,17beta-diol disulfate 2* | Metabolites | Lipid | RE IVW | 2 | -0.670 | 0.556 | 0.44 | 0.89 | 7,776 | Both | 24816252 |
| 1-arachidonoylglycerophosphoethanolamine* | Metabolites | Lipid | RE IVW | 3 | 1.179 | 1.242 | 0.44 | 0.89 | 7,798 | Both | 24816252 |
| Phospholipids in large HDL | Metabolites | Lipid | Simple median | 9 | 0.080 | 0.105 | 0.45 | 0.89 | 19,273 | Both | 27005778 |
| Octanoylcarnitine | Metabolites | Lipid | RE IVW | 3 | 0.342 | 0.365 | 0.45 | 0.89 | 7,790 | Both | 24816252 |
| Total lipids in large HDL | Metabolites | Lipid | Simple median | 9 | 0.077 | 0.103 | 0.45 | 0.89 | 19,273 | Both | 27005778 |
| X-11440 | Metabolites | Unknown metabolite | RE IVW | 3 | -0.112 | 0.123 | 0.46 | 0.89 | 7,686 | Both | 24816252 |
| Serum total triglycerides | Metabolites | Lipid | Simple mode | 8 | -0.155 | 0.198 | 0.46 | 0.89 | 21,545 | Both | 27005778 |
| Lactate | Metabolites | Carbohydrate | RE IVW | 2 | -0.950 | 0.843 | 0.46 | 0.89 | 24,871 | Both | 27005778 |
| Acetylcarnitine | Metabolites | Lipid | RE IVW | 2 | 0.299 | 0.265 | 0.46 | 0.89 | 7,805 | Both | 24816252 |
| X-08988 | Metabolites | Unknown metabolite | RE IVW | 2 | 0.665 | 0.590 | 0.46 | 0.89 | 7,776 | Both | 24816252 |
| Concentration of large HDL particles | Metabolites | Lipid | Simple median | 9 | 0.076 | 0.104 | 0.47 | 0.89 | 19,273 | Both | 27005778 |
| Mean diameter for HDL particles | Metabolites | Lipid | Simple median | 11 | 0.064 | 0.090 | 0.47 | 0.89 | 19,273 | Both | 27005778 |
| Histidine | Metabolites | Amino acid | RE IVW | 5 | -0.129 | 0.165 | 0.48 | 0.89 | 19,244 | Both | 27005778 |
| Phospholipids in medium VLDL | Metabolites | Lipid | Simple median | 9 | -0.106 | 0.150 | 0.48 | 0.89 | 21,240 | Both | 27005778 |
| HDL cholesterol | Metabolites | NA | FE Egger | 171 | -0.216 | 0.310 | 0.49 | 0.89 | NA | Both | NA |
| Decanoylcarnitine | Metabolites | Lipid | RE IVW | 3 | 0.359 | 0.424 | 0.49 | 0.89 | 7,766 | Both | 24816252 |
| X-12093 | Metabolites | Unknown metabolite | RE IVW | 2 | 0.204 | 0.201 | 0.50 | 0.89 | 2,854 | Both | 24816252 |
| Caprylate (8:0) | Metabolites | Fatty acid | RE IVW | 2 | -3.331 | 3.300 | 0.50 | 0.89 | 7,802 | Both | 24816252 |
| Leptin | Risk factor | Hormone | Wald ratio | 1 | 0.636 | 0.636 | 0.50 | 0.89 | 32,161 | Both | 26833098 |
| X-11334 | Metabolites | Unknown metabolite | Wald ratio | 1 | 1.407 | 1.407 | 0.50 | 0.89 | 5,462 | Both | 24816252 |
| X-09789 | Metabolites | Unknown metabolite | Wald ratio | 1 | 0.808 | 0.808 | 0.50 | 0.89 | 7,805 | Both | 24816252 |
| X-11444 | Metabolites | Unknown metabolite | Wald ratio | 1 | 0.253 | 0.253 | 0.50 | 0.89 | 7,758 | Both | 24816252 |
| 1-arachidonoylglycerophosphocholine* | Metabolites | Lipid | Wald ratio | 1 | 1.370 | 1.370 | 0.50 | 0.89 | 7,507 | Both | 24816252 |
| Epiandrosterone sulfate | Metabolites | Lipid | Wald ratio | 1 | -0.199 | -0.199 | 0.50 | 0.89 | 7,769 | Both | 24816252 |
| X-13859 | Metabolites | Unknown metabolite | Wald ratio | 1 | 3.673 | 3.673 | 0.50 | 0.89 | 7,002 | Both | 24816252 |
| Glucose | Metabolites | Carbohydrate | Wald ratio | 1 | 0.650 | 0.650 | 0.50 | 0.89 | 24,679 | Both | 27005778 |
| Isoleucine | Metabolites | Amino acid | Wald ratio | 1 | -1.166 | -1.166 | 0.50 | 0.89 | 24,776 | Both | 27005778 |
| Leucine | Metabolites | Amino acid | Wald ratio | 1 | -0.556 | -0.556 | 0.50 | 0.89 | 24,728 | Both | 27005778 |
| Total bilirubin | Metabolites | NA | Simple median | 57 | -0.031 | 0.046 | 0.51 | 0.89 | NA | Both | NA |
| Glutamine | Metabolites | Amino acid | RE IVW | 5 | 0.168 | 0.230 | 0.51 | 0.89 | 24,462 | Both | 27005778 |
| 10-undecenoate (11:1n1) | Metabolites | Fatty acid | RE IVW | 2 | 0.846 | 0.877 | 0.51 | 0.89 | 7,806 | Both | 24816252 |
| Erythronate* | Metabolites | Carbohydrate | RE IVW | 3 | 0.665 | 0.858 | 0.52 | 0.89 | 7,752 | Both | 24816252 |
| Phospholipids in chylomicrons and largest VLDL particles | Metabolites | Lipid | Simple mode | 6 | -0.149 | 0.217 | 0.52 | 0.89 | 21,542 | Both | 27005778 |
| Proline | Metabolites | Amino acid | RE IVW | 3 | -0.201 | 0.262 | 0.52 | 0.89 | 7,816 | Both | 24816252 |
| Height | Risk factor | Anthropometric | RE IVW | 52 | -0.063 | 0.101 | 0.54 | 0.89 | 73,137 | Women | 23754948 |
| X-12855 | Metabolites | Unknown metabolite | RE IVW | 2 | -0.387 | 0.434 | 0.54 | 0.89 | 5,120 | Both | 24816252 |
| X-11204 | Metabolites | Unknown metabolite | RE IVW | 2 | 1.569 | 1.761 | 0.54 | 0.89 | 7,799 | Both | 24816252 |
| 4-methyl-2-oxopentanoate | Metabolites | Amino acid | RE IVW | 2 | 2.059 | 2.313 | 0.54 | 0.89 | 7,776 | Both | 24816252 |
| Gamma-glutamylphenylalanine | Metabolites | Peptide | RE IVW | 2 | 1.919 | 2.157 | 0.54 | 0.89 | 7,753 | Both | 24816252 |

| | | | | | | | | | | | |
|---|---|---|---|---|---|---|---|---|---|---|---|
| 22:6, docosahexaenoic acid | Metabolites | Fatty acid | RE IVW | 5 | 0.133 | 0.199 | 0.54 | 0.89 | 13,499 | Both | 27005778 |
| Isovalerylcarnitine | Metabolites | Amino acid | RE IVW | 3 | 0.302 | 0.416 | 0.54 | 0.89 | 7,789 | Both | 24816252 |
| X-13431--nonanoylcarnitine* | Metabolites | Lipid | RE IVW | 2 | 0.080 | 0.092 | 0.54 | 0.89 | 6,591 | Both | 24816252 |
| Concentration of small LDL particles | Metabolites | Lipid | Simple median | 9 | -0.055 | 0.091 | 0.55 | 0.89 | 19,273 | Both | 27005778 |
| N-acetylornithine | Metabolites | Amino acid | RE IVW | 2 | 0.238 | 0.277 | 0.55 | 0.89 | 7,574 | Both | 24816252 |
| Betaine | Metabolites | Amino acid | RE IVW | 3 | -0.282 | 0.397 | 0.55 | 0.89 | 7,806 | Both | 24816252 |
| X-11315 | Metabolites | Unknown metabolite | RE IVW | 3 | 0.360 | 0.525 | 0.56 | 0.90 | 7,785 | Both | 24816252 |
| Total cholesterol in HDL | Metabolites | Lipid | Simple median | 11 | 0.079 | 0.137 | 0.57 | 0.90 | 21,555 | Both | 27005778 |
| Acetoacetate | Metabolites | Keto acid | RE IVW | 2 | 0.407 | 0.522 | 0.58 | 0.91 | 19,262 | Both | 27005778 |
| Free cholesterol in large HDL | Metabolites | Lipid | RE IVW | 13 | 0.055 | 0.098 | 0.59 | 0.92 | 21,559 | Both | 27005778 |
| Phospholipids in very large HDL | Metabolites | Lipid | RE IVW | 15 | 0.041 | 0.075 | 0.59 | 0.92 | 19,273 | Both | 27005778 |
| Vitamin D | Metabolites | NA | Simple mean | 50 | -0.063 | 0.119 | 0.60 | 0.92 | NA | Both | NA |
| Phospholipids in medium HDL | Metabolites | Lipid | RE IVW | 5 | 0.073 | 0.129 | 0.60 | 0.92 | 21,558 | Both | 27005778 |
| Total lipids in IDL | Metabolites | Lipid | Penalised median | 16 | -0.022 | 0.041 | 0.60 | 0.92 | 19,273 | Both | 27005778 |
| Dihomo-linolenate (20:3n3 or n6) | Metabolites | Fatty acid | RE IVW | 2 | -0.617 | 0.855 | 0.60 | 0.92 | 7,805 | Both | 24816252 |
| Levulinate (4-oxovalerate) | Metabolites | Amino acid | RE IVW | 2 | -0.486 | 0.679 | 0.60 | 0.92 | 6,982 | Both | 24816252 |
| Glycoprotein acetyls | Metabolites | Protein | RE IVW | 5 | -0.151 | 0.272 | 0.61 | 0.92 | 19,270 | Both | 27005778 |
| Concentration of medium HDL particles | Metabolites | Lipid | RE IVW | 4 | 0.099 | 0.176 | 0.61 | 0.92 | 19,273 | Both | 27005778 |
| Total lipids in small VLDL | Metabolites | Lipid | Simple median | 9 | -0.048 | 0.097 | 0.62 | 0.93 | 19,273 | Both | 27005778 |
| Total lipids in very large HDL | Metabolites | Lipid | Weighted median | 13 | -0.038 | 0.078 | 0.62 | 0.93 | 19,273 | Both | 27005778 |
| Birth length | Risk factor | Anthropometric | RE IVW | 2 | 0.096 | 0.145 | 0.63 | 0.93 | 28,459 | Both | 25281659 |
| Total lipids in small HDL | Metabolites | Lipid | RE IVW | 6 | -0.105 | 0.205 | 0.63 | 0.93 | 19,273 | Both | 27005778 |
| Total protein | Metabolites | NA | Simple mode | 162 | -0.175 | 0.368 | 0.63 | 0.93 | NA | Both | NA |
| 4-acetamidobutanoate | Metabolites | Amino acid | RE IVW | 2 | 0.586 | 0.925 | 0.64 | 0.93 | 6,930 | Both | 24816252 |
| Total lipids in medium HDL | Metabolites | Lipid | RE IVW | 4 | 0.085 | 0.168 | 0.65 | 0.94 | 19,273 | Both | 27005778 |
| X-11792 | Metabolites | Unknown metabolite | RE IVW | 3 | -0.063 | 0.121 | 0.65 | 0.94 | 2,442 | Both | 24816252 |
| Tyrosine | Metabolites | Amino acid | RE IVW | 3 | -0.064 | 0.123 | 0.66 | 0.94 | 24,925 | Both | 27005778 |
| Phospholipids in large VLDL | Metabolites | Lipid | Penalised median | 6 | -0.058 | 0.134 | 0.66 | 0.94 | 21,239 | Both | 27005778 |
| Free cholesterol | Metabolites | Lipid | Penalised median | 10 | -0.018 | 0.042 | 0.67 | 0.94 | 13,497 | Both | 27005778 |
| Total lipids in very small VLDL | Metabolites | Lipid | RE IVW | 16 | 0.016 | 0.037 | 0.67 | 0.94 | 19,273 | Both | 27005778 |
| Phenylalanine | Metabolites | Amino acid | RE IVW | 3 | -0.061 | 0.128 | 0.68 | 0.94 | 22,663 | Both | 27005778 |
| IGF-1 | Metabolites | NA | Weighted mode | 236 | -0.064 | 0.156 | 0.68 | 0.94 | NA | Both | NA |
| Kynurenine | Metabolites | Amino acid | RE IVW | 3 | -0.531 | 1.127 | 0.68 | 0.94 | 7,816 | Both | 24816252 |
| Extreme height | Risk factor | Anthropometric | Simple median | 41 | -0.012 | 0.031 | 0.69 | 0.94 | 16,196 | Both | 23563607 |
| Free cholesterol in small VLDL | Metabolites | Lipid | Simple median | 10 | -0.036 | 0.092 | 0.69 | 0.94 | 21,559 | Both | 27005778 |
| Concentration of small HDL particles | Metabolites | Lipid | RE IVW | 4 | -0.118 | 0.274 | 0.70 | 0.94 | 19,273 | Both | 27005778 |
| Concentration of very large HDL particles | Metabolites | Lipid | Simple median | 8 | 0.038 | 0.098 | 0.70 | 0.94 | 19,273 | Both | 27005778 |
| Total phosphoglycerides | Metabolites | Lipid | RE IVW | 7 | -0.070 | 0.172 | 0.70 | 0.94 | 13,519 | Both | 27005778 |
| Stearate (18:0) | Metabolites | Fatty acid | RE IVW | 2 | 1.431 | 2.833 | 0.70 | 0.94 | 7,803 | Both | 24816252 |
| Alkaline phosphatase | Metabolites | NA | Weighted mode | 168 | 0.002 | 0.004 | 0.70 | 0.94 | NA | Both | NA |
| Alpha-hydroxyisovalerate | Metabolites | Amino acid | RE IVW | 3 | 0.418 | 0.965 | 0.71 | 0.94 | 7,668 | Both | 24816252 |
| Cystatin C | Metabolites | NA | Simple mode | 226 | -0.128 | 0.349 | 0.72 | 0.94 | NA | Both | NA |
| SHBG | Metabolites | NA | Penalised mode | 139 | 0.002 | 0.007 | 0.72 | 0.94 | NA | Both | NA |

| | | | | | | | | | | | |
|---|---|---|---|---|---|---|---|---|---|---|---|
| Bradykinin, des-arg(9) | Metabolites | Peptide | RE IVW | 3 | -0.039 | 0.094 | 0.72 | 0.94 | 4,570 | Both | 24816252 |
| Total cholesterol in IDL | Metabolites | Lipid | Simple median | 17 | -0.028 | 0.077 | 0.72 | 0.94 | 19,273 | Both | 27005778 |
| 1-arachidonoylglycerophosphoinositol* | Metabolites | Lipid | RE IVW | 4 | -0.509 | 1.313 | 0.72 | 0.94 | 7,797 | Both | 24816252 |
| HWESASXX* | Metabolites | Peptide | RE IVW | 2 | -0.310 | 0.683 | 0.73 | 0.94 | 7,700 | Both | 24816252 |
| Total cholesterol in medium VLDL | Metabolites | Lipid | Simple median | 9 | -0.042 | 0.122 | 0.73 | 0.94 | 21,551 | Both | 27005778 |
| Phospholipids in very small VLDL | Metabolites | Lipid | RE IVW | 16 | -0.017 | 0.053 | 0.75 | 0.94 | 19,273 | Both | 27005778 |
| Cholesterol esters in medium VLDL | Metabolites | Lipid | Penalised median | 10 | -0.032 | 0.100 | 0.75 | 0.94 | 19,273 | Both | 27005778 |
| Testosterone | Metabolites | NA | Simple mode | 62 | -0.223 | 0.706 | 0.75 | 0.94 | NA | Both | NA |
| Myo-inositol | Metabolites | Lipid | RE IVW | 2 | -0.931 | 2.352 | 0.76 | 0.94 | 7,803 | Both | 24816252 |
| Citrate | Metabolites | Energy | RE IVW | 5 | 0.038 | 0.116 | 0.76 | 0.94 | 24,770 | Both | 27005778 |
| Total cholesterol in small LDL | Metabolites | Lipid | Penalised median | 12 | -0.014 | 0.045 | 0.76 | 0.94 | 21,556 | Both | 27005778 |
| 1-palmitoylglycerophosphoethanolamine | Metabolites | Lipid | RE IVW | 2 | -0.317 | 0.816 | 0.76 | 0.94 | 7,763 | Both | 24816252 |
| X-12844 | Metabolites | Unknown metabolite | RE IVW | 3 | 0.058 | 0.169 | 0.76 | 0.94 | 7,768 | Both | 24816252 |
| Phospholipids in small VLDL | Metabolites | Lipid | Penalised median | 10 | -0.030 | 0.099 | 0.76 | 0.94 | 21,551 | Both | 27005778 |
| Phospholipids in medium LDL | Metabolites | Lipid | Simple median | 13 | -0.027 | 0.090 | 0.77 | 0.94 | 21,558 | Both | 27005778 |
| Free cholesterol in large LDL | Metabolites | Lipid | Simple median | 15 | -0.025 | 0.084 | 0.77 | 0.94 | 21,555 | Both | 27005778 |
| Total cholesterol in LDL | Metabolites | Lipid | Simple median | 17 | -0.025 | 0.083 | 0.77 | 0.94 | 21,559 | Both | 27005778 |
| Apoliprotein A | Metabolites | NA | Simple mode | 160 | 0.294 | 0.989 | 0.77 | 0.94 | NA | Both | NA |
| Apolipoprotein B | Metabolites | Protein | Simple mode | 13 | -0.030 | 0.106 | 0.78 | 0.95 | 20,690 | Both | 27005778 |
| Cholesterol esters in large VLDL | Metabolites | Lipid | Simple median | 19 | 0.023 | 0.083 | 0.78 | 0.95 | 19,273 | Both | 27005778 |
| Androsterone sulfate | Metabolites | Lipid | RE IVW | 3 | -0.055 | 0.189 | 0.80 | 0.96 | 7,785 | Both | 24816252 |
| Concentration of very small VLDL particles | Metabolites | Lipid | RE IVW | 16 | -0.018 | 0.069 | 0.80 | 0.96 | 19,273 | Both | 27005778 |
| Propionylcarnitine | Metabolites | Lipid | RE IVW | 4 | -0.198 | 0.725 | 0.80 | 0.96 | 7,813 | Both | 24816252 |
| Triglycerides in small HDL | Metabolites | Lipid | Weighted median | 8 | 0.027 | 0.109 | 0.80 | 0.96 | 21,558 | Both | 27005778 |
| X-12524 | Metabolites | Unknown metabolite | RE IVW | 2 | 0.276 | 0.875 | 0.81 | 0.96 | 7,809 | Both | 24816252 |
| LDL direct | Metabolites | NA | Simple mode | 105 | 0.078 | 0.337 | 0.82 | 0.96 | NA | Both | NA |
| 3-dehydrocarnitine* | Metabolites | Lipid | RE IVW | 2 | -0.266 | 0.918 | 0.82 | 0.96 | 7,809 | Both | 24816252 |
| Extreme waist-to-hip ratio | Risk factor | Anthropometric | RE IVW | 2 | 0.018 | 0.063 | 0.82 | 0.96 | 10,255 | Both | 23563607 |
| Weight | Risk factor | Anthropometric | Simple median | 10 | 0.055 | 0.246 | 0.82 | 0.96 | 73,137 | Women | 23754948 |
| X-13496 | Metabolites | Unknown metabolite | RE IVW | 2 | 0.904 | 3.272 | 0.83 | 0.96 | 7,656 | Both | 24816252 |
| Triglycerides in very large HDL | Metabolites | Lipid | FE IVW | 12 | 0.007 | 0.032 | 0.83 | 0.96 | 21,536 | Both | 27005778 |
| Total cholesterol in large LDL | Metabolites | Lipid | Penalised median | 18 | -0.008 | 0.039 | 0.83 | 0.96 | 21,552 | Both | 27005778 |
| C-reactive protein | Metabolites | NA | Penalised median | 27 | -0.011 | 0.054 | 0.84 | 0.96 | NA | Both | NA |
| Cholesterol esters in large HDL | Metabolites | Lipid | Simple median | 10 | 0.020 | 0.098 | 0.84 | 0.96 | 19,273 | Both | 27005778 |
| Total lipids in very large VLDL | Metabolites | Lipid | Weighted median | 7 | -0.028 | 0.139 | 0.84 | 0.96 | 19,273 | Both | 27005778 |
| Phospholipids in large LDL | Metabolites | Lipid | Simple median | 18 | -0.015 | 0.079 | 0.85 | 0.96 | 21,550 | Both | 27005778 |
| Cholesterol esters in medium LDL | Metabolites | Lipid | Weighted median | 17 | -0.007 | 0.042 | 0.86 | 0.96 | 19,273 | Both | 27005778 |
| Butyrylcarnitine | Metabolites | Lipid | RE IVW | 5 | -0.042 | 0.223 | 0.86 | 0.96 | 7,796 | Both | 24816252 |
| Carnitine | Metabolites | Lipid | Simple median | 15 | -0.221 | 1.274 | 0.86 | 0.96 | 7,797 | Both | 24816252 |
| Transferrin Saturation | Risk factor | Metal | RE IVW | 3 | -0.007 | 0.036 | 0.87 | 0.96 | 23,986 | Both | 25352340 |
| 3-methyl-2-oxovalerate | Metabolites | Amino acid | RE IVW | 3 | 0.775 | 4.126 | 0.87 | 0.96 | 7,779 | Both | 24816252 |
| Apolipoprotein A-I | Metabolites | Protein | Penalised median | 7 | 0.017 | 0.102 | 0.87 | 0.96 | 20,687 | Both | 27005778 |
| X-12244--N-acetylcarnosine | Metabolites | Peptide | RE IVW | 3 | 0.099 | 0.533 | 0.87 | 0.96 | 6,608 | Both | 24816252 |

| | | | | | | | | | | | |
|---|---|---|---|---|---|---|---|---|---|---|---|
| Ferritin | Risk factor | Metal | RE IVW | 4 | 0.016 | 0.094 | 0.88 | 0.96 | 23,986 | Both | 25352340 |
| Glycoproteins | Metabolites | Protein | Simple median | 7 | 0.018 | 0.119 | 0.88 | 0.96 | 18,734 | Both | 27005778 |
| Total cholesterol in medium LDL | Metabolites | Lipid | Simple median | 16 | 0.011 | 0.081 | 0.89 | 0.96 | 21,559 | Both | 27005778 |
| Total lipids in medium LDL | Metabolites | Lipid | Simple median | 16 | 0.011 | 0.085 | 0.89 | 0.96 | 19,273 | Both | 27005778 |
| Concentration of medium LDL particles | Metabolites | Lipid | Simple median | 14 | 0.012 | 0.090 | 0.90 | 0.96 | 19,273 | Both | 27005778 |
| Cholesterol esters in medium HDL | Metabolites | Lipid | RE IVW | 6 | 0.016 | 0.117 | 0.90 | 0.96 | 19,273 | Both | 27005778 |
| Total lipids in large LDL | Metabolites | Lipid | Simple median | 18 | 0.010 | 0.082 | 0.90 | 0.96 | 19,273 | Both | 27005778 |
| Concentration of very large VLDL particles | Metabolites | Lipid | Penalised median | 6 | -0.017 | 0.141 | 0.90 | 0.96 | 18,960 | Both | 27005778 |
| X-12850 | Metabolites | Unknown metabolite | RE IVW | 3 | 0.056 | 0.409 | 0.90 | 0.96 | 6,251 | Both | 24816252 |
| Iron | Risk factor | Metal | RE IVW | 3 | -0.005 | 0.047 | 0.92 | 0.98 | 23,986 | Both | 25352340 |
| Total cholesterol in small VLDL | Metabolites | Lipid | Penalised median | 11 | -0.006 | 0.055 | 0.92 | 0.98 | 21,557 | Both | 27005778 |
| 2-aminobutyrate | Metabolites | Amino acid | RE IVW | 2 | -0.142 | 1.259 | 0.93 | 0.98 | 7,814 | Both | 24816252 |
| Phosphatidylcholine and other cholines | Metabolites | Lipid | RE IVW | 4 | -0.006 | 0.073 | 0.94 | 0.98 | 13,542 | Both | 27005778 |
| Copper | Risk factor | Metal | RE IVW | 2 | 0.007 | 0.073 | 0.94 | 0.98 | 2,603 | Both | 23720494 |
| X-11787 | Metabolites | Unknown metabolite | RE IVW | 2 | -0.047 | 0.574 | 0.95 | 0.98 | 7,811 | Both | 24816252 |
| Mean diameter for VLDL particles | Metabolites | Lipid | Simple median | 9 | 0.007 | 0.112 | 0.95 | 0.98 | 19,273 | Both | 27005778 |
| Concentration of medium VLDL particles | Metabolites | Lipid | Weighted median | 11 | -0.007 | 0.115 | 0.95 | 0.98 | 19,273 | Both | 27005778 |
| Total lipids in chylomicrons and largest VLDL particles | Metabolites | Lipid | Simple median | 7 | 0.009 | 0.170 | 0.96 | 0.98 | 18,960 | Both | 27005778 |
| X-13548 | Metabolites | Unknown metabolite | RE IVW | 2 | -0.165 | 2.646 | 0.96 | 0.98 | 6,022 | Both | 24816252 |
| 2-hydroxyisobutyrate | Metabolites | Amino acid | RE IVW | 3 | -0.020 | 0.359 | 0.96 | 0.98 | 6,539 | Both | 24816252 |
| Total cholesterol in medium HDL | Metabolites | Lipid | RE IVW | 5 | 0.007 | 0.159 | 0.97 | 0.98 | 21,558 | Both | 27005778 |
| Total cholesterol in large HDL | Metabolites | Lipid | Weighted mode | 10 | -0.003 | 0.085 | 0.97 | 0.98 | 21,558 | Both | 27005778 |
| Concentration of IDL particles | Metabolites | Lipid | Simple median | 15 | -0.003 | 0.080 | 0.97 | 0.98 | 19,273 | Both | 27005778 |
| Concentration of large LDL particles | Metabolites | Lipid | Simple median | 19 | -0.003 | 0.080 | 0.97 | 0.98 | 19,273 | Both | 27005778 |
| Free cholesterol in very large HDL | Metabolites | Lipid | Simple median | 15 | -0.003 | 0.102 | 0.97 | 0.98 | 21,542 | Both | 27005778 |
| Omega-3 fatty acids | Metabolites | Fatty acid | RE IVW | 5 | 0.006 | 0.286 | 0.98 | 0.99 | 13,544 | Both | 27005778 |
| Free cholesterol in medium HDL | Metabolites | Lipid | RE IVW | 4 | -0.003 | 0.174 | 0.99 | 0.99 | 21,559 | Both | 27005778 |
